# The effects of reduced nicotine content cigarettes in smokers with mood or anxiety disorders: a double-blind randomized trial

**DOI:** 10.1101/2022.05.24.22275536

**Authors:** Jonathan Foulds, Susan Veldheer, Gladys Pachas, Shari Hrabovsky, Ahmad Hameed, Sophia I Allen, Corinne Cather, Nour Azzouz, Jessica Yingst, Erin Hammett, Jennifer Modesto, Nicolle M Krebs, Courtney Lester, Neil Trushin, Lisa Reinhart, Emily Wasserman, Junjia Zhu, Jason Liao, Joshua E Muscat, John P Richie, A Eden Evins

## Abstract

**BACKGROUND:** The U.S. Food and Drug Administration and the government of New Zealand have proposed a reduction of the nicotine content in cigarettes to very low levels. This study examined the likely effects of this regulation in smokers with affective disorders.

**METHODS:** In a randomized controlled trial conducted at two sites (Penn State Hershey and Massachusetts General Hospital, Boston) 188 adult smokers with a current or lifetime anxiety or unipolar mood disorder, not planning to quit in the next 6 months, were randomly assigned to smoke either Usual Nicotine Content (UNC) (11.6 mg nicotine/cigarette) cigarettes, or Reduced Nicotine Content (RNC) cigarettes where the nicotine content per cigarette was progressively reduced to 0.2 mg in five steps over 18 weeks. Participants were then offered the choice to either receive assistance to quit smoking, receive free research cigarettes, or resume using their own cigarette brand during a 12-week follow-up period. Main outcomes were biomarkers of nicotine and toxicant exposure, smoking behavior and dependence and severity of psychiatric symptoms.

**RESULTS:** After switching to the lowest nicotine content cigarettes, compared to smokers in the UNC group, the RNC group had significantly lower plasma cotinine (metabolite of nicotine), urine NNAL (metabolite of NNK, a lung carcinogen), exhaled carbon-monoxide, cigarette consumption, and cigarette dependence. There were no significant effects on psychiatric symptoms. At the end of the 12-week treatment choice phase, those randomized to the RNC group were more likely to have quit smoking (18% RNC v 4% UNC, p=0.004).

**CONCLUSION:** Reducing nicotine content in cigarettes to very low levels reduces toxicant exposure and cigarette addiction and increases smoking cessation in smokers with mood and/or anxiety disorders, without worsening mental health.

**Trial registration:** TRN: NCT01928758, registered August 21, 2013

## Introduction

Tobacco smoking remains the leading preventable cause of premature morbidity and mortality in the U.S.(1). The U.S. Food and Drug Administration (FDA) regulates tobacco products, and in 2017, announced plans to reduce the nicotine content of cigarettes to minimally addictive levels (2).While these plans remain under consideration in USA, the government of New Zealand recently announced a plan to only allow reduced nicotine cigarettes to be sold (3). Previous studies have generally found that reduction of nicotine content in cigarettes is feasible and safe in smokers with and without comorbid psychiatric illness, and it has been estimated that this would save millions of lives (4–14). Over 25% of smokers have an affective (unipolar mood or anxiety) disorder, representing over 8 million people in the US. Affective disorder smokers (ADS) report more severe nicotine withdrawal symptoms and lower success rates when attempting cessation (15–18). It has been speculated that a policy to reduce the nicotine content in cigarettes may have the unintended consequences, particularly in vulnerable subgroups such as ADS, of exacerbating psychiatric symptoms or causing compensatory heavier smoking that could increase their exposure to toxicants in tobacco smoke (19, 20). Indeed, in one recent trial (12) among smokers with mood disorders, those randomized to very low nicotine cigarettes had significantly higher mean Beck Depression Inventory scores during the trial than those randomized to normal nicotine cigarettes. The largest randomized trial of reduced nicotine cigarettes in a non-psychiatric population (11), found that 20 weeks after randomization to very low nicotine cigarettes, 7% had quit smoking, as compared to 2% of those randomized to smoke regular nicotine cigarettes. This suggests that if ADS can tolerate reduced nicotine cigarettes (RNC) without psychiatric symptom exacerbation, reduced severity of nicotine dependence on RNC may improve smoking cessation rates in this population. This study examined the effects of reduced nicotine content cigarettes on psychiatric symptoms, severity of dependence, toxicant exposure and early abstinence rates in ADS.

## Methods

Detailed methods and design of this two-site, two-arm, double-blind, parallel group, randomized controlled 33-week trial have been previously reported (21), and are outlined in Supplementary Appendix 1, but are summarized here. This study was approved by the Penn State Hershey and Massachusetts General Hospital Institutional Review Boards.

### Study Population

Participants were 188 adults, aged 18-65, who smoked >4 cigarettes per day (CPD) for at least the past year, who reported no quit attempt in the past month, no plans to quit smoking in the next 6 months, and no current use of a smoking cessation aid, and who met lifetime diagnostic criteria for one or more anxiety or unipolar mood disorders as assessed with the Mini-International Neuropsychiatric Interview, (MINI) (22). Participants were excluded if they had a medical or psychiatric condition or behaviors that might affect participant safety or biomarker data. Recruitment occurred at Penn State Medical Center in Hershey, Pennsylvania (n=100) and Massachusetts General Hospital in Boston, Massachusetts (n=88).

### Procedures

During **Baseline I**, participants smoked their own brand of cigarettes for one week. At **Baseline II**, all participants were asked to use only SPECTRUM research cigarettes with a usual nicotine content (11.6 mg) for two weeks.

Participants who completed Baseline II and agreed to continue then entered the **Randomized Phase (III)**. They were randomized to either (1) continue to smoke the same 11.6 mg nicotine SPECTRUM research cigarettes they smoked in Baseline II for 18 additional weeks (UNC) or (2) switch to identical appearance cigarettes with progressively reduced nicotine content (RNC). Nicotine content in RNC cigarettes was reduced every 3 weeks over 18 weeks from 11.6 mg per cigarette to 0.2 mg/cigarette, remaining on this lowest level during the last 6 weeks of the randomized phase. Participants were randomized 1:1 to reduced nicotine or usual nicotine cigarettes based on a predetermined random number sequence generated by the study statistician stratified by site (Penn State and Mass. General) and by preferred flavor (regular/menthol). A Cigarette Management System was used to manage assigning randomized, blinded cigarettes to participants and to track cigarette inventory. The researchers and participants were blinded to the randomized allocation throughout the trial. In a prior pharmacokinetic study, a single UNC research cigarette provided a boost to plasma nicotine of 17.3 ng/ml, similar to an own-brand cigarette (19 ng/ml), whereas the 0.2mg nicotine RNC provided a nicotine boost of only 0.3 ng/ml (23). During the Randomized Phase, participants attended study visits and received research cigarettes every three weeks. They were provided with 150% of daily cigarette consumption reported at baseline to ensure they had an adequate supply to last until their next visit. Participants and study staff were blind to the experimental cigarette allocation throughout the randomized and treatment choice phases of the trial.

At the last visit (end of 18^th^ week) of the Randomized Phase, participants began the 12-week **Treatment Choice Phase (IV)**. Participants were given a copy of the U.S. Surgeon General Report, “How Tobacco Causes Disease” and were asked to choose one of the following options:

1. Return to smoking their usual brand of cigarettes for 12 weeks (at their own cost).
2. Continue to receive the same research cigarettes they were currently smoking (still double-blind) for a further 12 weeks (provided at no cost).
3. Quit smoking with brief counseling from the study team and the option to use oral nicotine replacement therapy (NRT [gum or lozenge]) for 11 weeks.

All participants were asked to attend two study visits in the Treatment Choice Phase at 4 and 12 weeks after the end of the randomized phase.

The sequence for all study visits, a list of all measures at these study visits and a detailed list of the nicotine content dosing schedule are provided in Supplementary Appendix 2, Figure S2 and Tables S1 and S2.

### Assessments

Biomarkers of exposure included plasma cotinine [primary outcome], exhaled carbon monoxide, urinary total NNAL and 1-hydroxypyrene. See Supplementary Appendix 2 for detailed methodology for biomarker analyses. These biomarkers and self-report of cigarette consumption were assessed at baseline visit 2 and repeated visits through 18 weeks after randomization (visit 10). Psychiatric and nicotine withdrawal symptoms were assessed with the Quick Inventory of Depressive Symptomatology (QIDS [depression measure]) (24), Overall Anxiety Severity and Impairment Scale (OASIS [anxiety measure]) (25), and Minnesota Nicotine Withdrawal Scale (26). Assessments of tobacco dependence (Fagerstrom Test of Nicotine Dependence (27) and the Penn State Cigarette Dependence Index (28)) were measured at each visit. Self-report of intention to quit smoking, and actual smoking cessation were assessed at the treatment choice phase (visits 10-12, weeks 21-33). Abstinence at visits 11 and 12 was defined as self-report of no tobacco use in the prior 7 days, validated by exhaled CO<10ppm.

### Sample size and Statistical analysis

The study was powered to detect a between group difference in plasma cotinine concentration of 58 ng/ml with at least 80% power, and a difference of 68 ng/ml with at least 90% power, based on 70 participants per group completing the randomized phase.

The statistical analysis focused on comparing the intervention and control groups, RNC vs. UNC, on (a) plasma cotinine concentration (primary outcome) at the end of the randomized phase; (b) secondary quantitative outcomes, e.g. exhaled CO,QIDS depression level and OASIS anxiety at the end of the randomized phase; (c) dropout rate during the randomized phase; (d) rate of psychiatric and other serious AEs and (e) the proportion of participants in each group choosing to try to quit and who quit smoking at the end of the treatment choice phase. Linear regression models were constructed for each quantitative outcome variable, for measures taken from the randomization visit through to the end of the randomized phase. Unadjusted regression models compared the two trial arms while controlling for the baseline value of the outcome measure. Adjusted models then evaluated the randomized treatment effect while adjusting for other baseline covariates that were selected via backward elimination using a significance level of 0.05. These models were built on data from subjects who completed the randomized phase, and were intended to focus on comparing outcomes between those who had completed 6 weeks of smoking the lowest nicotine content cigarettes in the RNC group, with those in the UNC group at that same visit (v10). A separate analysis was also conducted for participants who reported exclusive, per protocol, use of assigned research cigarettes (“compliers”), biochemically validated for those smoking the lowest nicotine content cigarettes using plasma cotinine as previously reported (29). This analysis aimed to focus on those who had not used any non-study cigarettes. Linear mixed-effect models for repeated measures were used to analyze the change over time (visits) in the main quantitative outcome measures. This analyses used data from all participants at all visits, regardless of dropout, and was intended as a sensitivity check that on the main quantitative outcomes (i.e. checking that the pattern of results on the main quantitative outcomes [cotinine, cigarettes per day, exhaled CO, QIDS, OASIS, Kessler K6, and PSS], remain the same in analyses including all participants). Chi-squared or Fishers Exact tests were was used to compare the intention to quit smoking (yes/no) at the end of the randomized phase (Visit 10), and abstinence in the treatment choice phase between the two groups. A Kaplan-Meier time-to-event analysis was used to compare the time from randomization to dropout between the two groups.

## Results

Participants were recruited between September 2015 and August 2017, and the last participant completed the study in March 2018. Figure 1 shows the participant flow diagram for the trial, including the number and reasons for dropout. A total of 143/188 (76%) of randomized participants completed the randomized phase of the trial, 73% (69/94) for the RNC group and 79% (74/94) for the UNC group. A time-to-event analysis revealed no significant difference in time-to-dropout between the two arms (log-rank p=0.41). There were no significant between-group differences in key demographic, clinical and smoking history variables (Table 1). Over 54% in both groups were taking psychiatric medications and over 57% met criteria for a current mood or anxiety disorder (the rest having past diagnoses).

**Figure 1.**
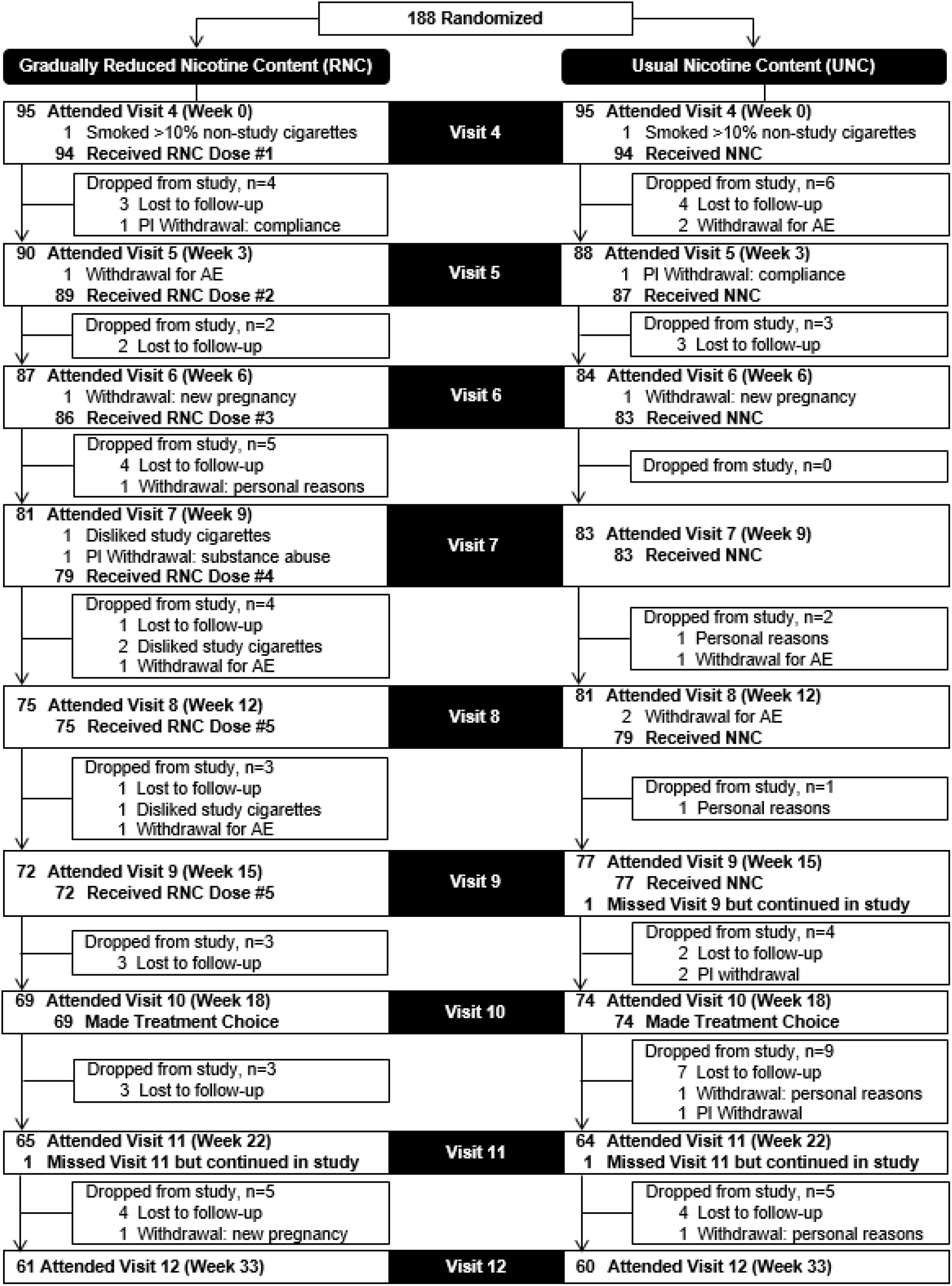
Participant Flow Diagram.

**Table 1:** Study Participant Demographic and Smoking Characteristics at Baseline Phase 1.

|  | **Reduced nicotine**<br>**content (n = 94)** | **Usual nicotine**<br>**content (n = 94)** |
| --- | --- | --- |
| Female, % (n) | 63.8 (60) | 57.4 (54) |
| Race, % (n) |  |  |
| African American | 16.0 (15) | 11.7 (11) |
| White | 74.5 (70) | 80.9 (76) |
| Other | 9.5 (9) | 7.4 (7) |
| Age (in years), mean (SD, range) | 43.3 (11.7, 21-65) | 43.1 (13.3, 19-65) |
| Bachelor's degree or higher, % (n) | 21.3 (20) | 20.2 (19) |
| Currently employed full-time, % (n) | 37.6 (35)<br>[n=93] | 38.0 (35)<br>[n=92] |
| Median number of prior attempts to quit<br>smoking (% with no prior quit attempts) | 2 (19.1) | 2 (26.6) |
| Smoke menthol cigarettes, % (n) | 40.4 (38) | 38.3 (36) |
| Number of years as daily smoker,<br>mean (SD, range) | 25.4 (12.4, 2-49) | 26.1 (13.4, 1-53) |
| Cigarettes per day, mean (SD, range) | 18.7 (10.0, 5-60) | 20.5 (10.0, 5-60) |
| Exhaled carbon monoxide (in ppm),<br>mean (SD, range) | 27.6 (17.0, 4-100) | 27.7 (16.5, 6-85) |
| Moderate or higher environmental<br>smoke exposure score, % (n) | 69.1 (65) | 74.2 (69) [n=93] |
| Fagerström Test for Cigarette<br>Dependence score, mean (SD, range) | 5.8 (2.3, 0-10) | 6.0 (2.2, 1-10) |
| CESD score, mean (SD, range) | 18.2 (8.7, 4-43)<br>[n=92] | 18.9 (7.7, 5-43)<br>[n=93] |
| Kessler K6 score, mean (SD, range) | 5.8 (5.4, 0-22)<br>[n=93] | 6.9 (5.3, 0-20) |
| Penn State Cigarette Dependence Index<br>score, mean (SD, range) | 12.9 (3.4, 5-20)<br>[n=93] | 13.4 (3.4, 6-20) |
| Lifetime suicidality, % (n) | 34.0 (32) | 29.8 (28) |
| Number of MINI mood/anxiety disorder<br>diagnoses, % (n) |  |  |
| One current diagnosis | 30.9 (29) | 33.0 (31) |
| Two or more current diagnoses | 26.6 (25) | 35.1 (33) |
| Past diagnosis/-es only | 42.5 (40) | 31.9 (30) |
| Current/[Past] mood disorder, % | 24.5 / [67.0] | 27.7 / [63.8] |
| Current/[Past] anxiety disorder, % | 47.9 / [48.9] | 63.8 / [45.7] |
| Currently prescribed at least one<br>medication for psychiatric reasons, % (n) | 54.3 (51) | 55.3 (52) |

Baseline and end of treatment values for the main outcomes are given in Table 2.

**Table 2.**
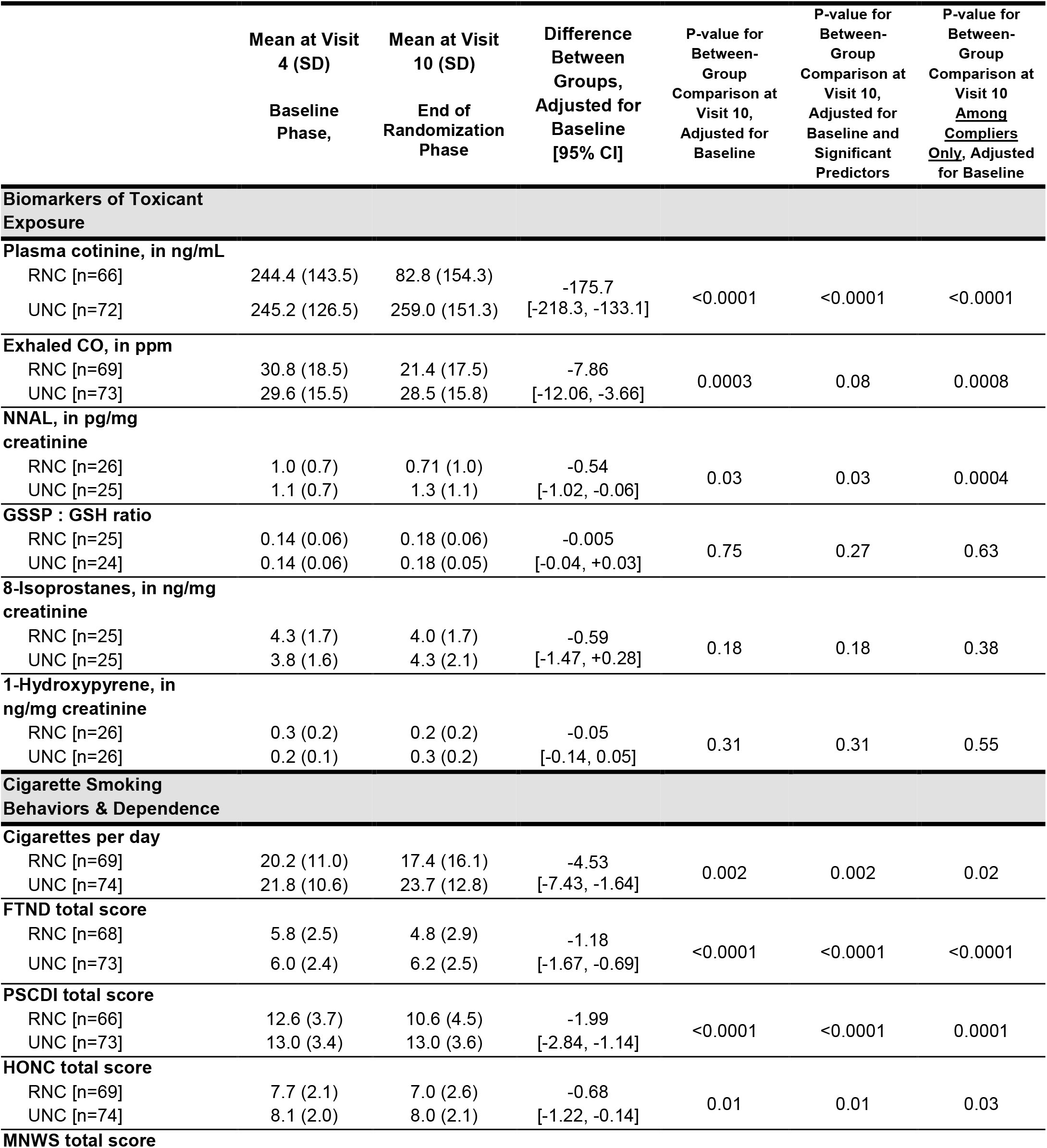
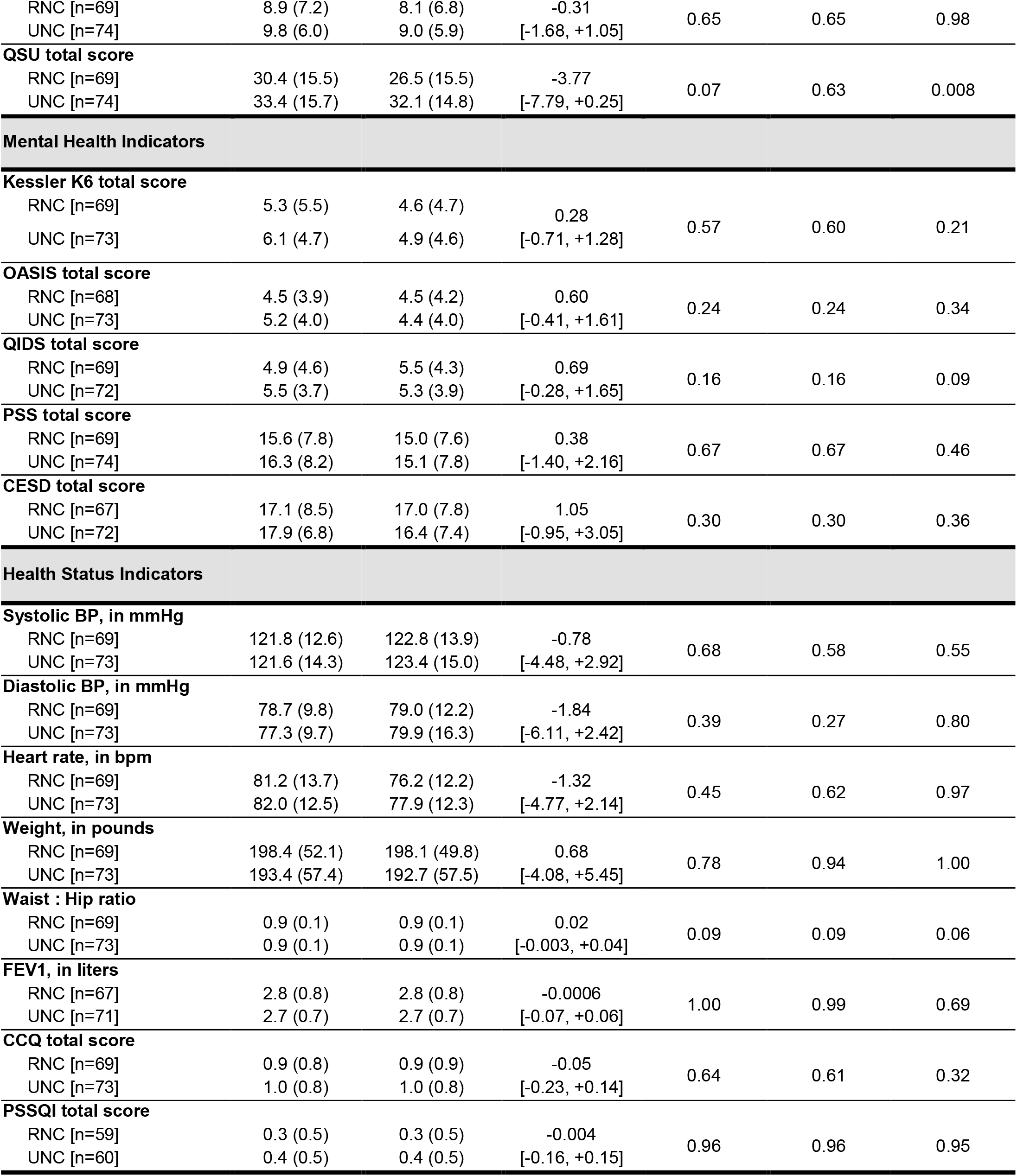
Means at baseline (visit 4) and end of randomized phase (visit 10) for each group and results of statistical comparisons.

Multivariable linear regression models show indicators of exposure, plasma cotinine, exhaled CO and NNAL concentration, and measures of nicotine dependence, CPD, FTND, PSCDI, were significantly lower at the end of the randomized phase in the RNC group as compared to the UNC group. Assessments of psychiatric and nicotine withdrawal symptoms, CES-D, QIDS, OASIS, PSS and MNWS, showed no significant between group differences. There were also no significant effects of treatment group on health indicators or biomarkers of oxidative stress (glutathione, 8-isoprostanes). Linear mixed-effect models, incorporating data from all randomized participants and visits showed an identical pattern of results (full results shown in Figures S3-S54 in Supplementary Appendix 2) with significant visit (time) by group interactions for cotinine, CPD, and CO but not for any of the mental health indicators.

Figure 2 shows the primary outcome data for plasma cotinine, exhaled CO, daily cigarette consumption and the FTCD measure of nicotine dependence throughout the study.

**Figure 2A-D.**
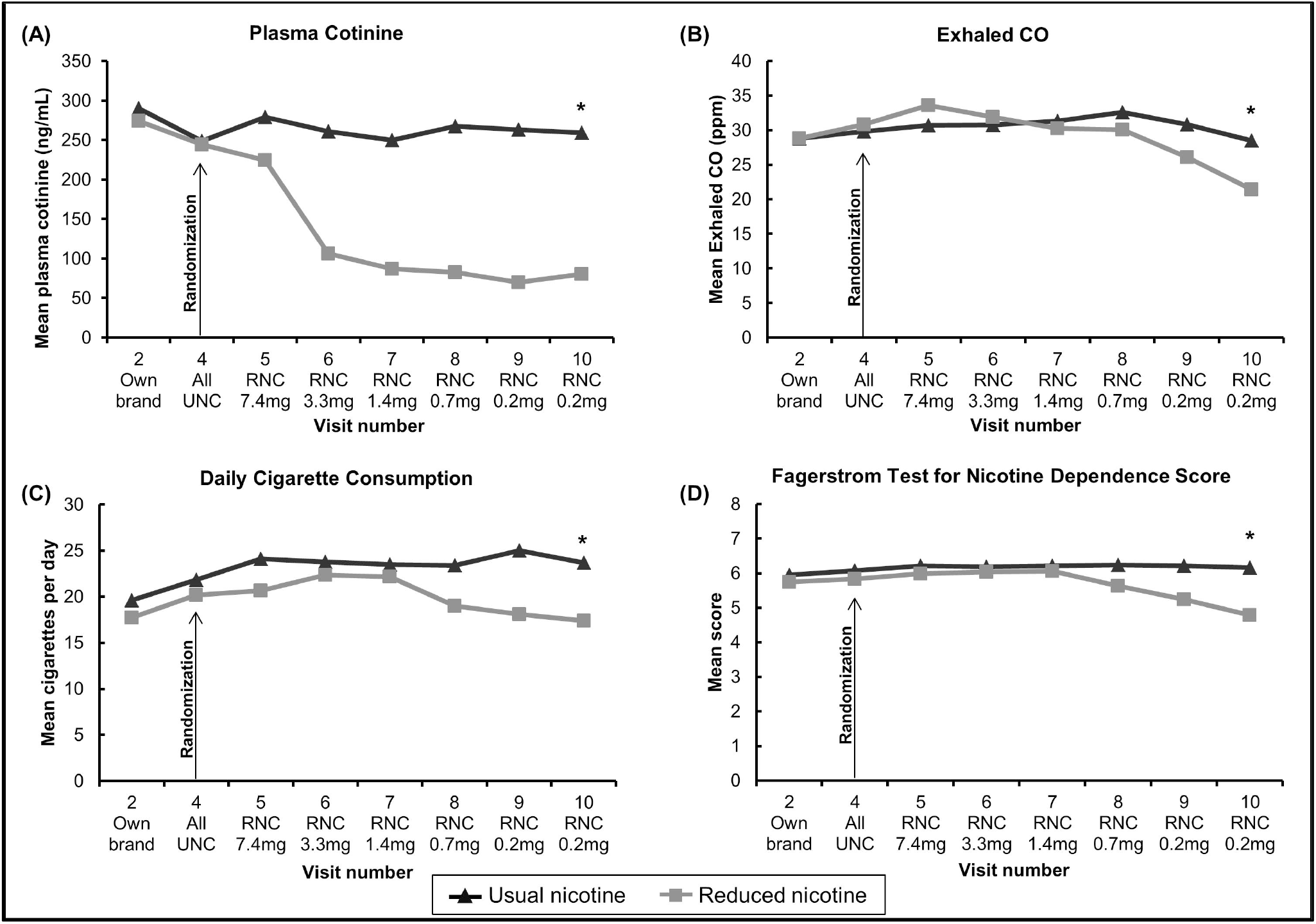
Changes in plasma cotinine, exhaled CO, daily cigarette consumption, and FTND nicotine dependence score among completers (n=143) smoking either Usual Nicotine Content (n=74) cigarettes or Reduced Nicotine Content (n=69) cigarettes. *indicates significant between group difference at Visit 10, controlling for Visit 4 baseline

Figure 3 shows the outcome data for mental health measures (OASIS, QIDS, PSS) and the carcinogen exposure biomarker NNAL throughout the study. The other mental health and general health indicators showed similar patterns with no significant differences between groups.

**Figure 3A-D.**
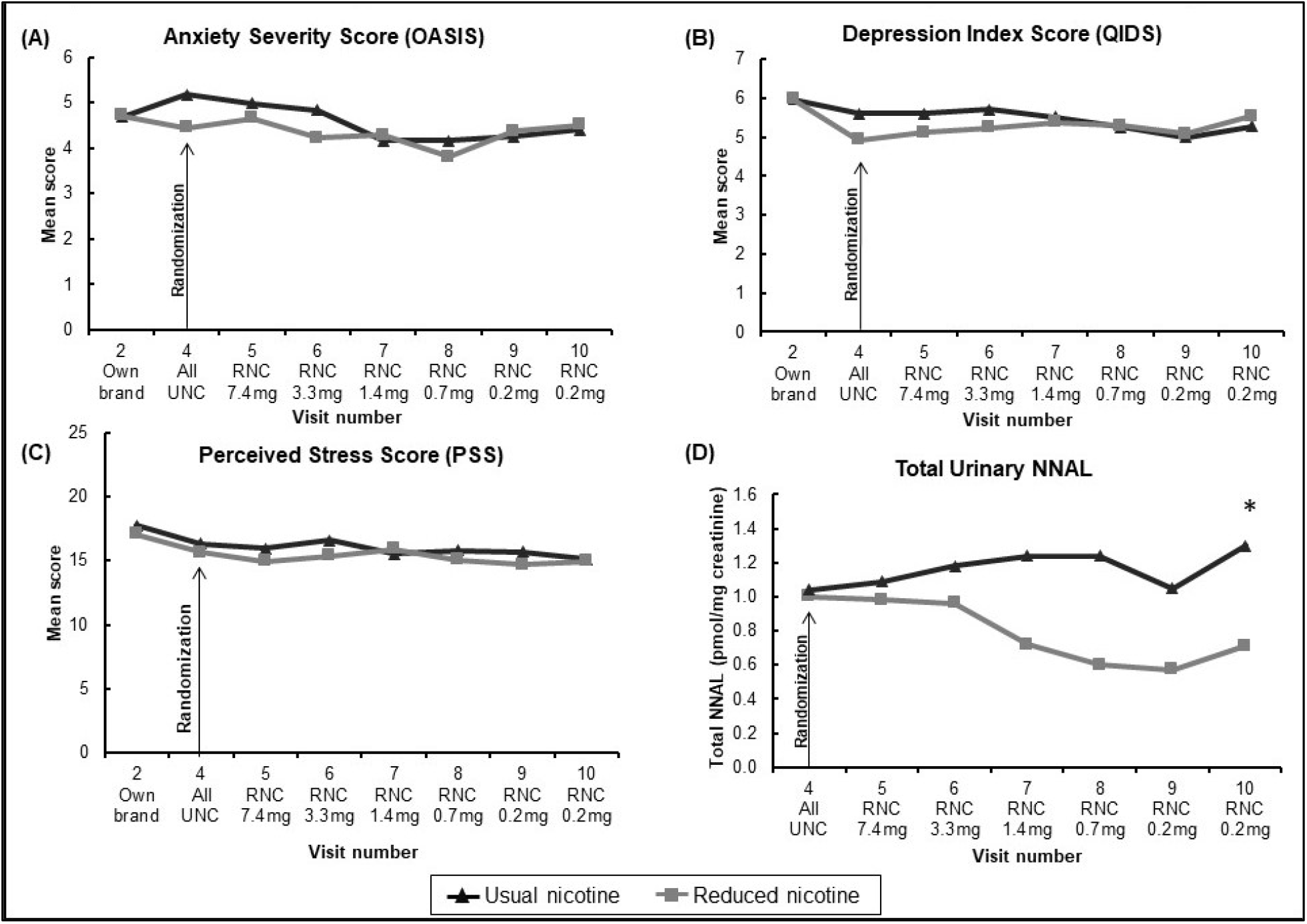
Changes in OASIS, QIDS, PSS, and NNAL^#^ among completers (n=143) smoking either Usual Nicotine Content (n=74) cigarettes or Reduced Nicotine Content (n=69) cigarettes. *indicates significant between group difference at Visit 10, controlling for Visit 4 baseline # urinary NNAL measured in randomly selected n=26 on RNCs and n-=25 on UNCs.

### Protocol adherence

Adherence with the study protocol to smoke only the assigned research cigarettes during the trial was imperfect for both groups, with 62 (84%) participants in the UNC and 41 (59%) in the RNC group meeting self-report and biochemical criteria for strict adherence, defined as non-use of non-research cigarettes (29). The overall pattern of results comparing the subgroups in each arm with strict adherence was very similar to the results reported above for study completers. The Supplementary Appendix 2 provides figures for each outcome showing the pattern of change in each group among (a) all completers to visit 10 (b) all completers who were compliant with their assigned research cigarettes at visit 10 (compliers) and (c) for main outcome measures [per protocol] the pattern of results for all participants attending each visit (n which varied by visit).

### Adverse Events

A total of 144/188 participants (76.6%) reported at least one adverse event (AE) during the randomized phase of the trial, with very similar frequencies in the intervention groups: 75.5% in the RNC group and 77.7% of the UNC group. Two-thirds (215/327) of the AEs were considered “mild”. Thirteen serious adverse events (SAEs) occurred in 12 participants during the randomized phase; 4 among participants randomized to RNC cigarettes and 9 among those randomized to UNC cigarettes. Three of these SAEs were psychiatric, 1 in the RNC group and two in the UNC group. Eight of these participants were withdrawn from the trial (3 on RNCs, 5 on UNCs) due to their SAE. Details of AEs are in the Supplementary Appendix 2 (Tables S4-S10).

### Increases in use of psychiatric medications

As shown in Table 1, more than half of each group was using a psychiatric medication at enrollment. Ten participants (10.6%) in the UNC group and 12 (12.8%) in the RNC group increased their dose or started a new psychiatric medication during the trial. Of participants who were not taking a psychiatric medication at randomization, 4 participants in the UNC and 5 participants in the RNC group started taking a psychiatric medication during the randomized phase.

### Treatment Choice and Smoking Cessation

143 participants attended the last randomized phase visit and entered the treatment choice phase of the trial. 33/69 (47.8%) of those on RNCs and 25/74 (33.8%) on UNC cigarettes chose to try to quit smoking; 25/69 (36.2%) of those on RNCs and 46/74 (62.2%) on UNCs chose to continue smoking study cigarettes, while 11/69 (15.9%) on RNC and 3/74 (4.1%) on UNC chose to return to smoking their own brand cigarettes. The association between the treatment choice and study arms was significant, (Chi-Squared, p=0.003).

At the end of the treatment choice phase (visit 12), 17/94 (18.1%) of the RNC group and 4/94 (4.3%) in the UNC group met study criteria for abstinence, defined as self-report of no tobacco use in the previous 7 days and exhaled CO <10ppm, considering dropouts to be smokers, Fisher’s exact test, p=0.004. The mean CO of those abstinent at visit 12 was <4ppm for both groups and all but two (one of each group) were also abstinent 8 weeks earlier (visit 11).

## Discussion

This study found that when ADS switch to cigarettes with gradually reduced nicotine content, they have progressively lower plasma cotinine, and once they are smoking cigarettes with very low nicotine content they smoke fewer cigarettes per day, have a lower exhaled CO and report being less addicted to their cigarettes than smokers randomly assigned to continue smoking usual nicotine cigarettes. We found no evidence that using reduced nicotine cigarettes, versus UNCs, was associated with worsening general health or mental health problems or adverse events. When offered a choice to quit smoking, more of those randomized to RNC cigarettes actually succeeded in quitting smoking over 12 weeks, despite the fact that only slightly more of the RNC group (n=33) than the UNC group (n=25) chose to try to quit. To our knowledge, this is the first randomized trial of reduced nicotine cigarettes in smokers with affective disorders to find that randomization to RNC cigarettes was associated with significantly increased rates of biochemically-validated smoking cessation. The higher biochemically validated quit rate in the RNC group is consistent with the lower measured dependence in that group towards the end of the randomized phase of the trial.

Limitations of the study include the facts that 26% of the participants did not complete the randomized phase and imperfect adherence to the protocol for exclusive use of research cigarettes in those who did complete the trial. However, the rate of dropout was similar in the two groups and was anticipated in the trial protocol, which aimed to conduct the main analyses on approximately 70 completers in each group. The pattern of results was virtually identical for “compliers-only” analyses as for analyses of all completers. While the trial included smokers with a lifetime history of mood and/or anxiety disorders and did not require a current mood or anxiety disorder for enrollment, and those who were recently suicidal or had recently received inpatient psychiatric treatment were excluded, almost a third of the sample had a history of attempted suicide, suggesting that the sample was at high risk of worsening mental health. However, this study did not find that switching to RNC cigarettes worsens mental health.

The present study is consistent with others (9, 12, 13) in being broadly reassuring about the effects of switching to very low nicotine cigarettes on mental health outcomes in those with unipolar mood and anxiety disorders, and adds the findings of reduced toxicant exposure and increased probability of successful smoking cessation when treatment is offered. A recent trial (11) that excluded smokers with serious psychiatric illness demonstrated that abrupt nicotine reduction in cigarettes is feasible. Future research should examine the effects of abrupt nicotine reduction in cigarettes on smokers with psychiatric conditions, and also assess the effects of availability of other non-combusted nicotine sources (e.g. electronic cigarettes or oral nicotine products) on the effects of abrupt nicotine reduction in cigarettes.

## Conclusion

Lowering the permissible nicotine content in cigarettes to very low levels over 15 weeks reduces toxicant exposure and increases smoking cessation without worsening mental health among smokers with mood or anxiety disorders.

## Data Availability

With publication, requests for de-identified individual participant data and/or study documents (data dictionary, protocol, statistical analysis plan, measures/manuals/informed consent documentation) will be considered. The requestor must submit a 1-page abstract of their proposed research, including purpose, analytical plan, and dissemination plans. The Leadership Committee of the Penn State Center for Research on Tobacco and Health will review the abstract and decide based on the individual merits. Review criteria and prioritization of projects include potential of the proposed work to advance public health, qualifications of the applicant, the potential for publication, the potential for future funding, and enhancing the scientific, geographic, and demographic diversity of the research portfolio. Following abstract approval, requestors must receive institutional ethics approval or confirmation of exempt status for the proposed research. An executed data use agreement must be completed prior to data distribution. Contact is through Dr. Jonathan Foulds.

## Abbreviations

NNAL: 4-(methylnitrosamino)-1-(3-pyridyl)-1-butanol
NNK: 4-(methylnitrosamino)-1-(3-pyridyl)-1-butanone
CO: carbon monoxide
DSM-5: Diagnostic and Statistical Manual of Mental Disorders, 5th Edition

## Declarations

### Ethics approval and consent to participate

#### Funding

Research reported in this publication is supported by the National Institute on Drug Abuse of the National Institutes of Health under Award Number P50DA036107 and the Center for Tobacco Products of the U.S. Food and Drug Administration. The content is solely the responsibility of the authors and does not necessarily represent the official views of the National Institutes of Health or the Food and Drug Administration. Participants at the Penn State University site are seen at the Clinical Research Center supported by the Penn State Clinical & Translational Research Institute, Pennsylvania State University CTSA, NIH/NCATS Grant Number UL1 TR000127.

#### Authors’ Contributions

JF, AEE and JEM conceived the original idea for the trial, sought and obtained funding. All authors read and approved the final manuscript.

## Acknowledgements

We gratefully acknowledge the members of our Data and Safety Monitoring Board (Drs. John Hughes [Chair], Michael Steinberg, and David Mauger) and external consultants on our TCORS research (Drs Dorothy Hatsukami and Neal Benowitz). We are also grateful for assistance from Dr Abid Kazi, Alyse Fazzi, and Richard Sargent.

## Authors’ information

^1^Penn State Center for Research on Tobacco and Health, Department of Public Health Sciences, Pennsylvania State University College of Medicine, MC CH69, 500 University Drive, P.O. Box 850, Hershey, PA, 17033, USA. ^2^Department of Public Health Sciences, Pennsylvania State University College of Medicine, Hershey, PA, USA. ^3^Department of Psychiatry, Pennsylvania State University College of Medicine, Hershey, PA, USA. ^4^Massachusetts General Hospital Center for Addiction Medicine, Boston, MA, USA. ^5^Harvard Medical School, Boston, MA, USA

