## Supplement 1 for "The effects of reduced nicotine content cigarettes in smokers with mood or anxiety disorders: a double-blind randomized trial"

PENNSTATE
1855

### Protocol for
### Human Subject Research
### with Use of Test Article(s)

**Protocol Title:** Reduced Nicotine Cigarettes in Smokers with Mood and Anxiety Disorders

**Principal Investigator:**
Jonathan Foulds, PhD
Public Health Sciences
717-531-3504


**Version Date: 1-10-19**

**Clinicaltrials.gov Registration #:** NCT01928758

#### Table of Contents

1.0 Objectives
2.0 Background
3.0 Inclusion and Exclusion Criteria
4.0 Recruitment Methods
5.0 Consent Process and Documentation
6.0 HIPAA Research Authorization and/or Waiver or Alteration of Authorization
7.0 Study Design and Procedures
8.0 Data and Specimen Banking For Future Undetermined Research
9.0 Statistical Plan
10.0 Confidentiality, Privacy and Data Management
11.0 Data and Safety Monitoring Plan
12.0 Risks
13.0 Potential Benefits to Subjects and Others
14.0 Sharing Results with Subjects
15.0 Economic Burden to Subjects
16.0 Number of Subjects
17.0 Resources Available
18.0 Other Approvals
19.0 Subject Stipend (Compensation) and/or Travel Reimbursements
20.0 Multi-Site Research
21.0 Adverse Event Reporting
22.0 Study Monitoring, Auditing and Inspecting
23.0 References
24.0 Appendix

### 1.0 Objectives

#### 1.1 Study Objectives

Tobacco smoking is the leading preventable cause of premature morbidity and mortality in the United States [1], and cessation provides immediate and sustained improvement in health and quality of life [2]. Nicotine is the primary addictive substance in cigarettes, and cigarettes smoked in the United States typically contain an average of 11 mg of nicotine [3-5]. It should be noted that the nicotine content of a cigarette is a different concept than the “nicotine yield” as measured by a smoking machine, and which was previously printed on cigarette packs. It is now known that the main factor affecting the machine-smoked nicotine yield in current commercially available cigarettes is the amount of ventilation in the filter. For example the brands formerly known as “Marlboro Lights” actually contained almost an identical amount of nicotine as regular Marlboro Reds (11mg), but had more ventilation holes in the filter. It is now known that smokers smoke highly ventilated (low nicotine yield) cigarettes differently from regular cigarettes, and end up absorbing almost as much nicotine and toxicants by increasing puff frequency and volume. It is partly for this reason that the terms “Light” and “Mild” are no longer allowed. The Family Smoking Prevention and Tobacco Control Act [6] gave the FDA jurisdiction to regulate tobacco products, including the nicotine content of cigarettes. Progressive reduction of the nicotine content of cigarettes to very low levels is a potential way to reduce the addictiveness of cigarettes, and may help established smokers to quit smoking [7, 8]. Preliminary studies have found that progressive reduction of nicotine content of cigarettes is feasible and safe in smokers without comorbid psychiatric illness [8-11]. However, it is not known whether progressive nicotine reduction is feasible and safe in the large subgroup of smokers with comorbid psychiatric illness. Smokers with psychiatric illnesses purchase over 40% of cigarettes sold in the US [12], have a higher prevalence of smoking, greater severity of nicotine dependence and lower cessation rates than smokers without comorbid psychiatric illness [12, 13]. Smokers with a prior mood or anxiety disorder report more severe nicotine withdrawal symptoms during a cessation attempt [14, 15]. This suggests the possibility of more difficulty switching to very low nicotine content cigarettes, and potentially more compensatory smoking of lower nicotine content cigarettes that could increase exposure to other toxins in tobacco smoke [16, 17]. On the other hand, if smokers with mood and anxiety disorders can safely transition to significantly reduced nicotine content cigarettes, it is plausible that further progression to smoking cessation may be more achievable from a lower level of nicotine dependence. Prior to implementation of progressive nicotine reduction on a national level, it is important to establish the feasibility and safety of this approach in this subgroup of smokers.

**The overall aim of this proposed study is to evaluate the effect of progressive nicotine reduction in cigarettes on smoking behavior, toxicant exposure and psychiatric symptoms in smokers with comorbid mood and/or anxiety disorders.** To do so, we will randomly assign adult smokers with a history of unipolar mood and/or anxiety disorder to smoke research cigarettes that will contain either a) Usual Nicotine Content (UNC): nicotine content (11.6mg) similar to popular brands of cigarettes; or b) Reduced Nicotine Content (RNC): the nicotine content per cigarette is progressively reduced from approximately 11.6 mg to 0.2 mg over 5 months. All subjects will participate in lead-in periods prior to randomization to assess normal smoking behavior and to establish ability to tolerate research cigarettes prior to randomization. It is our hypothesis that nicotine intake, as measured by plasma cotinine concentration, will decline as a function of cigarette nicotine content in the RNC group. Further, it is our hypothesis that by gradually reducing the nicotine content of the cigarettes in three week steps, there will not be significant increases in biomarkers of tobacco smoke exposure, severity of nicotine withdrawal symptoms, mood and anxiety symptomatology or protocol non-adherence over time in the experimental group (RNC) as compared with the UNC control group.

***Aims:***

**1.** To assess the effect of switching to gradually reduced nicotine content cigarettes on product use patterns and biomarkers of exposure in smokers with mood and/or anxiety disorders.

***Hypothesis 1.a.:*** Smokers assigned to the RNC group will have lower plasma cotinine concentrations during the last 3 weeks of the randomized phase of the study than those assigned to the UNC group.

***Hypothesis 1.b.:*** There will be no significant increase in key markers of tobacco use (e.g. cigarette consumption), biomarkers of tobacco smoke exposure (e.g. NNAL, CO), or health effects (e.g. blood pressure) in those assigned to the RNC group vs. those assigned to UNC.

**2.** To assess the effect of switching to gradually reduced nicotine content cigarettes on psychiatric and nicotine withdrawal symptoms in smokers with unipolar mood and / or anxiety disorders.

***Hypothesis 2:*** There will be no significant increase in ratings of psychiatric or nicotine withdrawal symptoms in those assigned to the RNC group as compared to those assigned to UNC.

***3.*** To assess the effect of switching to gradually reduced nicotine content cigarettes on self-perception of tobacco dependence, self-report of intention to quit smoking, and actual smoking cessation.

***Hypothesis 3:*** Among smokers who complete the randomized nicotine reduction phase, smokers assigned to the RNC group will have lower perceived dependence, be more likely to report intention to quit smoking, and be more likely to make a smoking cessation attempt and achieve cigarette abstinence at visit 12..

##### 1.2 Primary Study Endpoints

Plasma cotinine concentrations during the last 3 weeks of the randomized nicotine reduction phase (visit 10).

##### 1.3 Secondary Study Endpoints

Biomarkers of tobacco smoke exposure (e.g. NNAL, CO), cigarette consumption, health effects (e.g. blood pressure) and mental health measures (e.g. nicotine withdrawal, perceived stress, QIDS, Kessler K6).

The proportion of each group making a choice to try to quit smoking will be an endpoint, as will the proportion succeeding in quitting successfully during the choice phase. Intent to treat (ITT) abstinence (based on the assumption that a loss-to-follow-up subject resumes smoking) and completers' abstinence (based on the assumption that the probability of loss to follow up is independent of smoking status) will be analyzed. Abstinence will be biochemically verified (CO < 10ppm).

#### 2.0 Background

##### 2.1 Scientific Background and Gaps

**Reducing nicotine content in cigarettes:** This approach has the potential to significantly reduce the public health harm from tobacco dependence. However, the impact of reduced nicotine cigarettes on smoking behavior, toxicant exposure and cessation needs to be assessed empirically prior to implementation of a national nicotine reduction policy [7]. The primary concern about a progressive nicotine reduction strategy is that it may have the unintended consequence of compensatory increase in tobacco smoke exposure as nicotine content in cigarettes is reduced (if smokers smoke more cigarettes per day, inhale more smoke per cigarette, or both) [16]. It has been postulated that compensatory smoking of reduced nicotine cigarettes may change the way cigarettes are smoked in a manner that results in increased toxicant exposure, even if compensation (with respect to nicotine extraction) is incomplete [17]. This could occur, for example, by changes in the puffing patterns causing changes in the temperatures achieved in the rod and tip of the cigarette and consequent changes in the chemicals inhaled by the smoker.

##### 2.2 Previous Data

Early studies of the behavioral effects of a progressive nicotine reduction strategy have been encouraging. A series of studies by Benowitz and colleagues [9-11] examined the effects of a progressive nicotine reduction strategy in which smokers switched to cigarettes with progressively lower nicotine content (or yield) over 4 weeks or 6 months. In their initial study of 20 smokers, gradual reduction of nicotine content was feasible and did not increase apparent exposure to tobacco smoke toxins [9]. In a follow up study [10], 135 otherwise healthy smokers were randomly assigned to one of two groups: (a) an experimental group that smoked their usual brand followed by research cigarettes with progressively reduced nicotine content (RNC over five, month-long steps), or (b) a control group that continued smoking their own brand of cigarettes for the entire study period. In the smokers assigned to RNC cigarettes, nicotine intake, as assessed by plasma cotinine concentration, declined progressively as the nicotine content of the cigarettes was reduced, while toxin exposure remained stable or, in the case of the carcinogen, NNAL, was reduced. The titration was well tolerated, but smokers assigned to the RNC group reported decreased "vigor" scores and increased "confusion" scores on the Profile of Mood States (POMS) during the time they were smoking 2mg and 1mg nicotine content cigarettes (compared with baseline), and smokers assigned to the RNC group who were adherent to study procedures also had a 2kg increase in body weight during the study period. These effects are consistent with nicotine withdrawal effects [18]. Importantly, significantly more of the RNC group (51%) than the control group (14%), were interested in quitting smoking by the end of the tapering period. These studies [8-10, 19, 20] demonstrate that a progressive nicotine reduction strategy is feasible, results in reduced nicotine exposure, increases desire to quit smoking and does not result in increased exposure to other toxicants.

These studies have been well conducted, clear in their conclusions and represent a critical first step in the process of empirically evaluating the effects, intended or unintended, of a policy of progressive nicotine reduction in cigarettes. However, there are some issues regarding generalizability of the results that must be evaluated prior to implementation of such a policy. A large proportion (>40%) of cigarettes sold in the United States are sold to someone with a current psychiatric illness, and smoking prevalence is demonstrably higher among those groups. However, enrollment in the studies conducted to date has focused on relatively well-educated, otherwise healthy smokers. In the most recent study[10], 35% of those screened were excluded for current drug or alcohol dependence and 20% for other health issues, and those enrolled had an average of 15 years education. Further, there was differential drop out for those with more severe nicotine dependence as evidenced by higher baseline Fagerstrom Test for Nicotine Dependence (FTND) scores.

Studies of potentially modified risk tobacco products to date have measured a range of biomarkers, but have tended to focus on certain reliable key measures of nicotine exposure (e.g. cotinine), smoke exposure (e.g. exhaled carbon monoxide), and carcinogen exposure (e.g. NNAL). These markers have the advantages of being well validated, are relatively specific to tobacco smoking, and change relatively quickly in response to changes in exposure(39). However, tobacco products are a major cause of oxidative stress and evaluation of the potential risks of different tobacco products should include measurement of biomarkers of oxidative stress. Tobacco smoke is an abundant source of free radicals, containing over $10^{17}$ reactive oxygen and nitrogen species (ROS/RNS) per puff and considerable evidence indicates that these agents play fundamental roles in the development of many of the major smoking-caused diseases including cancer, COPD and heart disease (1). Our team has pioneered the use of certain measures of oxidative stress, including demonstrating differences between levels in cigarette smokers and non-smokers (40), and demonstrating relationships between these measures and tobacco-related cancer (41). We therefore propose to extend that line of research, by assessing three measures of oxidative stress at baseline and during the randomized portion of our proposed trial. We believe this will be the first time that valid biomarkers of oxidative stress will be assessed in a randomized trial of RNC cigarettes.

##### 2.3 Study Rationale

Approximately one-third of adult smokers in the U.S. had a mental disorder in the past 12 months, compared with less than one in five non-smoking adults [21]. Twenty three percent of adult smokers have a comorbid anxiety disorder and 13% have a comorbid mood disorder [21]. Among nicotine dependent smokers, 22% have a mood disorder and 23% have an anxiety disorder (compared with 7% and 10% of nonsmokers) [22]. Smokers with a comorbid psychiatric illness may be more vulnerable to unintended effects of a progressive nicotine reduction policy at the national level. These smokers, by the nature of their comorbid psychiatric illness, experience chronic high levels of psychological distress, and may be more likely to exhibit compensatory smoking behavior in order to maintain systemic nicotine concentrations. Smokers with mental illnesses report wanting to quit smoking, trying to quit smoking, and use of smoking cessation aids at similar rates to people without mental health problems, but may be less likely to succeed when they attempt to quit smoking [21, 23].

The reasons for the well-established, high prevalence of comorbid mood and anxiety disorders in those with nicotine dependence are complex and not fully understood. One of the most basic reasons may be that nicotine withdrawal symptoms such as anxiety and depression may be harder to cope with for those already suffering related disorders. Many studies have found that smokers with anxiety disorders report elevated nicotine withdrawal symptoms during early abstinence, compared to those without [15, 24-26]. Potential mechanisms have been identified but remain uncertain. For example, smokers with a prior major depressive episode have greater smoking-induced brain dopamine release than those without such a history [27]. Non-nicotine constituents in cigarette smoke may also be involved. Monoamine oxidase A (MAO-A) is a brain enzyme that modulates mood because it metabolizes monoamines, including dopamine. During tobacco abstinence, heavy smokers experience an increase in brain MAO-A. This effect, which covaries with severity of depression, suggests one possible biological mechanism for worsening of depressed mood during withdrawal from cigarette smoking [28].

For the reasons described above, smokers with mood and anxiety disorders may be particularly vulnerable to unintended effects of a policy of progressive nicotine reduction in cigarettes. It is not known if smokers with these conditions are more likely to “oversmoke” reduced nicotine cigarettes and experience increased toxin exposure as a result. It is also possible that smokers with mood and anxiety disorders may experience some exacerbation in their psychiatric symptoms as they gradually withdraw from nicotine under a nicotine reduction policy. On the other hand, if smokers with mood and anxiety disorders can safely transition to significantly reduced nicotine content cigarettes, it is plausible that further progression to smoking cessation may be more achievable from a lower level of nicotine dependence. We therefore propose to examine the effects of switching to reduced nicotine content cigarettes in smokers with current or past anxiety and mood disorders. We see this as a critical test of the feasibility of the RNC regulatory strategy.

#### 3.0 Inclusion and Exclusion Criteria

##### 3.1 Inclusion Criteria

1. Aged 18 - 65
2. Smoke >4 cigarettes/day for at least the past 12 months
3. No quit attempt in prior month and not planning to quit smoking within next 6 months (to ensure stability of smoking)
4. Plan to live in local area for next 8 months
5. Meet DSM-V lifetime diagnostic criteria for a current or lifetime Anxiety Disorder (Panic disorder, agoraphobia, social phobia, obsessive compulsive disorder, post-traumatic stress disorder, generalized anxiety disorder) or a unipolar Mood Disorder (e.g. Dysthymia, Major Depression,) based on MINI (Mini International Neuropsychiatric Interview) structured interview
6. Women not pregnant or nursing and taking steps to avoid pregnancy
7. Able to understand and consent to study procedures
8. Read and write in English
9. No members of the household currently involved in any trial related to reduced nicotine cigarettes
10. Participants who become subject to correctional supervision through probation, parole, home confinement, electronic monitoring, work release or other monitored, non-custodial supervision after signing the consent form.

    Notes related to prisoner participation: The prisoner participant must be permitted by their probation/parole officer, court or other authority to go to the research office visits, and any monitoring devices need to be programmed to reflect that additional time/place of travel. These issues will be resolved by the prisoner and their supervising authority on a case-by-case basis.

    Rationale to include prisoners: Potential benefit of the subject (intent and reasonable probability of improving the health or well-being of the subject).

#### 3.2 Exclusion Criteria

1. Unstable or significant medical conditions and conditions such as elevated blood pressure (Systolic >160 mm Hg at baseline), COPD or kidney failure and those that are likely to affect biomarker data
2. Use of non-cigarette nicotine delivery product in the prior week (including cigars, pipes, chew, snus, hookah, electronic cigarette and marijuana)
3. Currently reducing or planning to reduce cigarette consumption in next month.
4. Use of a smoking cessation medicine in prior month (any nicotine replacement, varenicline, bupropion)
5. Uncontrolled serious psychotic illness (includes diagnosis of schizophrenia, bipolar disorder, eating disorder, and dementia) or inpatient treatment for a mental health condition in the past 6 months
6. More than weekly use in the past 3 months of illegal drugs or prescription drugs that are not being used for medically prescribed purposes or inpatient treatment for these in the past 6 months
7. Alcohol use that would hinder the participant's ability to participate.
8. Current suicide risk on clinical assessment (above "low risk" score on MINI diagnostic interview)
9. Unwillingness to provide blood samples or history of repeatedly fainting during blood draws
10. Unwilling to remain on one flavor of cigarette (regular or menthol) for the duration of the trial or smokes hand-rolled cigarettes
11. Any other condition or situation that would, in the investigator's opinion, make it unlikely that the participant could comply with the study protocol.
12. Those who are prisoners at the time of enrollment.
13. Those who become custodial inmates at any correctional facility after enrollment in the research.

#### 3.3 Early Withdrawal of Subjects

##### 3.3.1 Criteria for removal from study

**Participant withdrawal prior to randomization (Baseline I and II)**

Baseline Phase I and II have been designed to identify participants who are not able or willing to comply with the full study protocol after randomization. During Baseline I, participants will smoke their own usual brand of cigarettes for 1 week. At Baseline II, all participants will be asked to smoke research cigarettes with a normal amount of nicotine (around 11mg). Participants who are removed prior randomization will be replaced until a total of 200 participants have been randomized (100 at the Hershey site and 100 at Mass. General Hospital).

At any time prior to randomization participants will be withdrawn from the study if:

1. **They report using non-cigarette nicotine products at more than one visit.** This includes any number of cigars, pipes, snuff, chew, hookah, electronic cigarette, marijuana, or any other illegal smoked substance.

During Baseline II only, participants will be withdrawn from the study if:

1. **Participant's total cigarette consumption includes more than 10% of non-research cigarettes** in the 6 days prior to visit 4 only (Average cigarettes from day 15-20, e.g. 4 or more out of 30 cigarettes in 6 days for a 5 cpd smoker; 18 or more out of 180 cigarettes in 6 days for a 30 cpd smoker).
2. **Participant has reduced their cigarette consumption by more than 50% from baseline** (when cigarettes per day are averaged over days 15-20).
3. **Significant baseline smoking rate increase:** A participant will be withdrawn from the study if they meet **BOTH** of the following criteria:

a. The average cigarette per day (CPD) increase by more than 100% from the average CPD at the Baseline I assessment (visit 1) when calculated over the previous 6 days.
b. The average of two consecutive expired breath carbon monoxide measurements increase according to the following:
    i. CO is greater than 50 ppm if CO at assessment visit 1 is <20 ppm.
    ii. CO is greater than 60 ppm if CO at assessment visit 1 is 20 – 34 ppm.
    iii. CO is greater than 70 ppm if CO at assessment visit 1 is 35 – 49 ppm.
    iv. CO is greater than 80 ppm if CO at assessment visit 1 is 50 – 60 ppm.
    v. CO is greater than 90 ppm if CO at assessment visit 1 is 61 – 70 ppm.

Participants may be discontinued by the investigator at any point during the study for any of the following reasons:

- Pregnancy (confirmed by urinary pregnancy test every 4-8 weeks. See Time and Events table).
- High risk of suicide based on MINI Suicide module or has made a suicide attempt
- Elevated blood pressure at any visit (Systolic >160 mmHg).
- Cardiovascular (CVD) event**:** including MI (heart attack), PTCA (angioplasty/stenting), bypass surgery, stroke, peripheral vascular disease,, arrhythmias and new valvular disease (e.g., mitral or aortic regurgitation).
- DVT/PE (deep vein thrombosis/pulmonary embolism)
- Participants may choose to remove themselves from the study by informing the research team at any point during the study.
- Significant baseline smoking rate increase starting at visit 4 as described above and ending when participant is no longer receiving research cigarettes. Either one, or both criteria combined:
    - The average CPD increases by more than 100% from the average CPD at the baseline assessment visit.
    - Expired breath carbon monoxide increases, as described above
- Worsening substance use in which the participant is behaving inappropriately at visits or demonstrates an inability to continue with the study.
- Any hospitalization in which participation in the study could be detrimental to the recovery process. This could include recovery from a major surgery, worsening of psychiatric symptoms, etc.)
- For any missed visit where the participant would have received new research cigarettes, participants will be reviewed and considered for withdrawal on a case by case basis.
- Any situation where participant is not able to smoke research cigarettes for a period of more than 2 consecutive weeks
- Participant doesn't comply with the protocol, behaves in an inappropriate or threatening manner, admits to lying about eligibility criteria, or is participating in other smoking research studies that could affect the primary outcome measures.

##### 3.3.2 Follow-up for withdrawn subjects

If participants are withdrawn from the study for any of the reasons noted above during Baseline I or II (prior to randomization), they will be replaced until a total of 200 participants have been randomized to the study. Subjects who voluntarily withdraw from the study will be asked to complete a questionnaire regarding the reasons for dropping out and what they didn't/did like about the study.

### 4.0 Recruitment Methods

#### 4.1 Identification of subjects

All recruitment for this study will be routed through IRB Protocol #2213 which will also serve as the initial recruitment point of contact.

#### 4.2 Recruitment process

Interested volunteers calling the study center number will first complete the eligibility script and questions for IRB Protocol #2213. If a participant's responses match our study's specified inclusion criteria they will be forwarded to our study's coordinator for further screening.

#### 4.3 Recruitment materials

All study materials for this project will be approved through IRB Protocol #2213. To summarize, materials in that project include letters, newsletters, flyers and radio/print text. Participants will be recruited from the Harrisburg/Hershey/Lancaster community by using the following resources:

a) Media advertisements (newspaper, radio, internet)
b) Study posters/flyers placed on community message boards and in local businesses, medical clinics
c) Community newsletters including the Hershey Medical Center employee newsletter
d) Newsletters/letters sent to past study participants who consented to being contacted about future research
e) Internet locations including but not limited to:
    i. HMC Facebook page
    ii. Penn State TCORS website
    iii. Craigslist

.

#### 4.4 Eligibility/screening of subjects

1. **Screening 1 (phone):** We will consider the screening process and eligibility questions in IRB protocol #2213 as Screening 1. This process includes a brief phone screening to determine basic eligibility for any of our study center protocols. Then, participants will complete the screening for this study in two additional steps.

2. **Screening 2 (phone):** A full script and screening questions for this study are in the "Consent Forms and Recruitment Materials" section of the IRB application.

3. **Screening 3 (In person, Visit 1):** After a participant has met basic eligibility criteria over the phone, they will be scheduled to come into the study center where they will be consented to the study and further screened for eligibility. See section 7.2 for further details.

### 5.0 Consent Process and Documentation

#### 5.1 Consent Process

##### 5.1.1 Obtaining Informed Consent

###### 5.1.1.1 Timing and Location of Consent

When participants attend their first in person assessment visit, they will have the study explained to them in detail, have the opportunity to ask questions and then will be asked to sign the consent form. Participants will be given a signed copy of the form. This will take place in a private clinic room at the Penn State CTSI.

###### 5.1.1.2 Coercion or Undue Influence during Consent

Once potential study volunteers are identified they will be given information about the study and offered the opportunity to participate. The researchers obtaining consent will be instructed to clearly indicate that the participant's enrolling in the trial is purely voluntary and the researchers will not offer comments about whether they believe the participant should enroll in the study or not. Given the number of contacts and visits involved in the study protocol, the compensation provided to the participant is modest.

###### 5.1.2 Waiver or alteration of the informed consent requirement

N/A

##### 5.2 Consent Documentation

###### 5.2.1 Written Documentation of Consent

An IRB approved consent form will be used to document consent. Both the researcher and the participant will retain a copy of the consent. The participant's signed consent form will be periodically uploaded into the participant's REDCap record at the Hershey site. Harvard/Mass General will store their consents in a locked filing cabinet in a locked research office.

###### 5.2.2 Waiver of Documentation of Consent

Participants who are interested in the study will be asked to consent to allow the researcher to pre-screen them for the study over the phone by asking all screening questions of all participants (eligible or ineligible). Participants will be asked if this information can be retained so that the study team will know reasons that participants are not eligible for the study.

In addition, participants who are not eligible for the study, or those who begin the phone screener but are not interested in completing it after learning more about the study, will be asked if they would be interested in being contacted for future studies being conducted by our research team. They will be informed that by providing their name and phone number, they will be consenting to allow the study team to contact them in the future.

##### 5.3 Consent – Other Considerations

###### 5.3.1 Non-English Speaking Subjects

Non-English speaking subjects will not be eligible for this study because all study visits and materials need to be conducted/provided in English.

###### 5.3.2 Cognitively Impaired Adults

Cognitively impaired adults will not be eligible for this study.

###### 5.3.2.1 Capability of Providing Consent

N/A

###### 5.3.2.2 Adults Unable To Consent

N/A

###### 5.3.2.3 Assent

N/A

###### 5.3.3 Subjects who are not yet adults (infants, children, teenagers)

###### 5.3.3.1 Parental Permission

N/A

###### 5.3.3.2 Assent

N/A

#### 6.0 HIPAA Research Authorization and/or Waiver or Alteration of Authorization

##### 6.1 Authorization and/or Waiver or Alteration of Authorization for the Uses and Disclosures of PHI

**Check all that apply:**

X Authorization will be obtained and documented as part of the consent process.

☐ Partial waiver is requested for recruitment purposes only *(Check this box if patients' medical records will be accessed to determine eligibility before consent/authorization has been obtained)*

☐ Full waiver is requested for entire research study *(e.g., medical record review studies)*

X Alteration is requested to waive requirement for written documentation of authorization

##### 6.2 Waiver or Alteration of Authorization for the Uses and Disclosures of PHI

###### 6.2.1 Access, use or disclosure of PHI representing no more than a minimal risk to the privacy of the individual

**6.2.1.1 Plan to protect PHI from improper use or disclosure**
All data collected during the screening process will be directly entered into REDCap.

**6.2.1.2 Plan to destroy identifiers or a justification for retaining identifiers**
All study data will be retained indefinitely.

###### 6.2.2 Explanation for why the research could not be practicably be conducted without access to and use of PHI

N/A

###### 6.2.3 Explanation for why the research could not practicably be conducted without the waiver or alteration of authorization

In order to screen the participants prior to inviting them into the study center, the investigators are conducting a phone screen to determine if the participants are likely to be eligible for the study.

##### 6.3 Waiver or alteration of authorization statements of agreement

Protected health information obtained as part of this research will not be reused or disclosed to any other person or entity, except as required by law, for authorized oversight of the research study, or for other permitted uses and disclosures according to federal regulations.

The research team will collect only information essential to the study and in accord with the 'Minimum Necessary' standard (information reasonably necessary to accomplish the objectives of the research) per federal regulations.

Access to the information will be limited, to the greatest extent possible, within the research team.

All disclosures or releases of identifiable information granted under this waiver will be accounted for and documented.

#### 7.0 Study Design and Procedures

##### 7.1 Study Design

This is a two-arm, randomized, double-blind parallel group trial that will proceed in three phases over 33weeks. Participants and study staff will be blind to the experimental cigarette allocation until the end of the last visit. Visits will occur at consistent times during the day, when possible. For participants who do not show up for scheduled study visits up to 5 attempts will be made to contact them to reschedule their appointment. After the 5th attempt, a letter will be sent to the address on file informing them that the study team has been trying to reach them and asking them to contact the study center to reschedule. No further attempts to contact the participant will be made after the letter is sent (see study documents for letter).

1. **Baseline Phase:**
   **Baseline I:** participants will smoke their own usual brand of cigarettes for 1 week.
   **Baseline II**: all participants will be asked to smoke research cigarettes containing a normal amount of nicotine (11mg) and matching the flavor (regular or menthol) of their usual brand of cigarettes for 2 weeks.
2. **Randomized Double-Blind Nicotine Reduction Phase (18 weeks)**, participants who complete **Baseline II** and agree to continue will enter this phase. They will be randomized to either (1) continue to smoke the same research cigarettes they smoked in Baseline II for 18 weeks with their usual nicotine content (UNC) or (2) switch to progressively reduced nicotine content (RNC) cigarettes over 18 weeks.
3. **Treatment Choice Phase (12 weeks)**
   Prior to making their "Treatment Choice", all participants will receive a copy of the Surgeon General Report, "How Tobacco Causes Disease" and resources they can contact for help. Regardless of the participant's choice, all participants will attend two additional follow up visits 4 weeks (visit 11) and 12 weeks (visit 12) after the end of the randomized phase.

   **Choices that will be provided to the participant include:**
   1. Return to their usual brand
   2. Continue on the research cigarettes for a further 12 weeks (provided at no cost)
   3. Receive assistance to quit smoking (with counseling and short acting nicotine replacement therapy medication provided at no cost) for 11 weeks.

**7.2 Study Procedures**

**Screening [Day 0]**

After a participant has met basic eligibility criteria over the phone, they will be scheduled to come into the study center where they will be consented to the study and further screened for eligibility.

Informed consent will be obtained by research staff. During this process, the usual discussion of procedure, risks, side effects, confidentiality, voluntary participation, and right to refuse participation without prejudice will be explained to the participant. Participants must be capable of understanding the nature of this study, its potential risks, discomforts and benefits before signing consent.

**Measures and Procedures:**
The following information will be collected to determine eligibility:

1. NIDA Drug Screener (not eligible if any drugs are used weekly or more)
2. Medical history
3. Concomitant medication history to ensure that the participant is not taking any medications that may make them ineligible for the study
4. Measure vital signs. (inclusion systolic $\leq$160)
5. Pregnancy determination: Obtain Urine and process pregnancy test for women of child bearing age (have had a period in the past 12 months) who have not had a hysterectomy
6. Complete the MINI (inclusion=positive current or lifetime diagnosis for mood or anxiety disorder), MINI Suicide score is < 9

**Procedure for suicide risk at screening:** Scores of $\geq$ 9 on the MINI Suicide module will require an assessment with a licensed clinician to document a clinical plan. Participant with a score >8 will not continue in the study unless judged to be stable and not at high risk for suicide.

**Baseline I (1 week on own brand of cigarettes)**

After a participant is determined to be eligible for the study, they will complete the

| Time and Events Table | Baseline | | | | Randomization | | | | | | Treatment Choice | |
|---|---|---|---|---|---|---|---|---|---|---|---|---|
| | Baseline I | | Baseline II | | Step 1 | Step 2 | Step 3 | Step 4 | Step 5 | | | |
| Study Week Number | 1 | 2 | 3 | 4 | **7** | **10** | **13** | **16** | **19** | **22** | **26** | **34** |
| Study Day | 1 | 8 | 15 | 22 | **43** | **64** | **85** | **106** | **127** | **148** | **176** | **232** |
| Study Visit Number | 1 | 2 | 3 | 4 | **5** | **6** | **7** | **8** | **9** | **10** | **11** | **12** |
| **Measures/Questionnaires** | | | | | | | | | | | | |
| Tobacco and marijuana use and daily cigarette log* | | X | X | X | X | X | X | X | X | X | X | X |
| Concomitant medications* | | X | X | X | X | X | X | X | X | X | X | X |
| Adverse events* | | X | X | X | X | X | X | X | X | X | X | X |
| Demographics | X | | | | | | | | | | | |
| Tobacco use history, cigarette details* | X | | | | | | | | | | | |
| NIDA drug use (past 3 mo) | x | | | | | | X | | | X | | |
| Environmental smoke questionnaire | | X | | X | X | X | X | X | X | X | X | X |
| Perceived health risk | | X | | X | | | X | | | X | X | X |
| Cigarette liking scale | X | X | X | X | X | X | X | X | X | X | | |
| Nicotine dependence (HONC, FTND, PSU) | X | X | X | X | X | X | X | X | X | X | X | X |
| Minnesota withdrawal scale | X | X | X | X | X | X | X | X | X | X | X | X |
| Questionnaire smoking urges | X | X | | X | X | X | X | X | X | X | X | X |
| Audit-C | | X | | X | | X | | X | | X | | |

questionnaires, biomeasures and procedures outlined in the time and events table.

The following procedures will be conducted at all Baseline visits:

- Participants will be instructed on standard study procedures, online surveys, phone calls and on how to use the study cigarette log, will be given a supply of logs and small pencils, and will be asked to log the total number of cigarettes they smoke each day. The participants will be given the opportunity to ask any questions they may have regarding keeping their log.

- The participant and a member of the study staff will review a preliminary schedule of study visits, dates and times. Any scheduling conflicts will be resolved and the participant provided with a new schedule of visits. The importance of keeping on schedule with appointments will be discussed.

- Participant will strongly advised not to use other forms of tobacco or marijuana. Participants will also be asked to honestly report any other tobacco or marijuana use at subsequent visits.

- Participants will be asked to rate their confidence in complying with the study protocol on a scale from 1-10, with 1 being no confidence and 10 being complete confidence. Any scores < 6 will prompt the researcher to troubleshoot any challenges identified by the participant.

**Procedure for suicide risk at all visits where QIDS questionnaire is administered:** If participants indicate an answer of > 0 on item 12 of the QIDS questionnaire, the MINI Suicide module will be administered. Scores of $\geq$ 9 on the MINI Suicide module will require an assessment with a licensed clinician to document a clinical plan and a determination of retention or termination from the study.

| | | | | | | | | | | | | |
|---|---|---|---|---|---|---|---|---|---|---|---|---|
| Anxiety symptoms questionnaire (ASQ) | X | | | | | | X | | | X | X | X |
| CES-D | X | | | | | | X | | | X | X | X |
| Kessler K6 | X | X | | X | X | X | X | X | X | X | X | X |
| OASIS (anxiety) | X | X | | X | X | X | X | X | X | X | X | X |
| QIDS (depression) | X | X | | X | X | X | X | X | X | X | X | X |
| Perceived stress scale | X | X | | X | X | X | X | X | X | X | X | X |
| Clinical COPD questionnaire | | X | | X | X | X | X | X | X | X | X | X |
| Pittsburgh sleep symptom and Dreams questionnaires | X | | | | | | | | | | | |
| Interheart and WI Prepare | X | | | | | | | | | | | |
| Smoking Cessation (if chosen by participant) | | | | | | | | | | X | X | X |
| NRT disbursement (if chosen by participant) | | | | | | | | | | X | X | |
| Biomeasures/Procedures¥ | | | | | | | | | | | | |
| Weight | X | X | X | X | X | X | X | X | X | X | X | X |
| Height | X | | | | | | | | | | | |
| Waist hip ratio | X | | | | | | | | | X | | X |
| Exhaled CO | X | X | X | x | X | X | X | X | X | X | X | X |
| Blood pressure/pulse | X | X | X | X | X | X | X | X | X | X | X | X |
| Pulmonary function | X | X | | | X | | | X | X | X | X | X |
| Urine ( NNAL, 1-HOP, F2-Isoprostanes, Creatinine, ) | | X | | X | X | X | X | X | X | X | X | |
| Pregnancy test | X | | | X | | X | | X | | X | X | X |
| Blood (cotinine, GSH, GSSP, GSSP, Hemoglobin, DNA) | | X | | X | X | X | X | X | X | X | X | |
| Payment | $40 | $80 | $40 | $80 | $80 | $80 | $80 | $80 | $80 | $80 | $80 | $40+ compliance |
| * To be completed by the researcher. All others will be completed by the participant. ¥All biomeasures/procedures to be completed by researcher. Some biomarkers will be analyzed in selected subgroups of collected samples. | | | | | | | | | | | | |

**Visit 1 [Day 1]**
**Cigarettes:** Participants will be instructed to smoke their usual brand of cigarettes for the next week.

**Visit 2 [Day 8, ± 3 days]**
- Review of participants continued compliance and eligibility.
- The study cigarette log will be reviewed and the total number of cigarettes smoked each day will be recorded.
- Participants will be given a 1 week supply of research cigarettes (150% of baseline cigarettes per day) that has a normal nicotine content (approx. 11 mg). They will be instructed to smoke as they usually do.
- Compliance with the study protocol including not using other forms of tobacco or marijuana will be stressed.
- Participants will be asked to honestly report any use of non-research cigarettes, other tobacco or marijuana at subsequent visits.
- Participants will be instructed to bring all cigarette packs (unopened, opened or empty) back to the study center at each study visit. Study staff will record the number of cigarette smoked and returned. Participants will be reminded that they will receive a final payment based on the accuracy of the packs they return.

**Baseline II (2 weeks on usual nicotine content research (UNC) cigarettes)**
The following procedures will be completed at each visit during Baseline II:
- Participants will be evaluated for continued compliance and eligibility for the study.

- Compliance with the study protocol including not using non-research cigarettes, other forms of tobacco or marijuana will be stressed.
- Participants will be asked to rate their confidence in complying with the study protocol on a scale from 1-10, with 1 being no confidence and 10 being complete confidence. Any scores < 6 will prompt the researcher to troubleshoot any challenges identified by the participant.
- Participants will be asked to honestly report any use of non-research cigarettes, other tobacco or marijuana.
- Participants will be instructed to bring all cigarette packs (unopened, opened or empty) back to the study center at each study visit.

**Visit 3 [day 15]**

Participants will be given a 1 week supply of research cigarettes (150% of baseline cigarettes per day) that approximately match the nicotine content of their usual brand of cigarettes.

**Visit 4 [day 22]**
Participants who are eligible to continue to randomization will be given a choice to complete one of their weekly contacts via REDCap survey at home if they have a personal email address and reliable internet access.

Participants will be given a 3 week supply of either UNC or RNC research cigarettes (150% of baseline cigarettes per day) corresponding to the participant's treatment group allocation (see table in section 7.4.2).

**Randomized Double Blind phase (18 weeks on either UNC or gradually reduced nicotine content (RNC) cigarettes)**
The following procedures will be conducted at each phone call or online survey during this phase:

**Phone call or online survey [Day 29, 50, 71, 92, 113, 134]**
Based on the participant's choice at Visit 4, they will either receive a phone call or complete an online REDCap survey via an email link. During this contact, they will complete the daily cigarette log and other tobacco use questionnaire, the Minnesota Withdrawal Scale and the Cigarette Liking Scale. No personal information will be collected. If the participant does not complete this contact, they will still be eligible to proceed in the study. However, every effort will be made to collect data retrospectively for any missed contacts at the next in person participant contact.

**Payment:** $10

**Phone call [Day 36, 57, 78, 99, 120, 141]**
The main purpose of this contact is to remind the participant of their scheduled visit in 1 week and to collect daily cigarette log information. If the participant does not complete this contact, they will still be eligible to proceed in the study however every effort should be made to collect data retrospectively for any missed contacts at the next in person participant contact.

**Payment:** None

**The following procedures will be followed for all in person visits during this phase:**

- Participants will complete the questionnaires, biomeasures and procedures outlined in the "Randomized Double Blind Phase: Measures, questionnaires and procedures" Refer to the time and events table.
- The study cigarette log will be reviewed and the total number of cigarettes smoked in the previous 6 days will be recorded (mean cigarettes per day will be imputed during statistical analysis for any missing days).
- A schedule of all study contact dates and times will be reviewed with the participant (online surveys, phone calls and in person visits). Any scheduling conflicts will be resolved and the participant provided with a new schedule of visits.
- Participants will be asked to rate their confidence in complying with the study protocol on a scale from 1-10, with 1 being no confidence and 10 being complete confidence. Any scores < 6 will prompt the researcher to troubleshoot any challenges identified by the participant.
- Participants will be asked to honestly report any use of non-research cigarettes, other tobacco or marijuana at subsequent visits. Compliance with the study protocol will be stressed.
- All cigarette packs from the previous weeks (unopened, opened or empty) will be collected by the researcher.
- Participants will be reminded to bring all cigarette packs (unopened, opened or empty) back to the study center at each study visit
- Participants will be given a 3 week supply of either UNC or RNC research cigarettes (150% of baseline cigarettes per day) corresponding to the participant's treatment group allocation (see table in section 6.4.2).

**Study window for all randomization phase visits:** ± 7 days (no study window for phone calls or surveys) when possible

**Payment:** Participants will receive $80/visit ($20 for transportation, $20 for study visit completion and $40 for blood and urine) for every successfully completed study visit during the randomized phase

**Visit 10 [Day 148]**
**Treatment Choice:**
All participants will be encouraged to quit smoking and will be given information and resources to help smokers to quit (including the Surgeon General's Report on "How Tobacco Causes Disease"). Participants will then be given the choice to either

1. Continue on the research cigarettes they have been using, for the next 12 weeks,
2. Return to their usual brand of cigarettes, or
3. Quit smoking with brief counseling from the study team and the option to use oral nicotine replacement (11 weeks of gum or lozenge). Participants who choose to quit smoking will be offered a flexible treatment plan that includes optional extra contacts (2 in person and 4 possible phone calls). There will be no payments for the additional visits completed due to a participant choosing this option.

**Cigarettes:**

- If participants choose to continue on research cigarettes, they will be given a 4 week supply of either UNC or RNC research cigarettes (150% of baseline cigarettes per day) corresponding to the cigarettes they were given in the last phase of the participant's treatment group allocation (see table in section 7.4). Participants will be instructed to bring all cigarette packs (unopened, opened or empty) back to the study center at their next visit.
- If participants choose to return to their usual brand of cigarettes, they will not be given any further research cigarettes.
- If participants choose to quit smoking they will be given up to a 6 day's supply of research cigarettes corresponding to the cigarettes they were given in the last phase of the

participant's treatment group allocation (see table in section 7.4). These cigarettes are intended to last until their target quit date

**Participants Who Choose to Make a Quit Attempt:** These participants will be offered a flexible smoking cessation treatment in which they will have the option to receive up to 11 weeks of short acting nicotine replacement therapy (gum or lozenges) at no cost and cognitive behavioral based smoking cessation counseling provided by study staff either in person or over the phone. Participants interested in quitting smoking must be willing to set a quit date one day before the week 23 visit. These participants will be provided with up to a 6 day's supply of research cigarettes to use until their quit date.

By this visit, some participants will have transitioned to smoking very low nicotine content research cigarettes. In order to select an appropriate nicotine replacement (NRT) dose, participants will be asked to quit smoking the day before their next visit (week 23). Participants will be encouraged to call the state telephone quit line and/or use the Freedom from Smoking (FFS) quit website (http://www.ffsonline.org/) to supplement the brief, in-person smoking cessation counseling sessions provided by study staff.

The following contact schedule will be provided to the participant:

| **Schedule of contacts for participants who choose to quit smoking during the treatment choice phase** | | | | | | | | | | | | | | | |
|---|---|---|---|---|---|---|---|---|---|---|---|---|---|---|---|
| | Wk22<br><br>End randomized phase | -1d | Wk 23<br><br>Day 155 | +1d | Wk 24<br><br>Day 162 | Wk 25<br><br>Day 169 | Wk 26<br><br>Day 176 | Wk 27<br><br>Day 183 | Wk 28<br><br>Day 190 | Wk 29<br><br>Day 197 | Wk 30<br><br>Day 204 | Wk 31<br><br>Day 211 | Wk 32<br><br>Day 218 | Wk 33<br><br>Day 225 | Wk 34<br><br>Day 232 |
| In Person counseling session | X | | X | | | | X | | | | X | | | | |
| Telephone session | | | | X | X | | | | X | | | | X | | |
| Self guided session | | | | | | X | | X | | X | | X | | X | |
| Quit day | | X | | | | | | | | | | | | | |
| Possible NRT disbursement | | | X | | | | X | | | | X | | | | |
| End of NRT use | | | | | | | | | | | | | | | X |

**Treatment Choice Phase (12 weeks of participant choice to return to usual brand, continue on research cigarettes, or quit smoking with help)**

**All Participants During Treatment Choice Phase:**
The following procedures will be completed at study visit 11 and 12:

- Participants will complete the questionnaires, biomeasures and procedures outlined in the "Treatment Choice Phase: Measures, questionnaires and procedures" table below.
- The study cigarette log will be reviewed and the total number of cigarettes smoked will be recorded. The participants will be given the opportunity to ask any questions they may have regarding keeping their log. If needed, participants will be re-instructed on how to complete the log and will be given a supply of logs and small pencils.
- A schedule of the study contact dates and times will be reviewed with the participant (online surveys, phone calls and in person visits). Any scheduling conflicts will be resolved and the participant provided with a new schedule of visits.

**Cigarettes:**

- Participants will be asked to honestly report any use of non-research cigarettes (for those who continued on research cigarettes), other tobacco or marijuana.
- If participants chose to receive research cigarettes, all cigarette packs from the previous weeks (unopened, opened or empty) will be collected by the researcher.
- Participants will be instructed to bring all cigarette packs (unopened, opened or empty) back to the study center at their next visit.
- If participants choose to return to their usual brand of cigarettes, they will not be given any further research cigarettes.
- If participants choose to quit smoking they will not receive any research cigarettes even if they were not successful in quitting.

**Procedure for suicide risk at all visits where QIDS questionnaire is administered:** If participants indicate an answer of > 0 on item 12 of the QIDS questionnaire, the MINI Suicide module will be administered. Scores of ≥ 9 on the MINI Suicide module will require an assessment with a licensed clinician to document a clinical plan and a determination of retention or termination from the study.

**Payment:** Participants will receive $80 ($20 for transportation, $20 for study visit completion and $40 for blood and urine).

**Study window for all Treatment Choice Phase visits (not including additional visits for those who are making a quit attempt):** ± 7 days when possible

**Participants Who Choose to Make a Quit Attempt:**
**Week 23 [Day 155]**

At this visit participants will be evaluated by the researcher for nicotine withdrawal symptoms. If the participant chooses to use NRT they will receive 3 boxes of either nicotine gum (110 pieces/box) or lozenges (81 pieces/box) (according to participant's preference). At Hershey, NRT will be dispensed by the Penn State Investigational Drug Service (IDS). Adverse Events and concomitant medications will be assessed and a questionnaire (Smoking Cessation Quit Day) that includes the Minnesota Withdrawal Scale (MNWS) will be completed by the participant. Decisions about dosing will be determined based on participant reported withdrawal symptoms. Participants who will have tapered to very low nicotine cigarettes should have minimal withdrawal symptoms and will require very low NRT dose. A general guideline for NRT Dosing will be used to make recommendations to participants as follows:

- Group 1:
  - Not able to remain abstinent or
  - Reports a score of 2 or more on the MNWS items #1 and #4 (irritable/ angry and/or craving to smoke)
- Group 2:
  - Able to remain abstinent and
  - Reported a score of 0 or 1 on MNWS items #1 and #4 or
  - had few slips

| **2mg Oral NRT (gum or lozenge) dosing guidelines for participants based on withdrawal symptom group allocation** | | | |
|---|---|---|---|
| | Group | Max. number of pieces/day | Duration |
| Wk 23 | 1 | 1 piece as needed, not more than 1 every 1 hr or 16 in 24 hours | 3 weeks |
| | 2 | 1 piece as needed, not more than 1 every 4 hrs or 6 in 24 hours | 3 weeks |
| Wk 26 | 1 | 1 piece as needed, not more than 1 every 2 hrs or 12 in 24 hours | 4 weeks |

| | | | |
|---|---|---|---|
| | 2 | 1 piece as needed, not more than 1 every 6 hrs or 4 in 24 hours | 4 weeks |
| Wk 30 | 1 | 1 piece as needed, not more than 1 every 4 hrs or 6 in 24 hours | 4 weeks |
| | 2 | 1 piece as needed, not more than 1 every 8 hrs or 3 in 24 hours | 4 weeks |

The day after this visit, the researcher will call the participant to assess whether the participant is experiencing any side effects consistent with high nicotine levels and assess reducing the dose of NRT. As needed, additional courses of NRT will be given to the participant either at study visits or if the participant calls in to the study center to request additional NRT. At week 26 and 30 the dosing schedule will be discussed with the participant (see "Schedule of Contacts for participants who choose to quit" table in section 7.2).

**General visit, phone, and self-guided sessions:**
In addition to regular study visits (weeks 26 and 34), there will be two additional in person sessions (weeks 23 and 30), four phone sessions (weeks 23, 24, 28, and 32) and five self-guided sessions. In order to avoid missed phone visits, researcher and participant will attempt to agree on a standing appointment, day and time. At each contact expired CO will be measured, adverse events and concomitant medications will be assessed and participants will complete questionnaires. Participants will receive 20 minutes (or less) of standard individual cognitive behavioral therapy (CBT) based on the *Freedom from Smoking* (FFS) curriculum (http://www.ffsonline.org/), from the American Lung Association. They will be given strategies to cope with triggers/urges to quit and when appropriate, the researcher will help the participant make a test call to the quitline and/or use the FFS website.

**Week 26 [Day 176]**
If participants chose to receive research cigarettes, they will be given an additional 8 week supply of either UNC or RNC research cigarettes (150% of baseline cigarettes per day) corresponding to the cigarettes there were given in the last phase of the participant's treatment group allocation (see table in section 7.4).

Participants who chose to use NRT to quit smoking, could be given a refill of oral NRT (3 boxes of gum or lozenge) as deemed appropriate by the researcher according to the participant's NRT usage and withdrawal symptoms.

**Week 34 [Day 232]**
**Cigarettes and cessation counseling:** No further counseling, NRT or study cigarettes will be given to the participants.

**Payment:** In addition, those participants who are eligible for the compliance incentive payments for good study compliance will be informed. There are two possible compliance incentive payments:

- **Study visit attendance ($50):** Participants will be eligible for this compliance incentive if they attended all study visits and provide data.
- **Cigarette pack return accuracy ($50):** Participants will be eligible for this study compliance incentive if they returned all research packs (opened and unopened) within a margin of 4 packs for 6/8 study visits where research cigarette return was required (visit 3. 4, 5, 6, 7, 8, 9, and 10).

##### 7.3 Duration of Participation
Participants who complete all phases of the study will participate in the study for 33 weeks.

#### 7.4 Test Article(s) (Study Drug(s) and/or Study Device(s))

##### Description

Participants will be provided with research cigarettes that have a similar nicotine content to current commercially available cigarettes (11mg). Menthol smokers will be provided with menthol research cigarettes if that is their preference. The following table describes the dosing and schedule for all study participants including the reductions for those randomized to the intervention during the Double-Blind Phase.

At visit 10 participants enter the Treatment Choice Phase of the study. At this timepoint, participants are given a choice to go back to their own brand of cigarettes, to quit, or to continue receiving research cigarettes for an additional 3 months. If a participant is in the RNC arm and they choose to continue with the research cigarettes for 3 more months, these participants will receive the same dose of cigarettes they received during the previous 3 weeks.

For those who are in the UNC group and choose to continue on the research cigarettes, they will receive either the same dose they received during the previous 3 weeks OR they will receive the Step 1 RNC cigarettes (see table below) depending on supply availability.

| **Phase:** | **Baseline I** | **Baseline II** | **Randomized Double-Blind Nicotine Reduction Phase** | | | | | **Treatment Choice Phase** |
|---|---|---|---|---|---|---|---|---|
| **Wks/Phase:** | **2** | **2** | **3** | **3** | **3** | **3** | **3** | **12** |
| **Cigarette type:** | Own Brand | Usual Nicotine Research Cigarettes | Reduced Nicotine Step 1 | Reduced Nicotine Step 2 | Reduced Nicotine Step 3 | Reduced Nicotine Step 4 | Reduced Nicotine Step 5 | Variable |
| | *Nicotine content (mg/cigarette)* | | | | | | | |
| **RNC** *regular:* | Around 11mg[3] | 11 | 7.8 | 3.2 | 1.3 | 0.7 | 0.2 | Variable |
| **RNC** *menthol:* | | 12.1 | 7.0 | 3.4 | 1.4 | 0.7 | 0.2 | |
| **UNC** *regular:* | Around 11mg | 11 | 11 | 11 | 11 | 11 | 11 | Variable |
| **UNC** *menthol:* | | 12.1 | 12.1 | 12.1 | 12.1 | 12.1 | 12.1 | |

##### 7.4.3 Method for Assigning Subject to Treatment Groups

Participants will be randomized to one of two experimental conditions based on a pre-determined random number sequence generated by Dr. Liao (head of PS TCORS Data Management Core). A Cigarette Management System (CMS) has been created by the Public Health Sciences Data Management team to manage assigning randomized, blinded cigarettes to participants and to track cigarette inventory. This system will store the randomization code. Randomization will be stratified by site (Penn State and Mass. General/Harvard) and by menthol flavor (regular/menthol). This ensures a similar distribution of treatment assignment (1:1 for gradual reduction: control) across study sites to avoid potential confounding.

##### 7.4.4 Subject Compliance Monitoring

Participants will be given a daily log to complete on which they will tally the number of cigarettes they smoke each day. Participants will be given detailed instructions on how to complete the log and they will be reminded at each study contact to only use the research cigarettes they have been given. Questions will be asked at each contact to review their log and verify how many cigarettes they are smoking each day. Participants will also be asked to self-report their other tobacco use including products such as cigars, pipes, chew, snus, dip, hookah, electronic cigarettes and dissolvable tobacco. Exhaled carbon monoxide measurements and blood will be taken throughout the trial to verify smoking intensity, nicotine and cotinine levels.

##### 7.4.5 Blinding of the Test Article

Cigarette cartons will arrive in the IDS with a packaging slip that provides information about the cartons in the shipment which includes the nicotine content, carton bar code number and a batch/lot number. This information will be carefully recorded by the Cigarette Manager in our CMS (Cigarette Management System), removed from the carton and replaced with a blind code number. Individual packs do not contain any identifiable information. Each individual pack will be labeled with the carton blind code.

##### 7.4.6 Receiving, Storage, Dispensing and Return

###### 7.4.6.1 Receipt of Test Article

Experimental cigarettes will be provided free of charge to the investigators via the NIDA Drug Supply Program (see Notice: NOT-DA-13-002). Cartons (10 packs/carton) with packs of 20 cigarettes/pack will be received from the Research Triangle Institute in North Carolina.

###### 7.4.6.2 Storage

At Penn State Hershey, cigarettes will be stored in locked, -20 freezers that are maintained by the IDS. Standard pharmacy protocols for freezer temperature control monitoring and recording will be followed. Prior to cigarette assignment, the cigarettes may be stored unfrozen for a period of time up to 3 weeks. At Harvard/Mass General, cigarettes will be stored at room temperature in locked cabinets in a locked office space. At both sites, menthol cigarettes will be stored separately from regular cigarettes.

###### 7.4.6.3 Preparation and Dispensing

**Assignment:** When a new participant is entered into the CMS and is at the study visit where they receive cigarettes, the Cigarette Manager will enter the Participant ID, baseline cigarettes per day, and flavor preference into the CMS. The CMS will provide the Cigarette Manager with the Blind Code numbers for the appropriate cartons of cigarettes (e.g., the correct cigarettes/nicotine level for the corresponding stage for where the participant is in the study).

**Preparation:** Once the cigarettes are received, they will be blinded by the Cigarette Manager under the supervision of the IDS. At the time of dispensation, the CMS will instruct the Cigarette Manager which blind codes should be dispensed to the participant and the Cigarette manager will pull the correct cartons from the cigarette inventory. The CMS will also tell the Cigarette Manager how many packs of cigarettes to dispense to the participant. The cigarette manager will prepare a bag of cigarette packs that will be given to the study coordinator for the participant.

**Where:** All receiving, sorting, blinding of the initial study product will be done in IDS pharmacy space at Penn State Hershey. Dispensing of product to study participants will also be done in the IDS pharmacy space at Penn State Hershey. At Harvard/Mass General, cigarettes will be stored in locked cabinets in a locked research space and blinded cartons will prepared for dispensing in that space.

**Dosing:** Dosing of cigarettes will follow the schedule outlined in section 7.4.

**Dispensing:** Cigarette packs containing 20 cigarettes will be dispensed to the study coordinator during or prior to a participant's in person visit. All packs will be given to participants in the original packaging. Each individual pack of cigarettes will have a label with the participant ID number, the blind code number, the flavor (regular or menthol) and a list of possible nicotine doses the cigarettes may contain. Packs will also be labeled with a color coded sticker that indicates the visit number at which they were dispensed. The coordinator will give the cigarettes to the participant at their visit. The number of packs dispensed will be determined by the self-reported number of cigarettes per day they smoke when they enroll in the trial. Participants will be provided with 150% of the cigarettes necessary for each week of usual smoking. Participants will be able to smoke as many or as few cigarettes as they choose and will be required to return all packs at the next visit.

###### 7.4.6.4 Return or Destruction of the Test Article

The CMS will track all receipt and dispensing of cigarettes for the duration of the project. Reconciliation reports will be produced on an ongoing basis.

Participants will be instructed to return all research cigarette packs at the next visit, regardless of whether they are empty, unopened or partially used. Any unused experimental cigarettes that are returned from the participant will be destroyed by the IDS. Harvard/Mass General will follow their site's drug destruction policy. We will not ship experimental cigarettes back to the NIDA Drug Supply Program or recycle opened cartons.

###### 7.4.6.5 Prior and Concomitant Therapy

Concomitant medication use will be collected regularly throughout the trial to monitor participant health conditions.

Participants taking varenicline, bupropion or nortriptyline as a smoking cessation medication in the prior month will be excluded from the study. Participants taking bupropion or nortriptyline for depression management and who expect to continue use of the medication throughout the trial will be eligible to participate. Additionally, participants who are prescribed bupropion or nortriptyline for depression management at any point during the study will be eligible to continue with the study. Medications related to certain medical conditions that are exclusions to the study, such as COPD and current heart conditions, will serve to alert the study staff of the presence of these conditions during screening. Once the participant is entered into the randomized double blind phase of the study, there are no medications that will interfere with the participant's ability to participate.

### 8.0 Data and Specimen Banking

Blood and urine will be banked for future undetermined research. Participants will be given an option on the consent form that will provide permission for the researchers to bank their specimens for future projects. Only participants who consent to this option will have their specimens banked.

##### 8.1 Data and/or specimens being stored

Specimens will be stored with an ID code attached. The location of the specimen in the study freezer will be managed using Freezerworks database which will be housed on the PHS password secured network. The code number, visit number, date/time of collection, processing and storage, and consent options for future use of samples will be stored in Freezerworks in addition to REDCap. All other data associated with the ID code will be retained in REDCap.

##### 8.2 Location of storage

Specimens will be stored a locked freezer room in the research laboratory of Drs. Muscat & Foulds on the 3rd floor of the Cancer Institute.

##### 8.3 Duration of storage

Specimens will be stored indefinitely with code number attached. Data will be stored indefinitely with identifiers attached in REDCap and no identifiers in Freezerworks.

##### 8.4 Access to data and/or specimens

The lab managers, technicians, study coordinators and PI will have access to the freezer rooms where the specimens will be stored. The researchers will have access to the stored data in REDCap although role specific rights will be granted to forms (i.e., researchers who see participants will not have access to data that may allow them to become unblinded to a participant's treatment allocation, Harvard/Mass General coordinators will not have access to Penn State Hershey participant data, etc.).

##### 8.5 Procedures to release data or specimens

Investigators who are interested in obtaining samples from this project for ancillary studies will first be required to submit a detailed written proposal to Dr. Foulds. Dr. Foulds will then take the proposal to the overall Penn State Tobacco Center of Regulatory Science (TCORS) steering committee for review and approval. If the proposal is approved, the investigator will then need to obtain all other regulatory approvals (IRB, departmental scientific committees, etc.) prior to samples being released. Only de-identified data, as approved in the investigator's IRB application, will be released to the investigator. Blood and urine samples will only be released if the participant provided written consent to allow their samples to be used by other investigators (this option is included in the original consent form).

##### 8.6 Process for returning results

Investigators will be required to provide a written report on their study results to the TCORS steering committee. Individual participants will not be provided with the results of the analyses of their samples.

#### 9.0 Statistical Plan

##### 9.1 Sample size determination

The primary outcome variable is plasma cotinine concentration (measured as ng/ml). The mean value of this continuous measurement will be compared between two groups: the experimental group (RNC cigarettes) and the control group (UNC cigarettes). For plausible effect size and variation, we use results of Benowitz et al (2012)(10), where the mean and standard deviation of plasma cotinine concentration are 240 and 120 for the control group and 113 and 116 for the experimental group at the 22nd week follow-up visit (end of randomization phase for trial). It is expected that the cotinine level in the experimental group will reduce gradually after start of the trial and the mean difference of the cotinine level between these two groups for our study will be smaller than the difference of 127 ng/ml in Benowitz et al (2012). With a sample size of 70 participants per group, we are able to detect the difference in plasma cotinine concentration level between the two groups as small as 58 ng/ml (which could occur on the 8.6mg nicotine content cigarettes) with at least 80% power (and 68 ng/ml with at least 90% power). Allowing for a 30% participant -withdraw rate, we plan to recruit 100 participants for each group (a total of 200). The alpha level used for the power analysis is 0.05.

Additional statistical power analyses were performed based on secondary hypotheses (e.g. hypothesis 1.b): to examine the difference in biomarkers of tobacco smoke exposure, nicotine withdrawal, and psychiatric symptomatology, with a sample size of 70 subject completers per group, we are powered to detect an effect size of 0.5 (difference is half of the standard deviation) between the two groups with at least 83% power. Also, with a completing sample size of 70 per group we are able to detect a 15 – 20% difference in the proportion of subjects willing to quit between these two groups (depending on the proportion in the control group) with about 80% power. If the proportion difference was as large as the value in the Benowitz et al (2012) study (51% of the RNC group versus 14% of the control group) then our statistical power will be as high as 99.8%. It was assumed that both groups have equal drop-out rate (30%) when the sample size was calculated above. However, it is possible that we could have differential drop-out rate in the two groups. Benowitz et al (2012) had 9% dropout in the control group versus 33% in the intervention group. We have 99% statistical power in detecting a difference as large as this.

##### 9.2 Statistical methods

***Preliminary Analyses:*** Basic baseline statistics including means (SDs), medians (IQR), and frequency distributions (percentages) will be reported for demographic characteristics; smoking history and other baseline measures. Ranges for all variables will be obtained to identify possible outliers and coding errors. Where necessary, some characteristics will be reported by the two groups under investigation.

***Specific Analyses***

***Overall analytic plan:*** Linear regression models will be built for each outcome variables of interest. The primary endpoint for these analyses will be the end of the randomized trial phase (Visit 10), while treating Visit 4 as the baseline. Unadjusted regression models will compare the difference between the two trial arms while controlling for the baseline value of the outcome measure. Adjusted models will evaluate the randomized treatment effect while adjusting for the covariates that were selected via backward elimination using a significance level of 0.05. These models will be built on data from subjects who completed the randomized nicotine reduction phase (Visit 10). We will conduct a separate statistical analysis for the subgroup of compliant participants.

Mixed effects regression models will also be used to study the change over the randomized nicotine reduction period of many longitudinal outcomes and to assess the difference of the trajectories between the randomized treatment arms. Models will be fit to evaluate the effects of time, group, and time-by-group interaction while adjusting for the baseline value (visit 4) for the corresponding outcome.

The hypotheses to be tested are as follows:
***Hypothesis 1.a.:*** Smokers assigned to the RNC group will have lower plasma cotinine concentrations during the last 3 weeks (visit 10) of the randomized phase of the study than those assigned to the UNC group.
***Hypothesis 1.b.:*** There will be no significant increase in key markers of tobacco use (e.g. cigarette consumption), biomarkers of tobacco smoke exposure (e.g. NNAL, CO), or health effects (e.g. blood pressure) in those assigned to the RNC group vs. those assigned to UNC.

***Hypothesis 2:*** There will be no significant increase in ratings of psychiatric or nicotine withdrawal symptoms in those assigned to the RNC group as compared to those assigned to UNC.

***Hypothesis 3:*** Smokers assigned to the RNC group will have lower perceived dependence, be more likely to report intention to quit smoking at the end of the randomized trial phase, and be more likely to achieve cigarette abstinence at Visit 12.
Both ITT abstinence (based on the assumption that a loss-to-follow-up subject resumes smoking) and completers' abstinence (based on the assumption that the probability of loss to follow up is independent of smoking status and abstinence can only be evaluated if the participant attended Visit 12) will be analyzed. Abstinence will be biochemically verified (CO < 10ppm).

A Kaplan-Meier survival analysis will be used to compare the time to dropout (from visit 4 date to date of dropping out of the randomized nicotine reduction phase) between the two treatment groups. Participants who completed the randomized nicotine reduction phase of the study (visit 10) will be considered as censored records at the time of visit 10. The date of dropout prior to Visit 10 is defined as the last scheduled contact date that a subject provides meaningful research data. The time to dropout will be compared between the two trial arms.

### 10.0 Confidentiality, Privacy and Data Management

#### 10.1 Confidentiality

The majority of study data at both sites will be collected and managed using REDCap (Research Electronic Data Capture). REDCap is a secure web application designed to support data capture for research studies, providing user-friendly web-based case report forms. REDCap is HIPAA compliant. Data are stored on a secure server at Hershey Medical Center and data in REDCap are encrypted. Access to the database requires authentication (a unique username and password) and a user matrix will be used to ensure that only appropriate data are accessed based on the individual's role on the project. Every interaction with the data is logged in REDCap creating an audit trail. Some participants may elect to complete one contact per month via a REDCap survey link that will be sent to their personal email account. No personal information will be sent through email as these emails will only contain a link to the participant's individual record in REDCap where they will be able to directly enter their information.

Paper files will be kept to a minimum and those that are generated will be stored in locked filing cabinets in the research offices of Dr. Jonathan Foulds.

Any electronic data generated (e.g. PFT reports) will be stored on the PHS computer network in password protected files.

##### 10.1.1 Identifiers associated with data and/or specimens

The following personal identifiers will be collected:

- Name
- Address
- Phone numbers
- Email addresses
- Date of birth
- Social security number

###### 10.1.1.1 Use of Codes, Master List

Study data will be collected in REDCap including a participant record number and a study specific code number. The list connecting the participant data to the code numbers will be stored electronically in REDCap and will not be destroyed. The PIs, Study Coordinators, Research assistants and Lab Managers will have access to the list.

##### 10.1.2 Storage of Data and/or Specimens

- **Electronic data** will be stored indefinitely in REDCap with identifiers attached. PFT reports will that are generated as a result of conducting the PFT test will be stored on the PHS computer network in password protected files. Data from these documents will be transferred into REDCap from the electronic file.
- **Paper records:**
  - Paper will be stored at the site where they are acquired for the length of the study in locked filing cabinets. Paper records will be scanned and uploaded into participant records in REDCap. Paper will be retained until the end of the study after which it will be destroyed. Electronic copies will be stored indefinitely as above.
- **Specimens:**
  - Specimens will be stored indefinitely with a code number attached.
  - Prior to processing, samples will be placed in a refrigerator by the study nurse, research assistant or study coordinator in the Clinical Research Center or in the key-card secured lab of Dr. Josh Muscat.
  - Within 24 hours, samples will be processed by a lab manager and will then be secured in a freezer located in a locked room within the research laboratory of Dr. Josh Muscat on the 3rd floor of the Cancer Institute.

##### 10.1.3 Access to Data and/or Specimens

**Study data in REDCap:** Primary study personnel including: Primary Investigators, Study Coordinators, Research Assistants, and Lab Managers will have access to the REDCap data. However, a REDCap user matrix will limit access to data based on the researcher's role in the study. These researchers will also have access to electronic data stored on the PHS files.

**Specimens:** Lab managers, technicians, researchers and the PIs will have access to the specimens once they have been processed and secured in the locked laboratory freezer room.

###### 10.1.4 Transferring Data and/or Specimens

Processed, frozen specimens from the Harvard study site will be shipped via commercial carrier to Dr. Muscat's Penn State laboratory for storage and analysis.

##### 10.2 Privacy

The research team will only have access to data that they have consented to provide and is provided by the participant during data collection contacts. HIPPA guidelines will be followed for all participants. Participants will be informed that they can refuse to answer any questions that make them feel uncomfortable. The majority of personal data that the participants provide will be entered directly into REDCap by the participant. All study visits, data collection and procedures will be completed in private consult rooms at the Penn State Hershey CTSI.

#### 11.0 Data and Safety Monitoring Plan

##### 11.1 Periodic evaluation of data

The study coordinator and the research nurse will be responsible for the daily oversight of subject safety. Patients will be under medical supervision while in the study and seen on an ongoing basis by our research staff who will assess adverse events and make appropriate referrals to the physician. Dr. Foulds and/or Dr. Hameed, will meet regularly with the study staff to review patient's progress and their experiences with the tobacco products, including any adverse events. Entrance criteria will be reviewed following screening. Medical history will be reviewed by Dr. Hameed or our nurse practitioner for any contraindications for the treatment products and vital signs will be checked at each in person visit.

Similar procedures will be instituted at Harvard University under supervision of Dr. Evins. Any adverse symptoms will be discussed over the telephone on our across site calls. Drs. Foulds and Evins will conduct internal monitoring of subject safety across all sites. In addition, the Data and Safety Monitoring Board will oversee the safety of the participants in the trial. The DSMB will receive summary reports on recruitment, retention, AEs, and CO every 6 months and will meet and produce a report and recommendation at least annually.

##### 11.2 Data that are reviewed

Data that will be reviewed include:

- Accrual and retention
- Adverse events and serious adverse events
- Protocol deviations/violations
- Misconduct
- Conflict of interest
- Participants' ability to achieve study requirements
- Significant changes in mental health status including suicidal ideation
- Changes in psychological measures of anxiety and depression (OASIS and QIDS)
- Changes in cigarette consumption from baseline
- Exhaled carbon monoxide increase from baseline

##### 11.3 Method of collection of safety information

All data, including safety data, will be coded directly into REDCap case report forms during study visits, over the phone or via online REDCap survey. Participant adverse events and serious adverse events will be assessed at each in-person study visit but can be reported at any time during the study.

##### 11.4 Frequency of data collection

Safety data including adverse events, serious adverse events and mental health assessments and carbon monoxide will be collected at each in-person visits. Cigarettes smoked per day will be collected regularly either at in person visits, over the phone or via online REDCap surveys.

##### 11.5 Individual's reviewing the data

The study coordinator and the research nurse will be responsible for the daily oversight of subject safety. Dr. Foulds and/or Dr. Hameed, will meet regularly with the study staff to review patient's progress and their experiences with the tobacco products, including any adverse events. Similar procedures will be instituted at Harvard/Mass General under supervision of Dr. Evins and Dr. Foulds.

##### 11.6 Frequency of review of cumulative data

Safety data will be reviewed cumulatively based on recommendations from the DSMB. They will receive summary reports every six months and convene at least annually.

##### 11.7 Statistical tests

Basic descriptive statistical methods will be used to analyze the safety data to determine whether harms are occurring. Changes in cigarette consumption from the baseline will be calculated. The exhaled carbon monoxide increase from baseline will be examined in the same way. In addition, the accrual and retention-dropout rate, completion rate, and the proportions of adverse events and serious adverse events will be generated. The data provided to the DSMB will not be separated by treatment group and will remain blinded. However, if the DSMB perceives a need to assess safety based on an unblinded analysis, they can request this information and it will be provided.

##### 11.8 Suspension of research

Due to the low risk of the intervention, it is unlikely that there will be a need to suspend the research. However, should the DSMB identify any issues after reviewing the data, they can develop stopping rules for the trial these recommendations will be followed.

#### 12.0 Risks

Potential risks for subjects are minimal. The cigarettes which will be administered to subjects have been previously tested and found to be of no greater risk than cigarettes the participants are already using. Only regular smokers who are not planning to quit or reduce their cigarette consumption will be recruited to the study. All participants will be free to reduce or quit smoking throughout the study, and smoking cessation treatment will be offered to all participants at the end of the randomized phase. Subjects will be under supervision throughout their participation in the study and adverse symptoms will be recorded at each in person clinic visit and monitored by the Project Leaders. The major side effects associated with RNC cigarettes are similar to usual brand cigarettes.

Additional potential risks include:

- **Increased compensatory smoking:** Compensatory smoking may lead to increased levels of toxicant exposure. In prior studies, compensatory smoking was minimal and higher levels of toxicant exposure were generally not observed. Cigarette consumption and exhaled carbon monoxide will be monitored throughout the trial. Safety rules are in place to identify and remove participants to increase their smoking rate significantly (see section 3.3.1).
- **Nicotine withdrawal symptoms:** Decreased nicotine cigarettes may result in nicotine withdrawal symptoms (e.g. irritability, anxiety, restlessness, depressed mood, increased appetite, fatigue, insomnia/sleep problems, impatience, headache, difficulty concentrating). These symptoms will be monitored regularly.
- **New pregnancy or intention to become pregnant:** Smoking is known to be harmful to the developing human fetus, either from cigarettes or at the recommended therapeutic dose of nicotine replacement therapy. For this reason women of child-bearing potential must agree to use adequate contraception (hormonal or barrier method of birth control; abstinence) prior to

study entry and for the standard duration of the study. Pregnancy status will be confirmed with a urinary pregnancy test every 4-8 weeks (see Time and Events Table).

- **Risks of standard venipuncture:** The discomfort associated with removing blood by venipuncture (by needle from a vein) is a slight pinch or pin prick when the sterile needle enters the skin. The risks include mild discomfort and/or a black and blue mark at the site of puncture. Less common risks include a small blood clot, infection or bleeding at the puncture site, and on rare occasions fainting during the procedure. There will be only a small amount of blood drawn (about 2 teaspoons) every three weeks.
- **Spirometry:** Risks associated with lung function tests spirometry may include shortness of breath, dizziness or headache while doing the breathing test. Participants with medical conditions that may place them at increased risk are being excluded.
- **Loss of confidentiality:** There is a risk of loss of confidentiality if information is obtained by someone other than the investigators. Precautions will be taken to prevent this including direct coding of data in REDCap.
- **Randomization in clinical trials:** Participants will be assigned to a treatment program by chance. The treatment they receive may prove to be less effective or to have more side effects than the other research treatment(s) or other available treatments.
- **Questionnaires:** It is possible that some of the questions in the questionnaires may make participants uncomfortable. They will be instructed that they are free to skip any questions that make them uncomfortable.
- **Use of oral nicotine replacement:** During the Treatment Choice Phase of the study, participants will be encouraged to quit smoking and will be offered a 11 week supply of oral nicotine replacement therapy (NRT) (lozenge or gum). NRT (lozenge or gum) is known to be safe and have only mild side effects. For this reason they are generally sold over the counter. Excess nicotine can cause mild symptoms such as nausea, dizziness, diarrhea and rapid heartbeat. Occasionally these (or other rarely occurring symptoms) are more severe. There is a slight risk that subjects who have progressed to very low nicotine cigarettes and then choose treatment with the NRT will obtain more nicotine than they are used to. A 24 hour period of cigarette and NRT abstinence will be recommended to the participants who want to quit. Participants will be seen after this waiting period and their craving and withdrawal will be assessed. Those who have very low or no withdrawal or craving will be offered the lowest dose and frequency of NRT. They may also choose to not use any NRT during their quit attempt. Those who elect to use NRT will be contacted regularly (see section 7.0 for study procedures during treatment choice phase) to monitor their tolerance.

  The use of oral nicotine replacement allows the participant to self-monitor their urges and cravings and use only the amount of NRT that they feel is necessary. A protocol to help the researcher determine the best dose of NRT to recommend to the participant will be used. Participants will be followed up within 48 hours of starting NRT to assess tolerance.

### 13.0 Potential Benefits to Subjects and Others

##### 13.1 Potential Benefits to Subjects

There are minimal benefits to study participants other than the (uncertain) possibility that participation in the study may reduce their nicotine dependence. Participants will receive financial compensation for their time and inconvenience. Those who complete the study will be offered help to quit smoking including evidence based cessation treatment with nicotine replacement therapy and counseling.

##### 13.2 Potential Benefits to Others

The main benefit to society and others from the study is a greater scientific understanding of the effect of switching to reduced nicotine content cigarettes.

### 14.0 Sharing Results with Subjects

This study is not designed to diagnose any disease or condition. However, if during the course of conducting clinical procedures (e.g., blood pressure, lung function), a participant is found to have a result outside of clinical norms, the result will be discussed with the participant at the visit where the result is identified (blood pressure, lung function). The participant will be given a letter indicating what procedure was done and will direct them to contact a medical provider for further evaluation. If a woman tests positive for pregnancy, the results will be shared with the participant and they will be advised to follow up with their doctor for prenatal medical care. They will not be allowed to participate in the study.

Abnormal medical results, adverse changes in mental health status, increased toxicant exposure (CO and cigarette consumption) and other outcomes of the study will be treated as Adverse Events. They will be discussed with the participant, documented and the results will be reviewed by the study PI.

At the end of the study, after all participants have completed the protocol, the investigators will send via email (if possible) or U.S. mail (if we do not have an email) all participants who completed the randomization visit (visit 4), a letter that lets them know to which study group they were randomized (see letter in study materials).

### 15.0 Economic Burden to Subjects

##### 15.1 Costs

There are no costs that subjects will be responsible for related to the research.

##### 15.2 Compensation for research-related injury

It is the policy of the institution to provide neither financial compensation nor free medical treatment for research-related injury. In the event of injury resulting from this research, medical treatment is available but will be provided at the usual charge. Costs for the treatment of research-related injuries will be charged to subjects or their insurance carriers.

### 16.0 Number of Subjects

A total of 280 participants will be enrolled in the study at two sites with the aim of randomizing 200 who complete the baseline phase. 100 participants will be enrolled in the randomized phase at Penn State Hershey. Participants will be started on the study protocol during Baseline I and Baseline II but will be removed from the study if they are not able to comply with the protocol. We expect that approximately 40 participants at each site drop out from the study prior to randomization (due to inability to comply with study protocol).

### 17.0 Resources Available

##### 17.1 Facilities and locations

All participant visits at Hershey will take place in the Penn State Hershey Clinical Research Center.

##### 17.2 Feasibility of recruiting the required number of subjects

The smoking prevalence in South Central Pennsylvania is 19% of the adult population and approximately 40% of these smokers have no current plan to reduce or quit smoking. Our recruitment strategy is designed to broadly disseminate information about the study to members of the community, and so we anticipate being able to recruit 140 over 3 years.

##### 17.3 PI Time devoted to conducting the research

Dr. Foulds has no clinical responsibilities and so the majority of his time is devoted to research, including this project. He is funded at 20% time for this study, and for additional time as co-PI of the overall Penn State Tobacco Center of Regulatory Science. The rest of the research team also have appropriate percent times covered by this grant.

**17.4 Availability of medical or psychological resources**

All of our participants will be seen by appropriately trained research staff or a Licensed Nurse Practitioner. Any serious AEs or concerning test results will be passed on to participants along with a letter to their doctor. Our protocol for assessing suicide risk requires the availability of a licensed psychiatrist to evaluate participants if they indicate any score on the MINI suicide $\geq$ 9 at screening or follow-up. Any score on the QIDS questionnaire > 0 will trigger the study staff to complete the MINI Suicide module. Scores $\geq$ 9 on the MINI will trigger a psychiatric assessment. Any urgent health problem will require accompanying the participant to the ER, which is located in the same building.

**17.5 Process for informing Study Team**

At least monthly team meetings will be conducted at PSU and Harvard/Mass General where study procedures, questions and issues will be discussed and resolved. In addition regular TCORS meetings will be held to discuss study progress. Offsite locations will have the ability to call in to these meetings.

### 18.0 Other Approvals

An Investigational Tobacco Product (ITP) application has been submitted to and approved by the FDA. A Certificate of Confidentiality has also been obtained.

### 19.0 Subject Stipend and/or Travel Reimbursements

Subjects will receive a $20 gift card per clinic visit to cover transportation costs for the 12 clinic visits ($240). In checks, subjects will receive $20 for completing each of the 12 clinic visits ($240), $10 for completing 6 survey or phone contacts ($60), and an additional $40 for the 9 visits when blood and urine samples are collected ($360). At the end of the study, participants will receive $50 complying with the cigarette pack protocol (accurate returned packs within 4 packs for 6/8 visits) and $50 if they attend all study visits (compliance incentive). The total payment for a subject who completes all study procedures will be $1,000. All payments will be made by check at Harvard/Mass General. Payments at each visit will be made according to the following schedule:

| Study Visit # | Transportation | Visit Completion | Blood/urine collection | Compliance incentive (attendance/ pack return) | Individual Visit Total | Overall Total |
|---|---|---|---|---|---|---|
| **1, 3 and 12** | $20 | $20 | | | $40 | $120 |
| **Survey or phone call (6 total)** | | $10 | | | $10 | $60 |
| **2, 4, 5, 6, 7, 8, 9, 10, 11** | $20 | $20 | $40 | | $80 | $720 |
| **12** | | | | $50/50 | $100 | $100 |
| **Study total** | | | | | | $1,000 |

### 20.0 Multi-Site Research

**20.1 Communication Plans**

The study team at both sites will meet at least monthly over the phone for the length of the trial to review study standard operating procedures, discuss challenges, accrual, AEs, resolve unique participant issues that occur during study visits and ensure compliance with the study protocol. The Hershey PI will participate in a site visit at Mass General prior to the start of the trial. The PI or other designated representatives will conduct periodic audits on at least an annual basis thereafter.

##### 20.2 Data Submission and Security Plan

The majority of study data will be directly coded into REDCap either by the research staff or at the participant's clinic visit. As a result, data security is embedded in the data collection plan. Paper copies of written consent will be periodically scanned and uploaded into the participant record in REDCap. Harvard/Mass General will retain their consents on paper and will not upload them into the REDCap project but they will be made available for periodic audits. Site audits will be conducted to ensure that paper data is secure. Electronic data checks will be conducted to ensure that electronic data is being entered correctly.

##### 20.3 Subject Enrollment

Screening and enrollment data will be directly coded into REDCap. Randomization will take place through the Cigarette Management System (CMS) which will be a web-based system available to the study sites (but the blind code will only be known to the Investigational Drug Service staff at Hershey, and the Data Management Core).

##### 20.4 Reporting of Adverse Events and New Information

All Adverse Event information (including serious AEs) will be coded directly into REDCap and will be monitored at each study site and by Drs. Foulds, Hameed and/or Evins on at least a monthly basis. Adverse events will be recorded directly into REDCap at each in-person participant visit. A summary report including analysis of AEs will be produced for the DSMB every 6 months and provided to the site PIs (without group breakdown), but only the DSMB will have the ability to examine patterns of AEs by group and to request an unblinded analysis if there is any cause for concern that needs to be clarified by an unblinded analysis.

##### 20.5 Audit and Monitoring Plans

The study team will meet at least monthly over the phone to discuss any issues that might arise with participants, screening, enrollment, and participant adherence to the study protocol or problems with implementing the protocol at each site. Data will be monitored through electronic data checks that will be executed in REDCap. The Penn State TCORS Data Management Core will conduct monthly data quality checks (missing data, data consistency across the same variable in different questions) and will inform the PIs at both sites of any irregularities that are detected.

#### 21.0 Adverse Event Reporting

##### 21.1 Adverse Event Definitions

| | |
|---|---|
| **Adverse event** | Any untoward medical occurrence associated with the use of the drug in humans, whether or not considered drug related |
| **Adverse reaction** | Any adverse event caused by a drug |
| **Suspected adverse reaction** | Any adverse event for which there is a reasonable possibility that the drug caused the adverse event. Suspected adverse reaction implies a lesser degree of certainty about causality than "adverse reaction".<br>• *Reasonable possibility.* For the purpose of IND safety reporting, "reasonable possibility" means there is evidence to suggest a causal relationship between the drug and the adverse event. |
| **Serious adverse event or Serious suspected adverse reaction** | Serious adverse event or Serious suspected adverse reaction: An adverse event or suspected adverse reaction that in the view of either the investigator or sponsor, it results in any of the following outcomes: Death, a life-threatening adverse event, inpatient hospitalization or prolongation of existing hospitalization, a persistent or significant incapacity or substantial disruption of the ability to conduct normal life functions, or a congenital anomaly/birth defect. Important medical events that may not result in death, be life-threatening, or require hospitalization may be considered serious when, based upon appropriate medical judgment, they may jeopardize the patient or subject and may require medical or |

| | |
|---|---|
| | surgical intervention to prevent one of the outcomes listed in this definition. |
| **Life-threatening adverse event or life-threatening suspected adverse reaction** | An adverse event or suspected adverse reaction is considered “life-threatening” if, in the view of either the Investigator (i.e., the study site principal investigator) or Sponsor, its occurrence places the patient or research subject at immediate risk of death. It does not include an adverse event or suspected adverse reaction that had it occurred in a more severe form, might have caused death. |
| **Unexpected adverse event or Unexpected suspected adverse reaction.** | An adverse event or suspected adverse reaction is considered “unexpected” if it is not listed in the investigator brochure, general investigational plan, clinical protocol, or elsewhere in the current IND application; or is not listed at the specificity or severity that has been previously observed and/or specified. |

#### 21.2 Recording of Adverse Events

Research subjects will be routinely questioned about adverse events at in person study visits.

All adverse events (serious or non-serious) and abnormal test findings observed or reported to study team believed to be associated with the study cigarettes will be followed until the event (or its sequelae) or the abnormal test finding resolves or stabilizes at a level acceptable to the investigator.

An abnormal test finding will be classified as an *adverse event* if one or more of the following criteria are met:

- The test finding is accompanied by clinical symptoms
- The test finding necessitates additional diagnostic evaluation(s) or medical/surgical intervention; including significant additional concomitant drug treatment or other therapy **Note:** Simply repeating a test finding, in the absence of any of the other listed criteria, does not constitute an adverse event.
- The test finding leads to a change in study drug dosing or discontinuation of subject participation in the clinical research study
- The test finding is considered an adverse event by the investigator.

#### 21.3 Causality and Severity Assessments

The investigator will promptly review documented adverse events and abnormal test findings to determine 1) if the abnormal test finding should be classified as an adverse event; 2) if there is a reasonable possibility that the adverse event was caused by the study drug(s) or device(s); and 3) if the adverse event meets the criteria for a *serious adverse event*.

If the investigator’s final determination of causality is “unknown and of questionable relationship to the study drug(s) or device(s)”, the adverse event will be classified as *associated with the use of the study cigarettes* for reporting purposes. If the investigator’s final determination of causality is “unknown but not related to the study cigarettes”, this determination and the rationale for the determination will be documented in the respective subject’s case history.

#### 21.4 Reporting of Adverse Reactions and Unanticipated Problems to the FDA

##### 21.4.1 Written IND Safety Reports

N/A

##### 21.4.2 Telephoned IND Safety Reports – Fatal or Life-threatening Suspected Adverse Reactions

N/A

#### 21.5 Reporting Adverse Reactions and Unanticipated Problems to the Responsible IRB

In accordance with applicable policies of The Pennsylvania State University Institutional Review Board (IRB), the investigator will report, to the IRB, any observed or reported harm (adverse event) experienced by a subject or other individual, which in the opinion of the investigator is determined to be (1) unexpected; and (2) probably related to the research procedures. Harms (adverse events) will be submitted to the IRB in accordance with the IRB policies and procedures. All product related adverse events of a non-serious nature will be reported to the IRB at the time of renewal.

Any reportable adverse events that occur at Harvard/Mass General will first be reported to their IRB. The subsequent IRB determination report from Harvard/Mass General will then be submitted to the Penn State Hershey IRB.

All serious adverse events that occur at either site (regardless of causality) will be submitted to the FDA within 5 business days.

The Data and Safety Monitoring Board will review all serious adverse events and provide recommendations. We will inform NIH and FDA of any significant action taken as a result of the Data and Safety Monitoring Board's findings. We will inform the subjects of any changes in risk.

##### 21.6 Unblinding Procedures

If an adverse event requires the subject to be unblinded, the unblinded study personnel (cigarette administrator) will be able to provide that information as needed. Otherwise, participants will not be unblinded to their cigarette allocation. The DSMB will begin by reviewing the protocol and establishing guidelines for data and safety monitoring including any additional procedures for unblinding of participants.

##### 21.7 Stopping Rules

The DSMB will also develop trial stopping rules to help define the point at which the risks to subjects of continuing the study outweigh the likely benefits. A brief report will be generated from each of these meetings for the study record and forwarded to the each of the study site's Institute's Institutional Review Boards (IRB).

#### 22.0 Study Monitoring, Auditing and Inspecting

##### 22.1 Study Monitoring Plan

###### 22.1.1 Quality Assurance and Quality Control

Data will be collected from participants and coded directly by either using the REDCap survey tool (participant entered data) or through REDCap data entry forms (researcher entered data). The codes that link the name of the participant and the study ID will be kept confidential in REDCap. Any paper forms (consent) will be securely transported to the PI's data entry center. Any additional data that is generated (i.e., initial electronic PFT test reports), will be stored electronically on the PHS server in password protected files.

Study data will be managed using REDCap (Research Electronic Data Capture). REDCap is a secure web application designed to support data capture for research studies, providing user-friendly web-based case report forms, real-time data entry validation (e.g. for data types and range checks), audit trails and a de-identified data export mechanism to common statistical packages (SPSS, SAS, Stata, R/S-Plus). The system was developed by a multi-institutional consortium which includes The Pennsylvania State University and was initiated at Vanderbilt University. The database is hosted at the Penn State Hershey Medical Center and College of Medicine data center, which will be used as a central location for data processing and management. REDCap data collection projects rely on a thorough study-specific data dictionary defined in an iterative self-documenting process by all members of the research team. This iterative development and testing process results in a well-planned data collection strategy for individual studies.

REDCap is HIPAA compliant. Data are stored on a secure server; data in REDCap are encrypted; access to the database requires authentication (a unique username and password); data are accessed based on the individual's role on the project; every interaction with the data is logged, creating an audit trail.

Random data entry checks will be implemented regularly to identify problems with data entry. Data quality tools included in REDCap will be utilized to identify incorrect data types, out of range data and outliers. In addition, electronic edit checks, and random internal quality and assurance checking will be performed manually. Data quality will be monitored by random inspection of the completed electronic forms by one of the research assistants and any problems detected will be discussed with the PI. If necessary, re-training of researchers will be conducted.

The responsibility for data quality and study conduct lies with the PI. Site visits will be conducted by the Penn State research team to ensure that procedures at both sites are being executed in compliance with the protocol, and IRB policies.

###### 22.1.2 Safety Monitoring

The DSMB will monitor the safety of study participants. The Principal Investigator will confirm that all adverse events (AE) are correctly entered into the AE case report forms by the coordinator; be available to answer any questions that the coordinators may have concerning AEs; and will notify the IRB, FDA, sponsor and/or DSMB of all applicable AEs as appropriate. All final assessments of AEs will be made by a licensed medical professional who is an investigator on the research.

The research coordinator will ensure that AEs are correctly entered into REDCap and complete the appropriate report form and logs; assist the PI to prepare reports and notify the IRB, FDA, and/or DSMB of all Unanticipated Problems/SAE's.

##### 22.2 Auditing and Inspecting

The investigator will permit study-related monitoring, audits, and inspections by the Penn State quality assurance program office(s), IRB, the sponsor, and government regulatory bodies, of all study related documents (e.g. source documents, regulatory documents, data collection instruments, study data etc.). The investigator will ensure the capability for inspections of applicable study-related facilities (e.g. pharmacy, diagnostic laboratory, etc.).
