## Supplement 2 for "The effects of reduced nicotine content cigarettes in smokers with mood or anxiety disorders: a double-blind randomized trial"

[§]Corresponding author

### Data Safety Monitoring Board Members

1. John Hughes MD, University of Vermont (Chair)
2. Michael Steinberg MD MPH, Robert Wood Johnson Medical School -Rutgers University,
3. Dave Mauger PhD, Penn State – College of Medicine

### Supplementary Methods

#### Author Contributions

Dr. Foulds wrote the first draft of the paper and circulated to all authors for comments. Drs. Liao, Zhu and Emily Wasserman conducted all analyses in consultation with Dr. Foulds and the other authors. All authors participated in designing the study and collecting the data. All authors share in the decision to publish the paper and in the responsibility for the manuscript as submitted.

#### Study Protocol and Statistical Analysis Plan

A complete study protocol, statistical analysis plan and summary of changes made after initiation of the study is provided in Supplementary Appendix 2.

#### Biomarker Laboratory Methods

#### Blood Biomarkers

Blood samples were collected and stored at -80 $^{o}$ to measure plasma cotinine, glutathione redox status, expressed as GSSP/GSH ratio, and hemoglobin. Levels of free (reduced) GSH and protein-bound GSH (GSSP) in blood were determined as described previously (1, 2). Cotinine concentration levels in plasma were measured using a solid-phase, enzyme-linked immunosorbent assay (ELISA) kit from Calbiotech (El Cajon, CA)(1). Briefly, standards, controls, and diluted plasma samples were added in duplicate to the wells coated with a polyclonal antibody to cotinine. After addition of the enzyme conjugate, the plates were incubated in the dark for one hour at room temperature. All wells were thoroughly washed, a substrate reagent was added, and the plates were incubated for an additional 20 minutes in the dark. After addition of a stop solution, the absorbance was measured at 450 nm. The concentration of cotinine (ng/mL) was calculated against the standard curve generated from the standards supplied in the kit.

**Urine Biomarkers**

Urine samples were collected and stored at -80 °C for 4-(methylnitrosamino)-1-(3-pyridyl)-1-butanol (NNAL), 1-hydroxypyrene (1-HOP), and 8-isoprostane analyses and values were standardized by creatinine to account for urinary dilution. Urinary creatinine was determined spectrophotometrically based on the reaction of creatinine with alkaline(3). Briefly, 50 µL of diluted urine were mixed with 200 µL of 0.12% of picric acid in 0.15 N sodium hydroxide and, after 30 minutes, absorbance at 490 nm was measured using a microplate reader (Biotek Synergy HTX). Creatinine levels were quantified relative to a creatinine standard curve.

Urine NNAL samples were thawed at room temperature, and centrifuged for 1 min at 106 rcf. To a 500 µl aliquot, 30 µl of $d_5$-NNAL internal standard in water, (Toronto Research Chemicals, 100 ng/ml), was added. The samples were then incubated overnight at 37 °C with 70 µl (600 units) of beta-glucuronidase (from E. coli, Sigma) in potassium phosphate buffer (pH6.8). After incubation the samples were cooled on ice, following which 48 µl of 0.1 N potassium hydroxide was added. The samples were then extracted twice with 1 ml of chloroform. The organic and aqueous layers were separated by centrifugation at 10,600 rcf for 5 minutes. The resulting chloroform extracts were combined and evaporated to dryness using a Savant RVT4104 SpeedVac. The samples were reconstituted in 200 µl of water, filtered using Millipore Ultrafree-MC-HV centrifuge filters (6790 rcf for 5 min), then transferred to inserts for LC/MS analysis.

LC/MS/MS analyses were carried out by positive ion electrospray on an AB Sciex 4500 Q-trap mass spectrometer (AB Sciex, Redwood Shores, CA) in line with a an Agilent 1100 HPLC system (Agilent Technologies, Santa Clara, CA). Chromatographic separation of NNAL was accomplished using an Agilent Zorbax C8 reverse phase column (5 micron, 250 mm x 4.6 mm), employing the following program at a flow rate of 0.8 ml/min: 5 min at initial conditions of 90% water (solvent A) and 10% acetonitrile (solvent B), before running a gradient to 40% solvent B in 15 minutes. The column was washed for 5 minutes at 95% solvent B before returning to initial conditions.

NNAL was analyzed employing multiple reaction monitoring of two ion transitions: *m/*z 210 to *m/*z 93 and *m/*z 210 to *m/*z 180 for NNAL and *m/*z 215 to *m/z* 93 and *m/*z 215 to *m/*z 185 for the internal standard. Interfering peaks were seen in a number of samples for the *m/*z 210 to 93 transition. Therefore NNAL was quantified using the *m/*z 210 to 180 transition with confirmation provided by the *m/*z 210 to 93 transition. The quadrupole was calibrated to unit resolution with a scan width of 0.6 to 0.8 and peak width half height. The scan time was 0.63 seconds.

1-HOP samples were thawed at room temperature, and centrifuged for 1 min at 106 rcf. Prior to adding the sample, 20µl of internal standard, 129.36ng/ml of $d_9$ 1-hydroxypyrene in acetonitrile (Toronto Research Chemicals), was evaporated to dryness using a Savant RVT4104 SpeedVac. To each tube was then added 980 µl of urine. Following vortex mixing, 1000 units of beta-glucuronidase (from E. coli, Sigma) in water was added. The samples were incubated overnight at 37ºC, then cooled on ice and extracted twice with 1 ml of chloroform. The organic and aqueous layers were separated by centrifugation at 10,600 rcf for 5 minutes. The chloroform extracts were combined and evaporated to dryness on the Savant SpeedVac. The samples were reconstituted in 100 to 150 µl of methanol, filtered using Millipore Ultrafree-MC-HV centrifuge filters (6790rcf for 5min), then transferred to inserts for LC/MS/MS analysis.

LC/MS/MS analyses were carried out by negative ion electrospray on an AB Sciex 4500 Q-trap mass spectrometer (AB Sciex, Redwood Shores, CA) in line with a an Agilent 1100 HPLC system (Agilent Technologies, Santa Clara, CA). Chromatographic separation of 1-HOP was accomplished using an Agilent Zorbax C8 reverse phase column (5 micron, 250 mm x 4.6 mm), using the following program at a flow rate of 0.8 ml/minutes: initial conditions of 95% water (solvent A) and 5% methanol (solvent B), run to 95% solvent B in 36 minutes. The column was washed for 5 minutes at 95% solvent B before returning to initial conditions.

1-HOP was analyzed employing multiple reaction monitoring of two ion transitions: *m/z* 217 to *m/z* 217 and *m/z* 217 to *m/z* 189 for 1-HOP and *m/z* 226 to *m/z* 226 and *m/z* 226 to *m/z* 198 for the internal standard. 1-HOP was quantified using the *m/z* 217 to *m/z* 217 transition with confirmation provided by the *m/z* 217 to *m/z* 189 transition. The vaporizer temperature was 600ºC and the ion spray voltage was –4000V. The quadrupole was calibrated to unit resolution with a scan width of 0.6 to 0.8 at peak width half height. The scan time was 0.42 seconds.

Urinary 8-isoprostane levels were measured using a commercially available competitive enzyme-linked immunoassay (ELISA) kit from Oxford Biomedical Research (Oxford, MI, USA). The manufacturer's protocol was followed accordingly. Briefly, urine samples and controls were incubated for 2 hours at 37°C with β-glucuronidase. Standards, diluted samples, and diluted controls were added in duplicate to wells coated with a polyclonal antibody specific to 15-F2t-Isoprostane. After addition of the enzyme conjugate, the plates were incubated for two hours at room temperature. All wells were thoroughly washed, substrate reagent was added, and the plates were incubated for an additional 30 minutes. The reaction was stopped with sulfuric acid and absorbance was measured at 450 nm. The concentration of 8-isoprostane (ng/mL) was calculated against a standard curve generated from the standards supplied in the kit.

**Rationale for Statistical Analysis Approach**

The aim of this trial is to provide information to the FDA about the likely effects of a regulatory strategy involving a product standard for all cigarettes that limits the permissible nicotine content in cigarettes to a very low level, that is minimally or non-addictive, on smokers with a history of mood and/or anxiety disorders. Considering the possibility that switching smokers with mood and/or anxiety disorders abruptly to cigarettes of very low nicotine levels could cause adverse effects (e.g. nicotine withdrawal causing exacerbation of existing or dormant mood or anxiety disorder, or smokers compensating for the low nicotine levels by inhaling more smoke and toxicants), we chose to acclimate the participants to the lowest nicotine content dose in a gradual, step-wise manner. Those who reached the lowest nicotine content dose remained on this level for a longer period of time than the other step-wise doses (6 weeks rather than 3 weeks) to allow these smokers to adjust fully to these very low nicotine content (VLNC) cigarettes. With this design in mind, the trial was primarily meant to assess the effect of smoking VLNC cigarettes, as compared with regular nicotine cigarettes. As published in our protocol paper (4) and original submitted protocol, we anticipated approximately 30% dropout and powered the trial under the assumption that 70 participants would complete the randomized phase to visit 10 in each group. At this time in the trial, the RNC group would have smoked VLNC cigarettes for 6 weeks. In a non-experimental environment, the potential FDA regulation would continue indefinitely into the future, therefore, we consider the outcomes assessed at the end of the randomized phase (V10 - after 6 weeks on the VLNCs) to be the best experimental estimate of these effects. It is for this reason that our statistical analyses primarily focus on the outcomes and measurements at visit 10, and therefore necessarily highlights the subgroup of randomized phase completers.

We followed the strategy for intention to treat analysis in randomized trials with missing outcome data (4) as follows:

1. Our main analyses included all randomized participants who provided data at our main outcome point (visit 10), regardless of compliance. By design, this subgroup is characterized by randomized phase completers.
2. To explore whether the intervention had an effect on the randomized phase completion rate (synonymously, the dropout rate), a time-to-dropout analysis assessed the study arm effect. The randomized phase dropout did not differ by the intervention.
3. Noting these findings, we view the compliers subgroup analysis as a type of sensitivity analysis. This subgroup is assumed to represent the maximal effect.
4. To further analyze all randomized participants without subgroups, linear mixed effects models were fit for select primary and secondary outcome measures: Plasma Cotinine, Total CPD, Exhaled CO, MNWS, QIDS, OASIS, PSS, and K6. These models include all randomized participants and assess the differences in trajectories between the study arms throughout the randomized phase of the trial.

### Supplementary Results

Figure S1. CONSORT diagram (for full details by group after randomization see Figure 1)

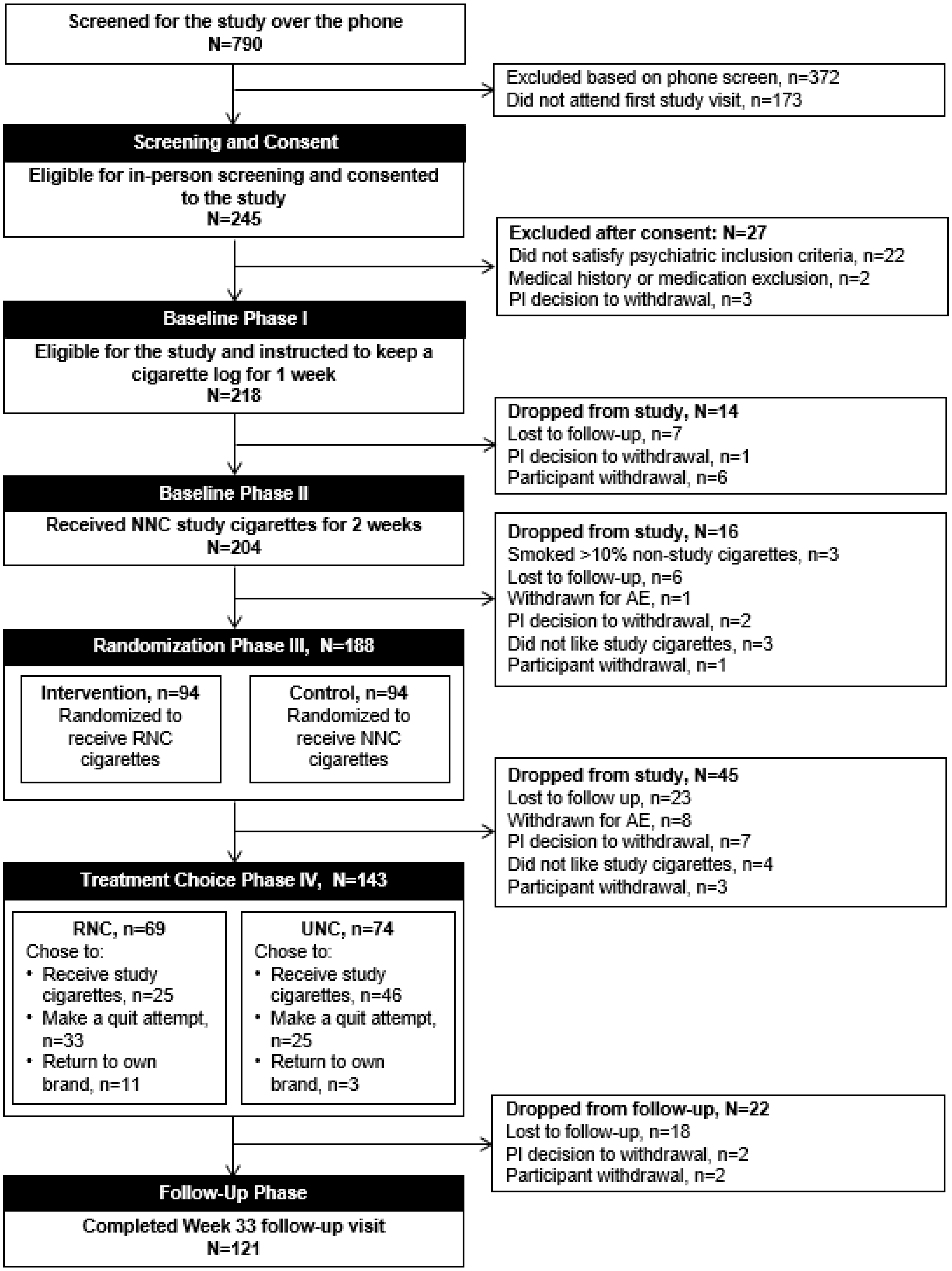

Figure S2. Study design flow diagram

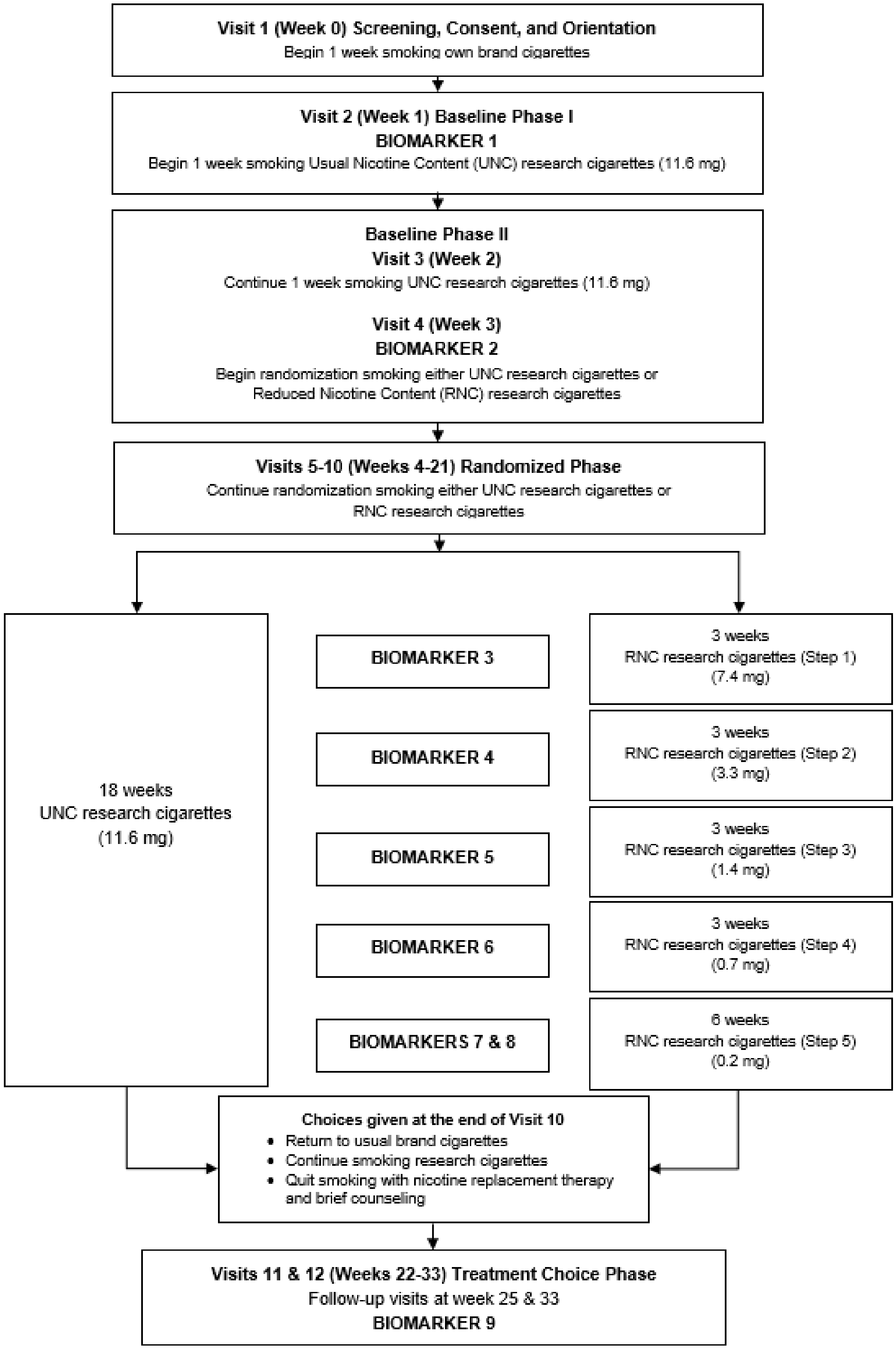

**Main Outcome Figures**

Three figures based on sample subgroups for each outcome measure (raw observed means):

1. Randomized Phase Completers
2. Randomized Phase Compliers
3. All Randomized Participants (For the following outcomes measures:: Plasma Cotinine, CPD, CO, MNWS, QIDS, OASIS, PSS, K6)

   * indicates a significant difference ($p<0.05$) between the two groups at Visit 10 after adjusting for the baseline measure at Visit 4 (Manuscript Table 2).

Plasma Cotinine (ng/mL)

Figure S3. Randomized Phase Completers (UNC, n=74; RNC, n=69)

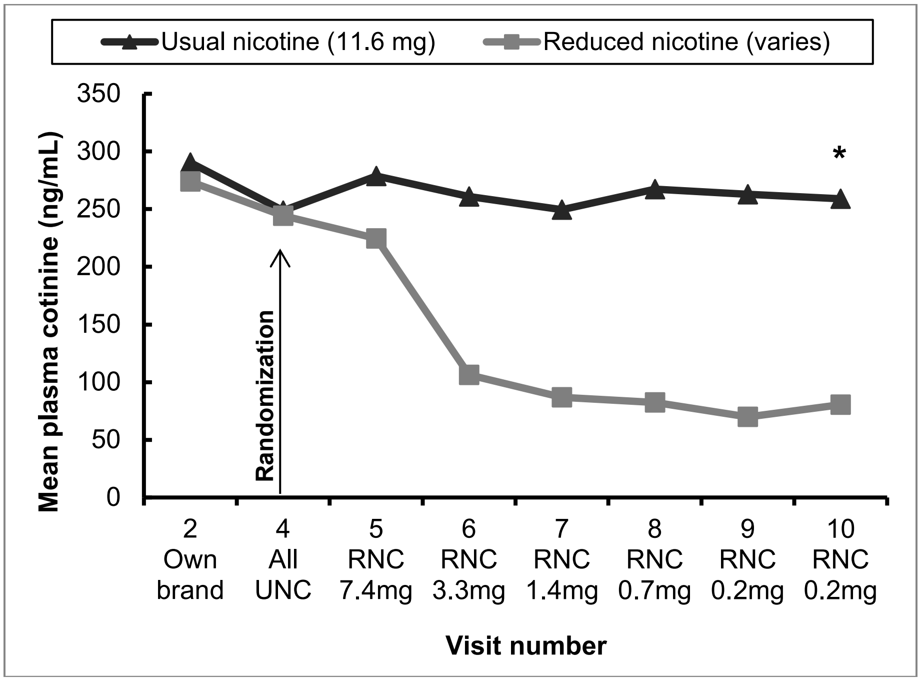

Figure S4. Randomized Phase Compliers (UNC, n=62; RNC, n=41)

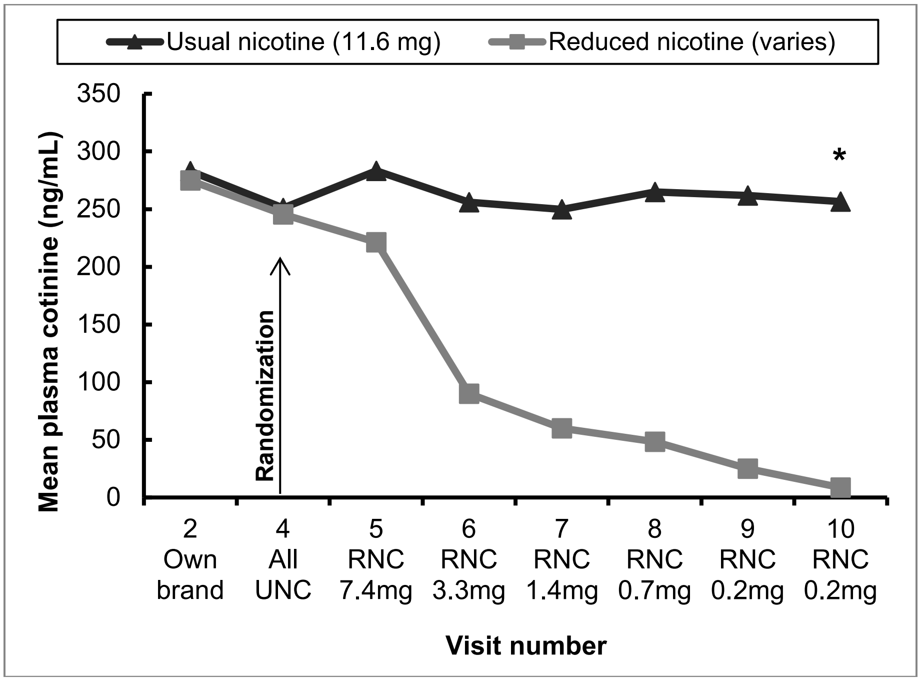

Plasma Cotinine (ng/mL)

Figure S5. All Randomized Participants (UNG n=94, RNG n=94)

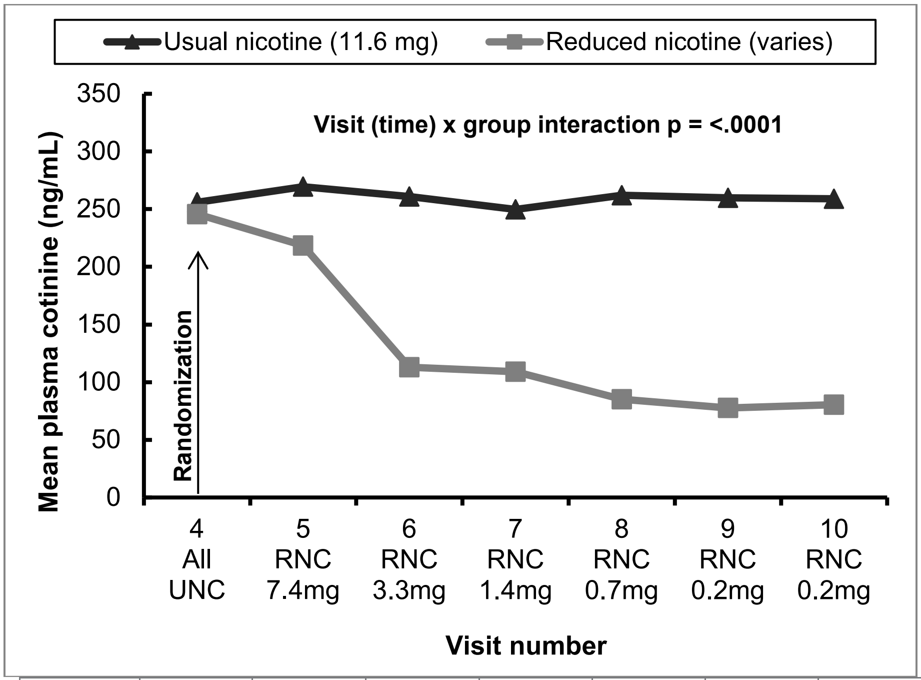

| | V4 | V5 | V6 | V7 | V8 | V9 | V10 |
|---|---|---|---|---|---|---|---|
| UNG | 93 | 84 | 81 | 81 | 81 | 76 | 72 |
| RNG | 91 | 86 | 84 | 80 | 73 | 69 | 68 |

Total Cigarettes per Day

Figure S6. Randomized Phase Completers (UNC, n=74; RNC, n=69)

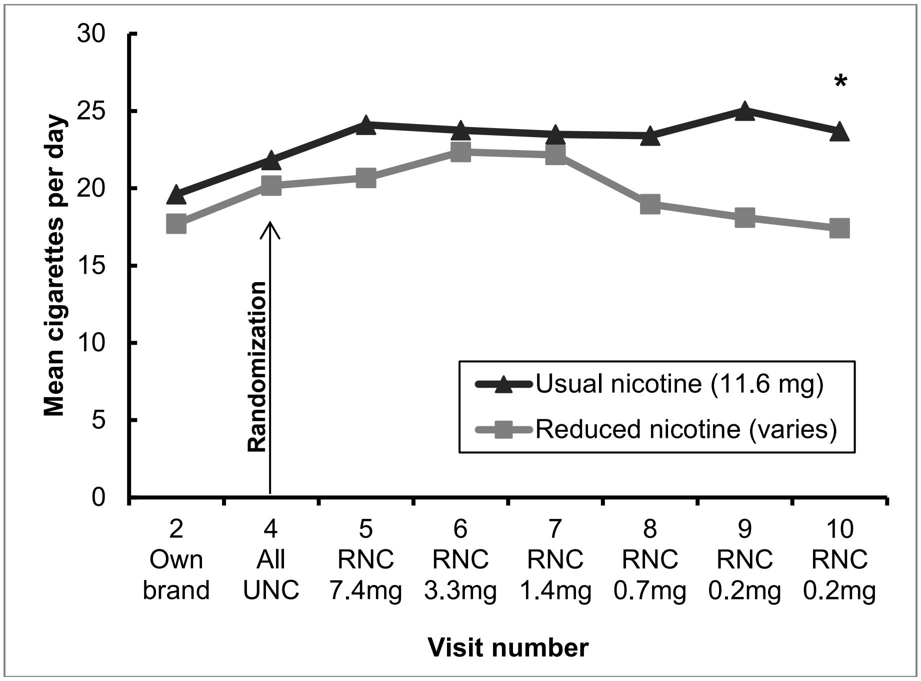

Figure S7. Randomized Phase Compliers (UNC, n=62; RNC, n=41)

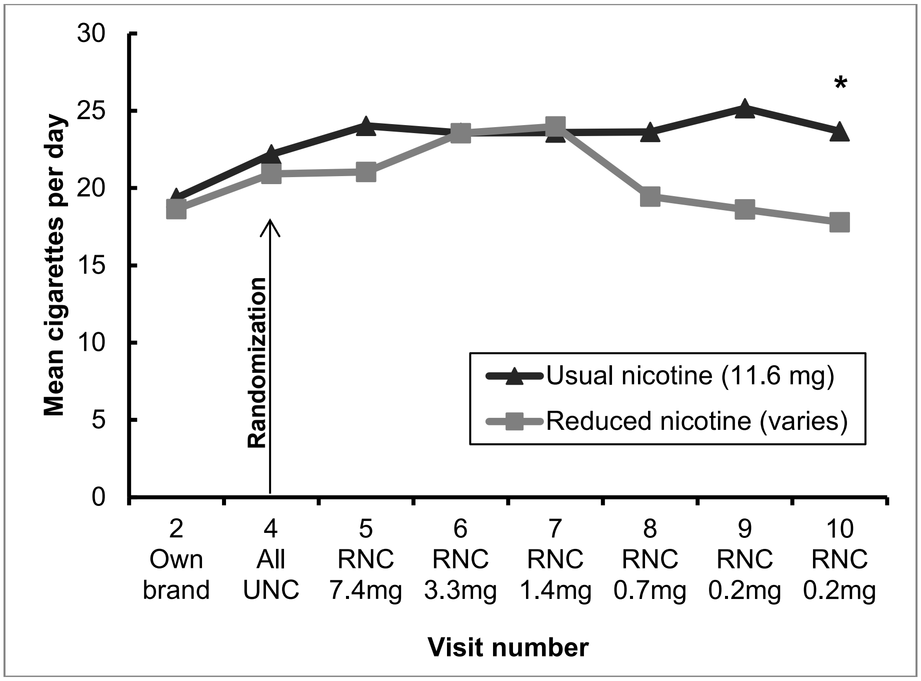

Total Cigarettes per Day

Figure S8. All Randomized Participants (UNG n=94, RNG n=94)

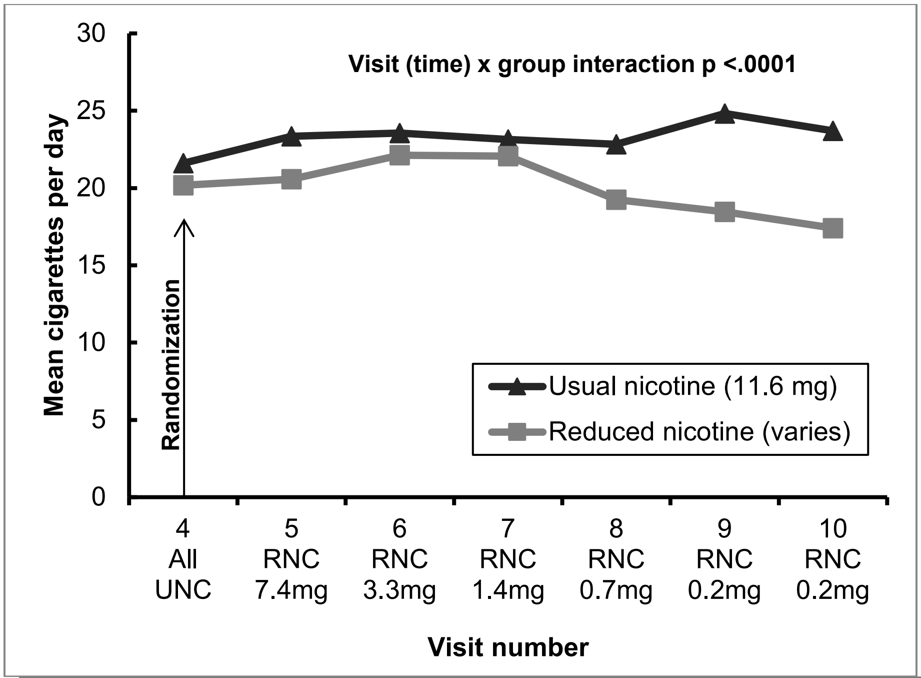

| | V4 | V5 | V6 | V7 | V8 | V9 | V10 |
|---|---|---|---|---|---|---|---|
| UNG | 94 | 89 | 84 | 83 | 81 | 77 | 74 |
| RNG | 94 | 90 | 87 | 80 | 72 | 72 | 69 |

Carbon Monoxide (Exhaled Breath Concentration, parts per million)

Figure S9. Randomized Phase Completers (UNC, n=74; RNC, n=69)

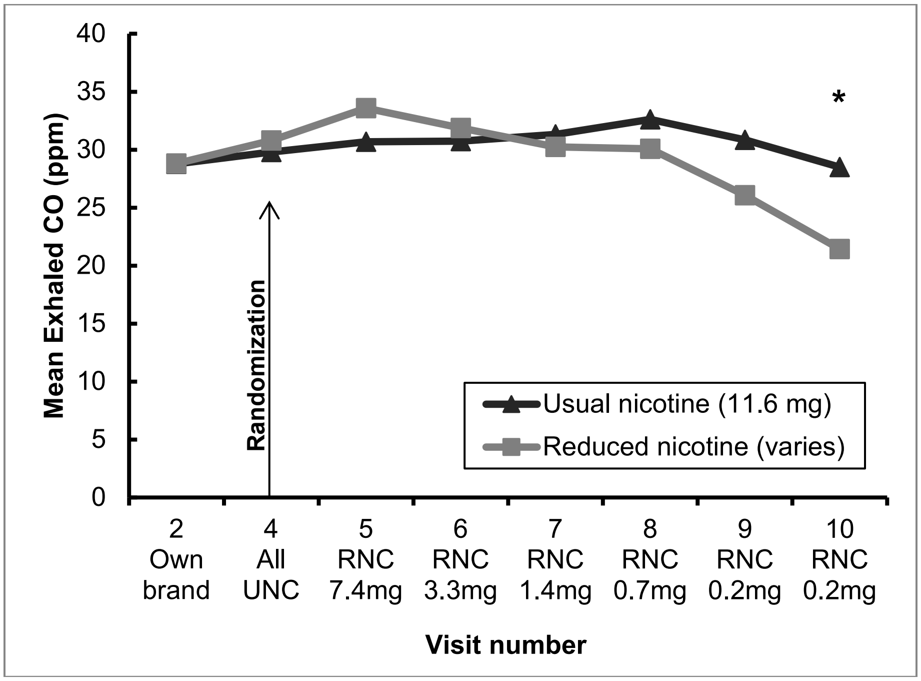

Figure S10. Randomized Phase Compliers (UNC, n=62; RNC, n=41)

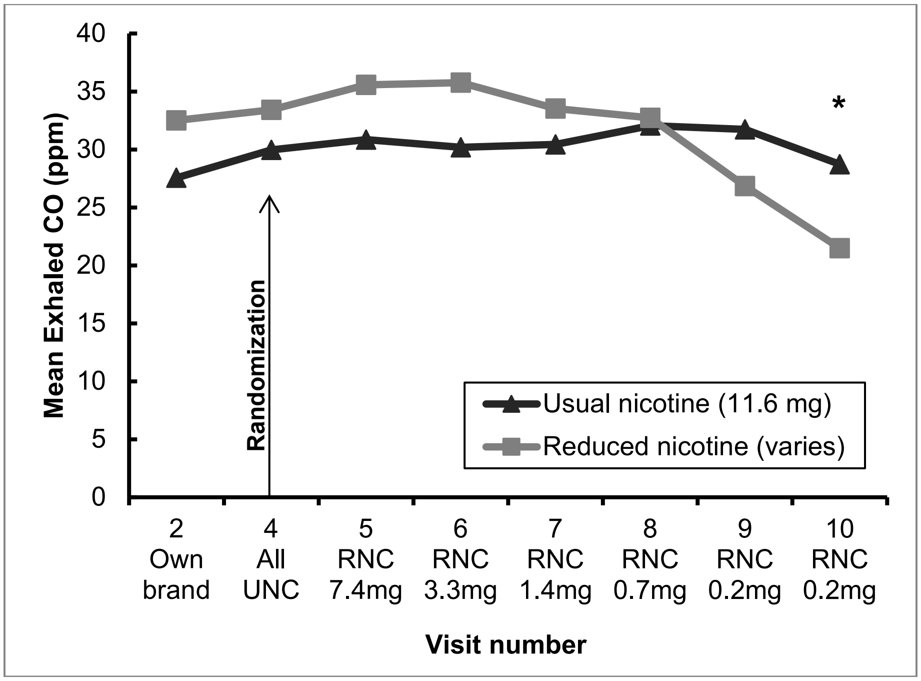

Carbon Monoxide (Exhaled Breath concentration, parts per million)

Figure S11. All Randomized Participants (UNG n=94, RNG n=94)

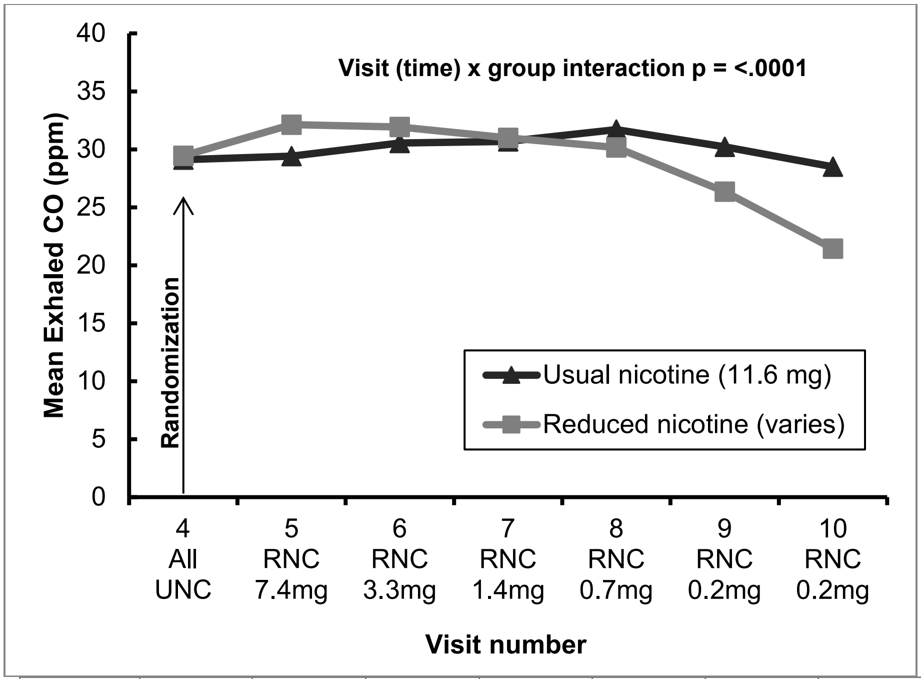

| | V4 | V5 | V6 | V7 | V8 | V9 | V10 |
|---|---|---|---|---|---|---|---|
| UNG | 94 | 87 | 84 | 83 | 81 | 77 | 73 |
| RNG | 94 | 90 | 87 | 80 | 75 | 72 | 69 |

FTND (Fagerström Test of Nicotine Dependence – Range: 0-10)

Figure S12. Randomized Phase Completers (UNC, n=74; RNC, n=69)

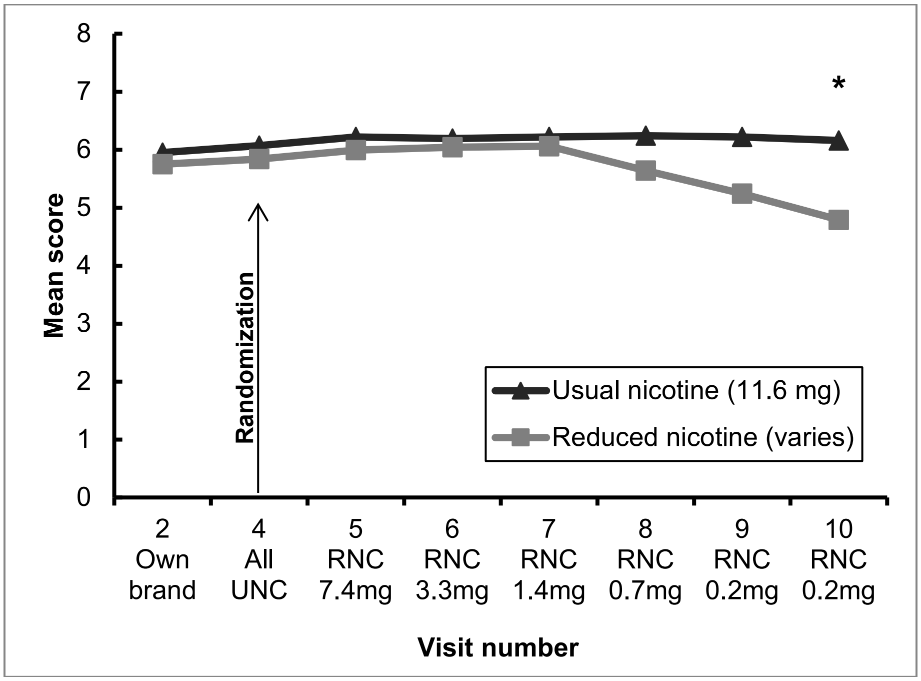

Figure S13. Randomized Phase Compliers (UNC, n=62; RNC, n=41)

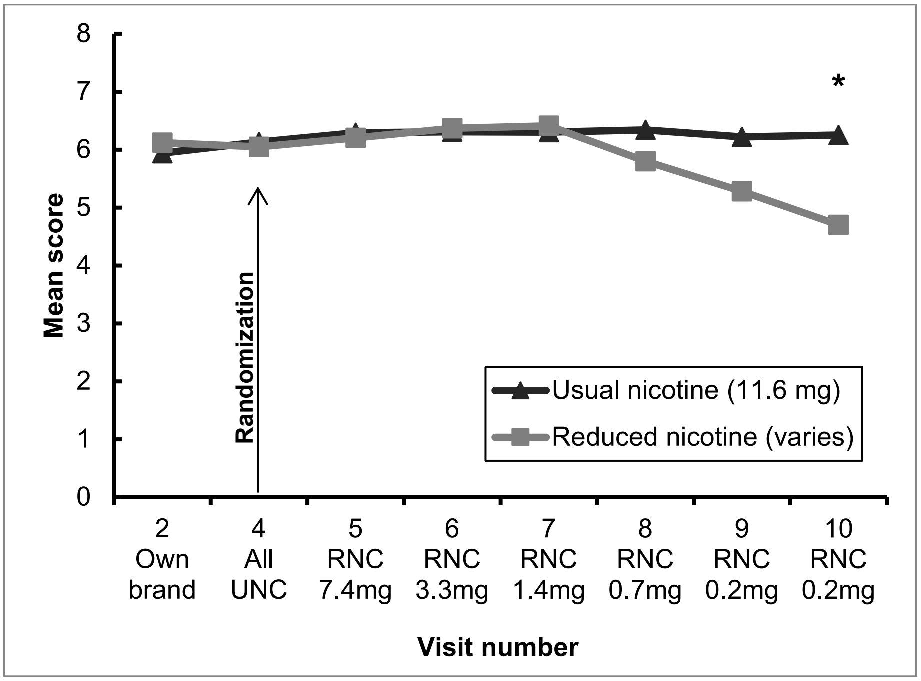

PSCDI (Penn State Cigarette Dependence Index – Range: 0-20)

Figure S14. Randomized Phase Completers (UNC, n=74; RNC, n=69)

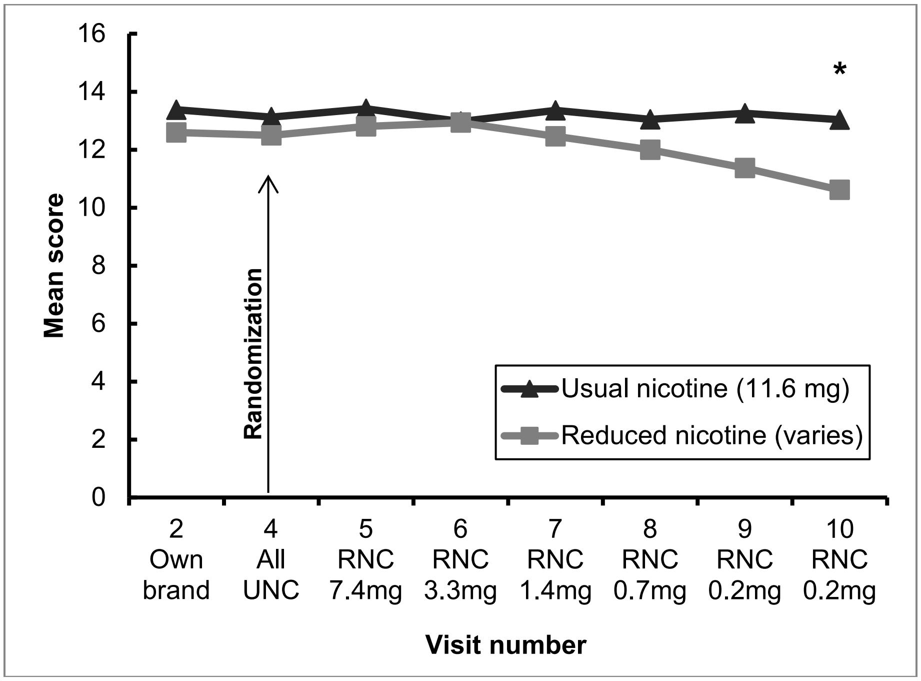

Figure S15. Randomized Phase Compliers (UNC, n=62; RNC, n=41)

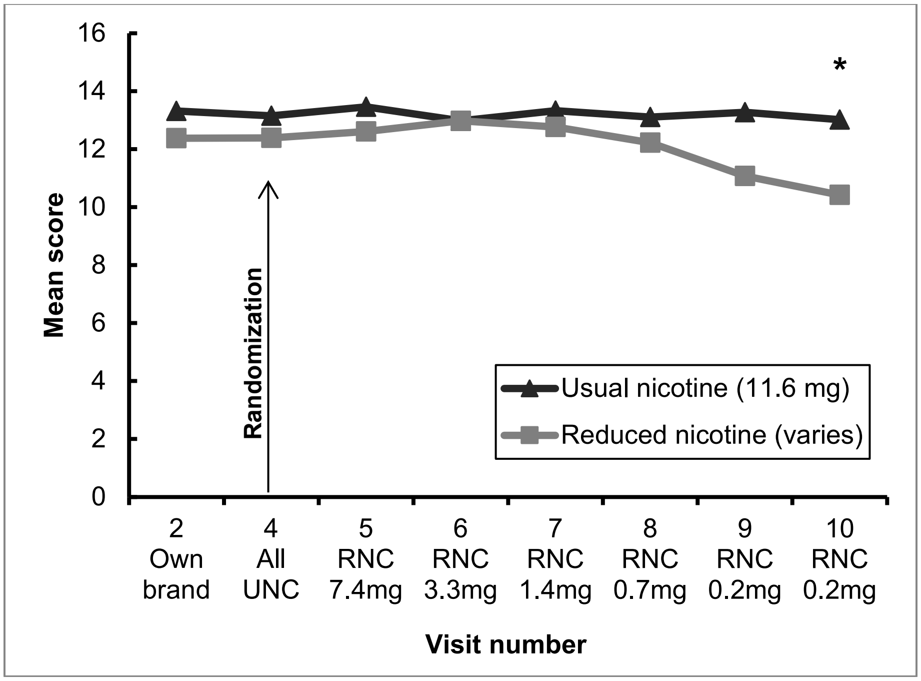

QSU Brief Total Score (Questionnaire on Smoking Urges – Range: 10-70)

Figure S16. Randomized Phase Completers (UNC, n=74; RNC, n=69)

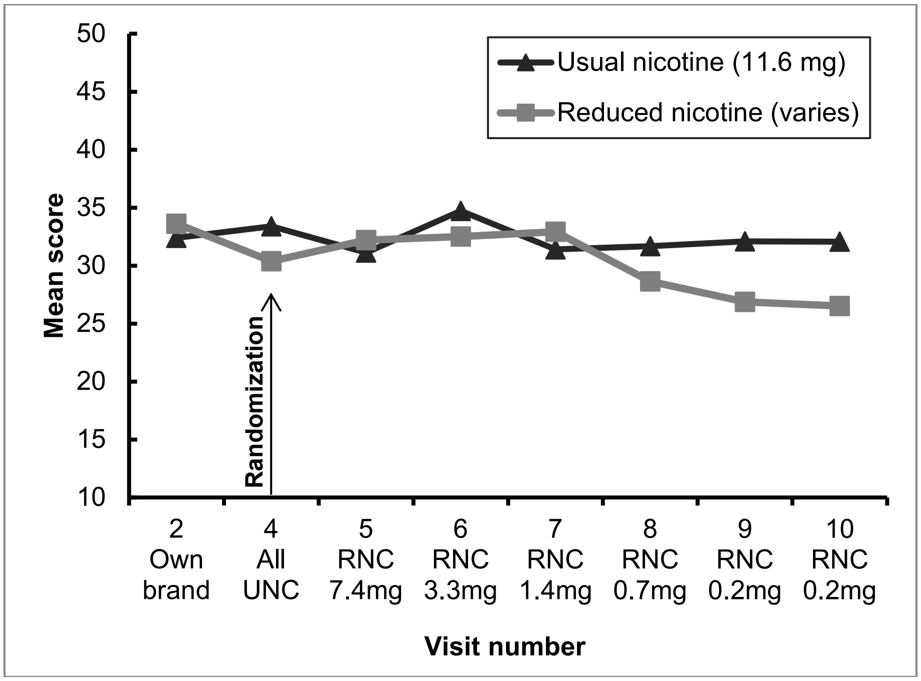

Figure S17. Randomized Phase Compliers (UNC, n=62; RNC, n=41)

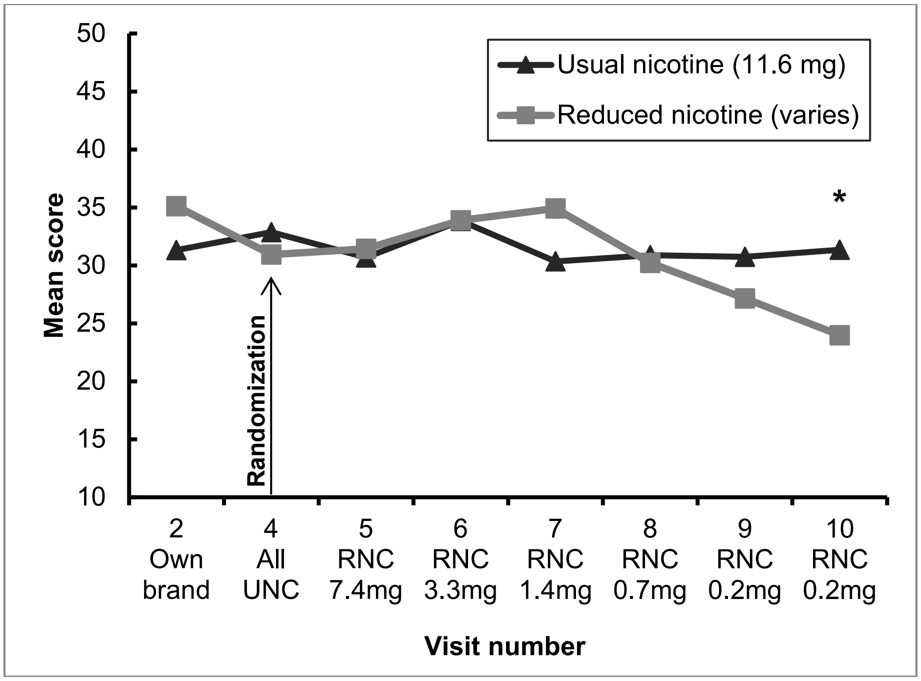

MNWS (Minnesota Nicotine Withdrawal Scale – Range: 0-32)

Figure S18. Randomized Phase Completers (UNC, n=74; RNC, n=69)

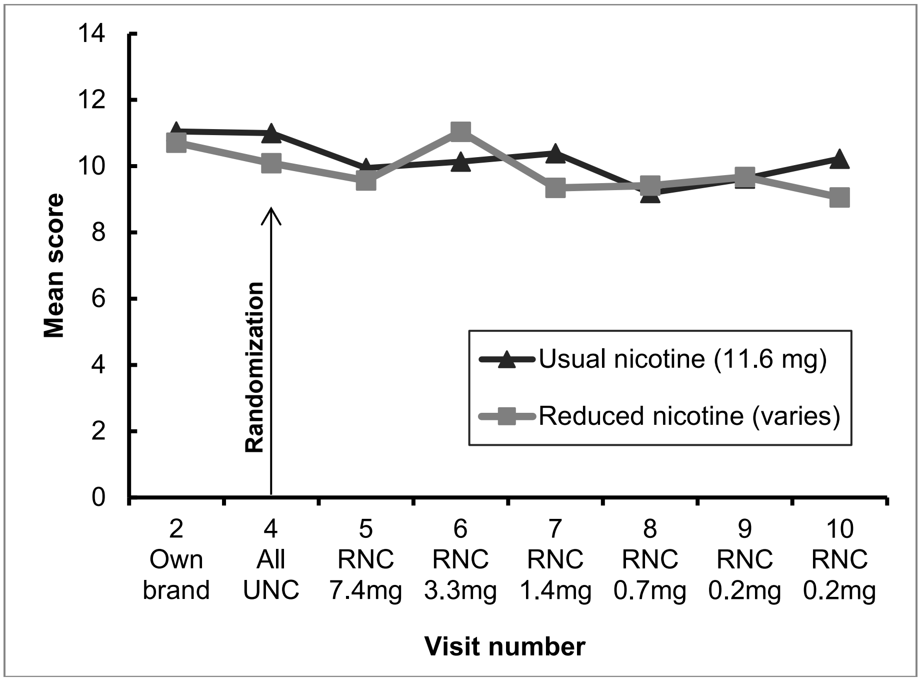

Figure S19. Randomized Phase Compliers (UNC, n=62; RNC, n=41)

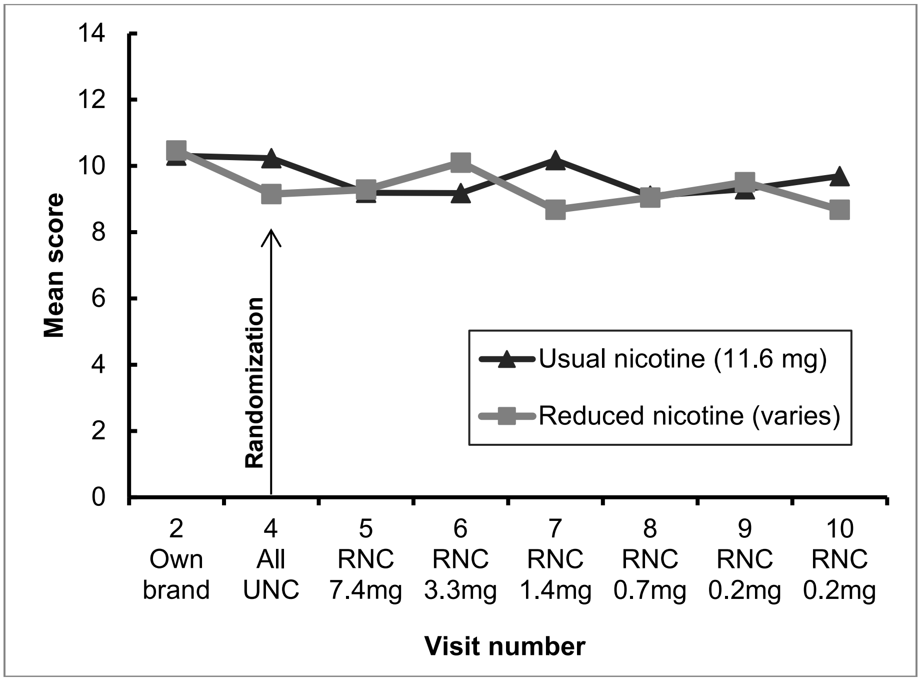

MNWS (Minnesota Nicotine Withdrawal Scale – Range: 0-32)

Figure S20. All Randomized Participants (UNG n=94, RNG n=94)

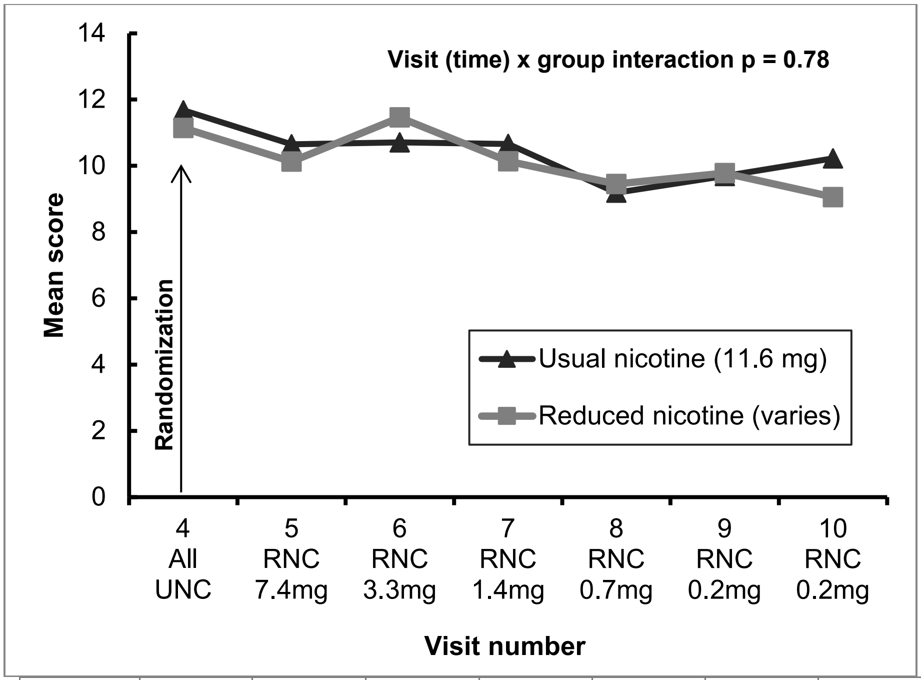

| | V4 | V5 | V6 | V7 | V8 | V9 | V10 |
|---|---|---|---|---|---|---|---|
| UNG | 94 | 88 | 84 | 83 | 81 | 77 | 74 |
| RNG | 94 | 90 | 87 | 81 | 75 | 72 | 69 |

QIDS (Quick Inventory of Depressive Symptomatology – Range: 0-27)

Figure S21. Randomized Phase Completers (UNC, n=74; RNC, n=69)

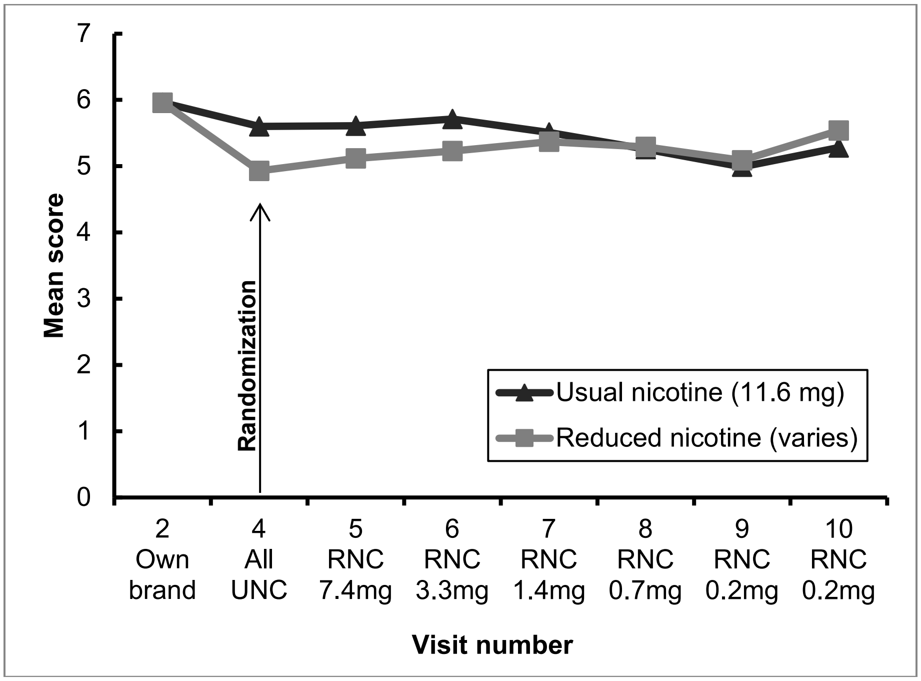

Figure S22. Randomized Phase Compliers (UNC, n=62; RNC, n=41)

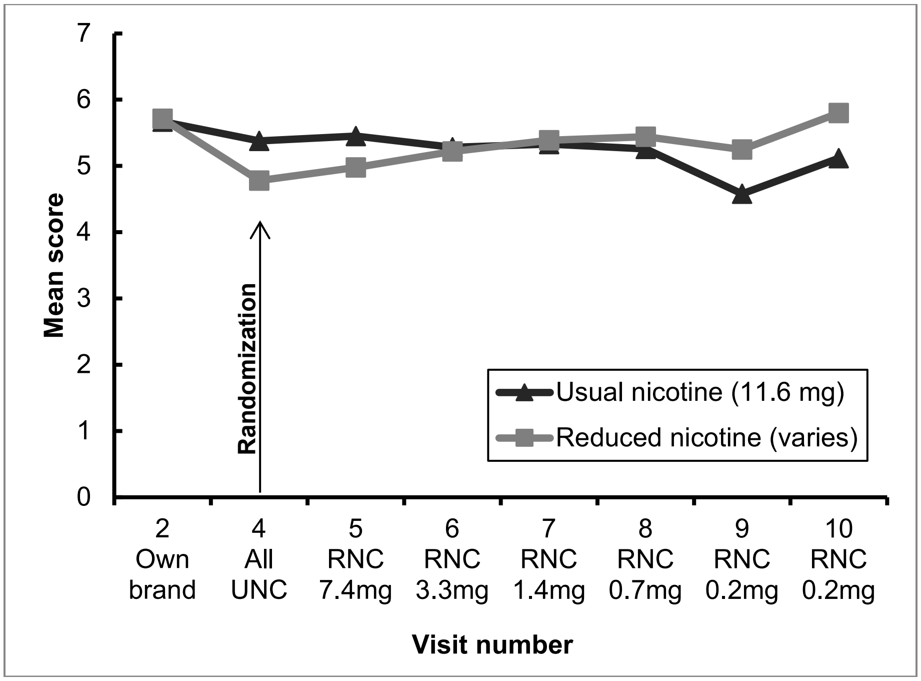

QIDS (Quick Inventory of Depressive Symptomatology – Range: 0-27)

Figure S23. All Randomized Participants (UNG n=94, RNG n=94)

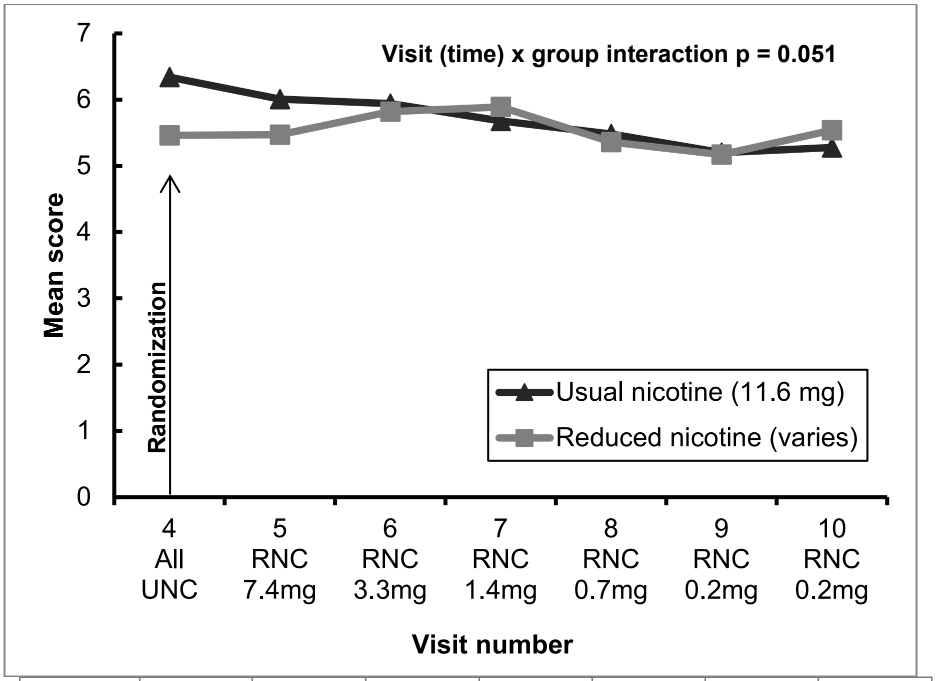

| | V4 | V5 | V6 | V7 | V8 | V9 | V10 |
|---|---|---|---|---|---|---|---|
| UNG | 93 | 86 | 82 | 82 | 80 | 75 | 72 |
| RNG | 94 | 89 | 87 | 80 | 75 | 71 | 69 |

OASIS (Overall Anxiety Severity and Impairment Scale – Range: 0-20)

Figure S24. Randomized Phase Completers (UNC, n=74; RNC, n=69)

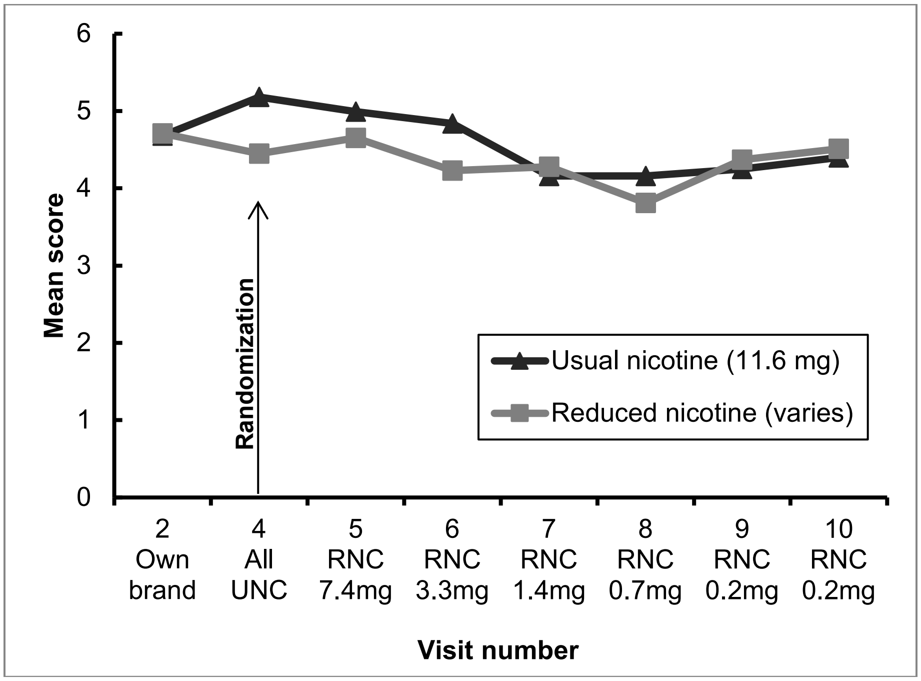

Figure S25. Randomized Phase Compliers (UNC, n=62; RNC, n=41)

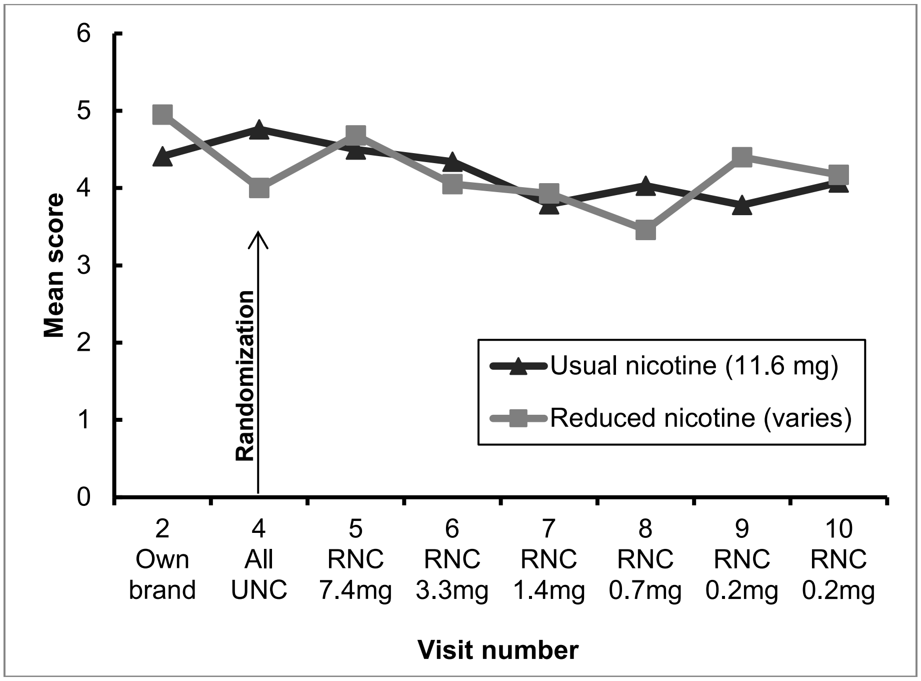

OASIS (Overall Anxiety Severity and Impairment Scale – Range: 0-20)

Figure S26. All Randomized Participants (UNG n=94, RNG n=94)

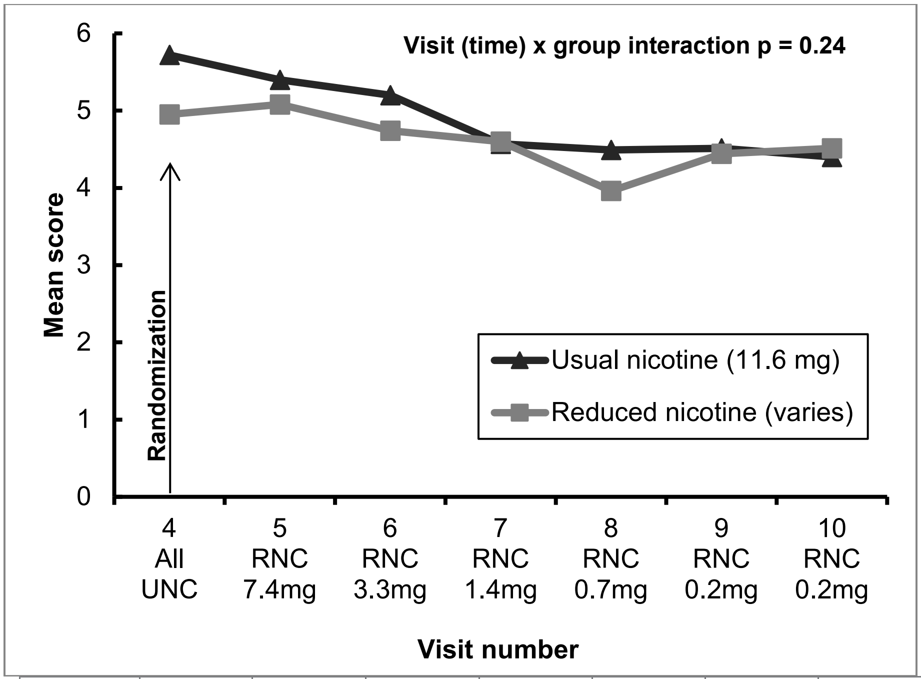

| | V4 | V5 | V6 | V7 | V8 | V9 | V10 |
|---|---|---|---|---|---|---|---|
| UNG | 94 | 88 | 84 | 82 | 81 | 76 | 73 |
| RNG | 94 | 90 | 87 | 81 | 75 | 71 | 68 |

Kessler K6 (Psychological Distress – Range: 0-24)

Figure S27. Randomized Phase Completers (UNC, n=74; RNC, n=69)

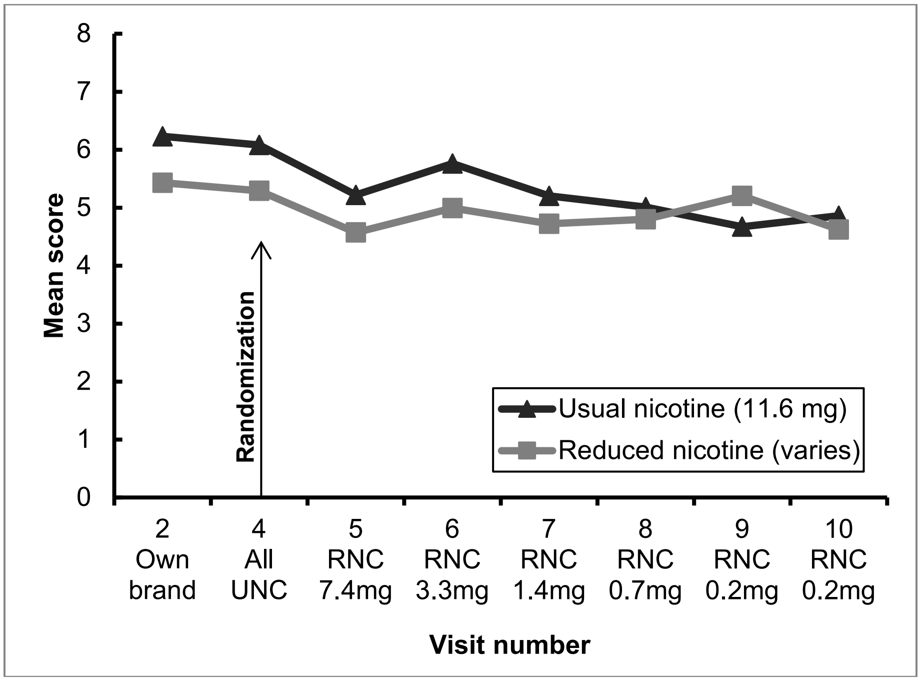

Figure S28. Randomized Phase Compliers (UNC, n=62; RNC, n=41)

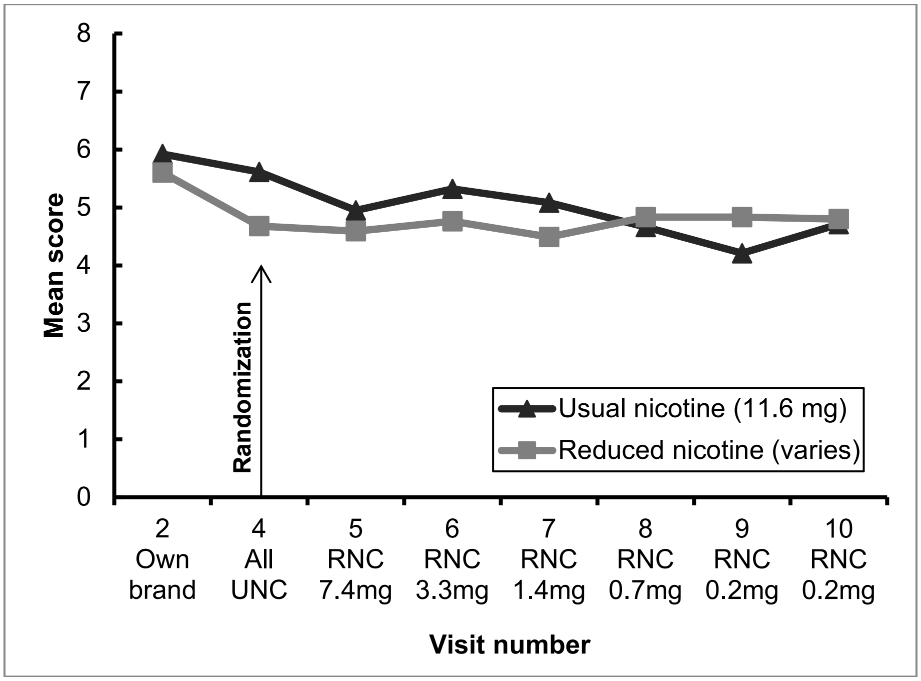

Kessler K6 (Psychological Distress – Range: 0-24)

Figure S29. All Randomized Participants (UNG n=94, RNG n=94)

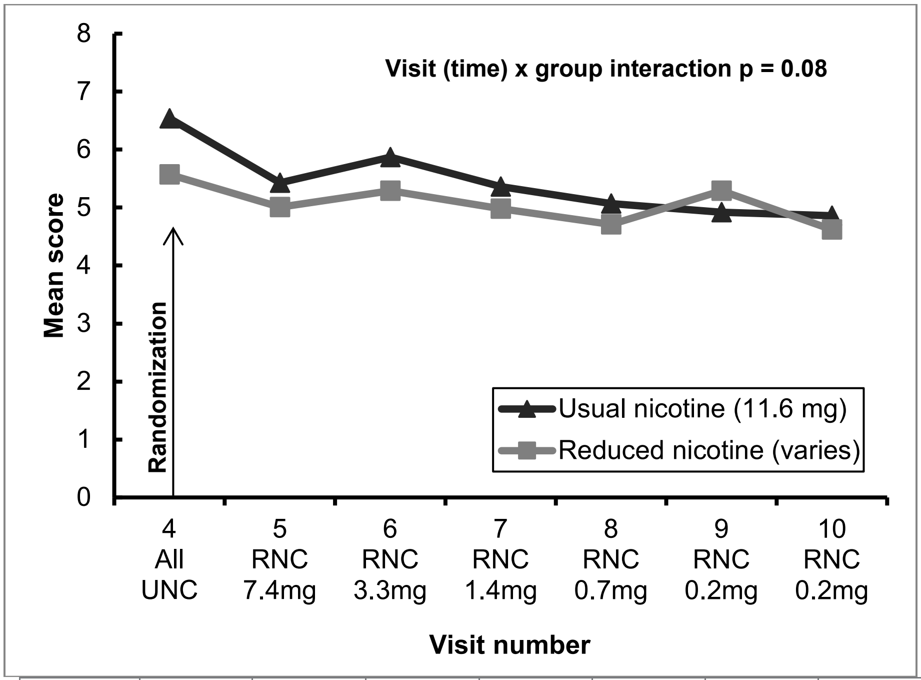

| | V4 | V5 | V6 | V7 | V8 | V9 | V10 |
|---|---|---|---|---|---|---|---|
| UNG | 93 | 88 | 84 | 83 | 81 | 77 | 74 |
| RNG | 94 | 90 | 87 | 81 | 75 | 72 | 69 |

PSS (Perceived Stress Scale – Range: 0-40)

Figure S30. Randomized Phase Completers (UNC, n=74; RNC, n=69)

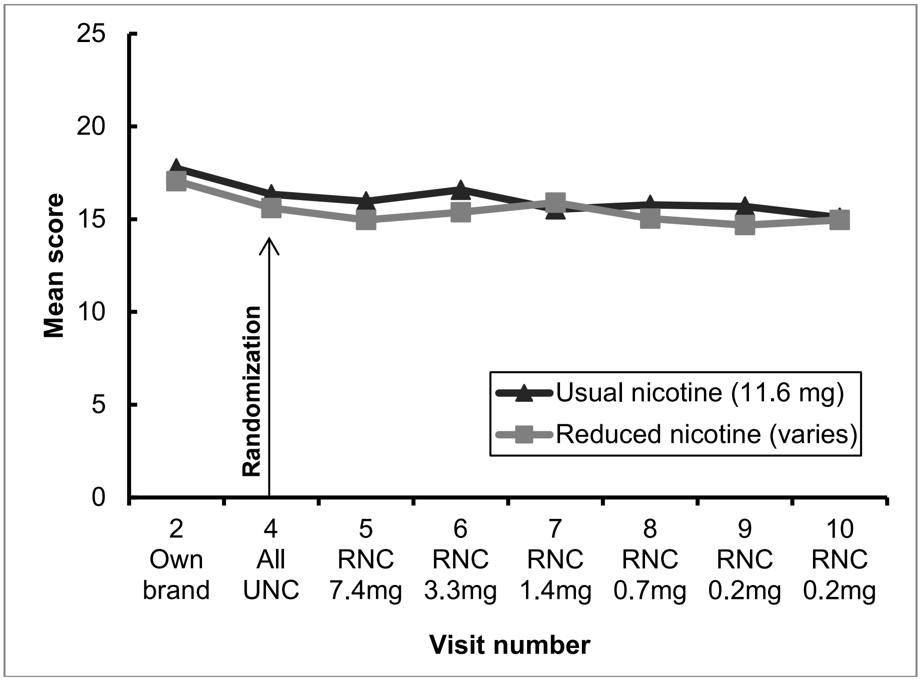

Figure S31. Randomized Phase Compliers (UNC, n=62; RNC, n=41)

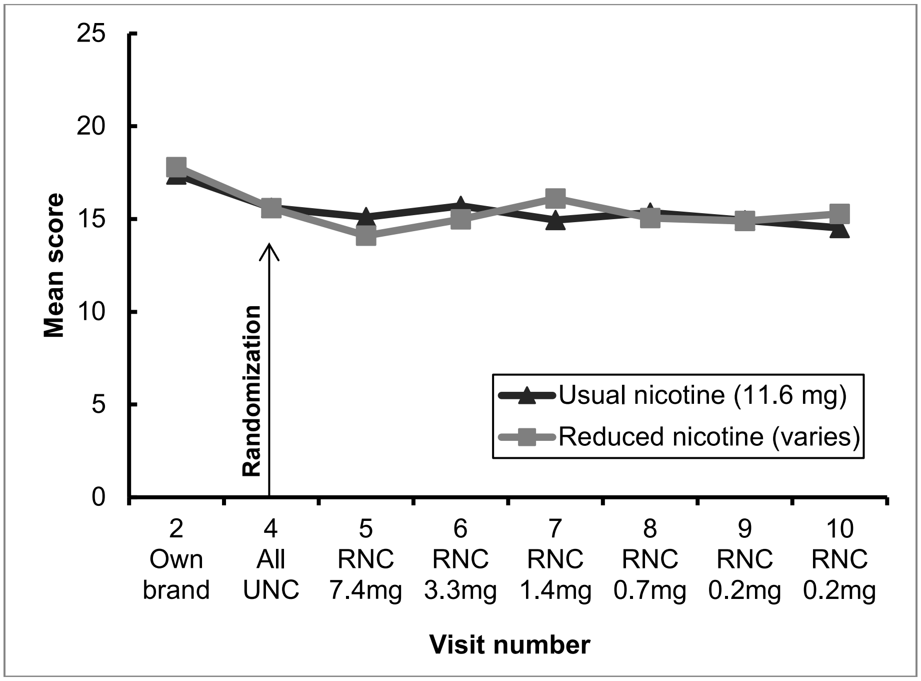

PSS (Perceived Stress Scale – Range: 0-40)

Figure S32. All Randomized Participants (UNG n=94, RNG n=94)

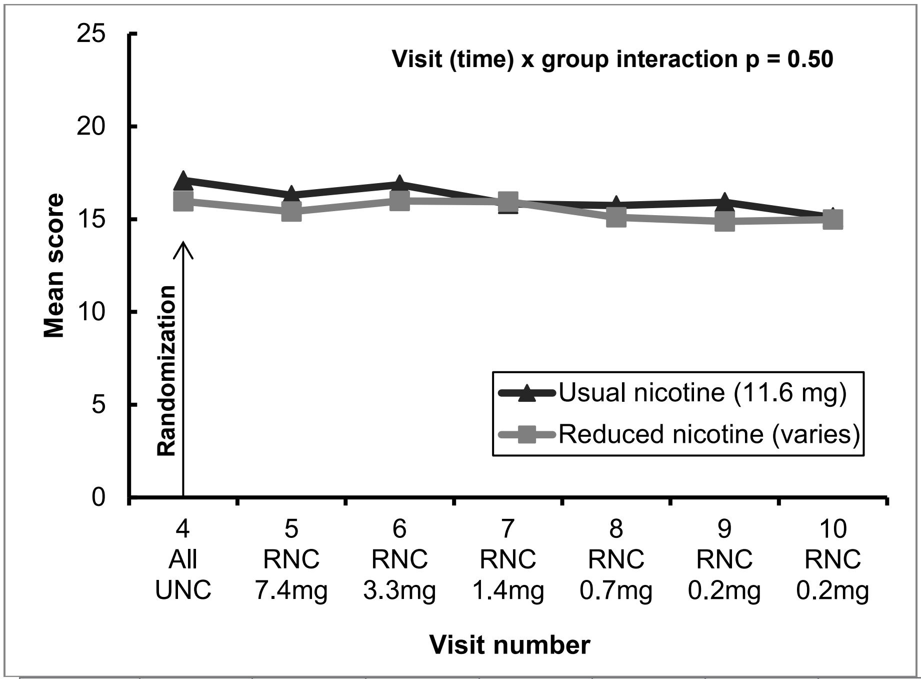

| | V4 | V5 | V6 | V7 | V8 | V9 | V10 |
|---|---|---|---|---|---|---|---|
| UNG | 94 | 87 | 84 | 83 | 81 | 77 | 74 |
| RNG | 94 | 90 | 87 | 81 | 75 | 72 | 69 |

CES-D (Center for Epidemiological Studies - Depression Scale – Range: 0-60)

Figure S33. Randomized Phase Completers (UNC, n=74; RNC, n=69)

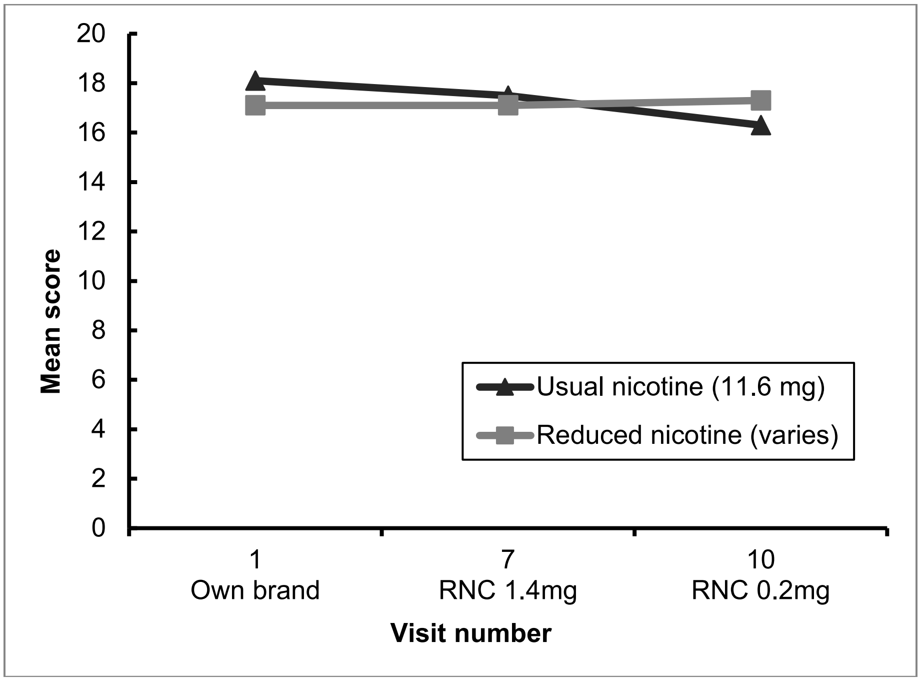

Figure S34. Randomized Phase Compliers (UNC, n=62; RNC, n=41)

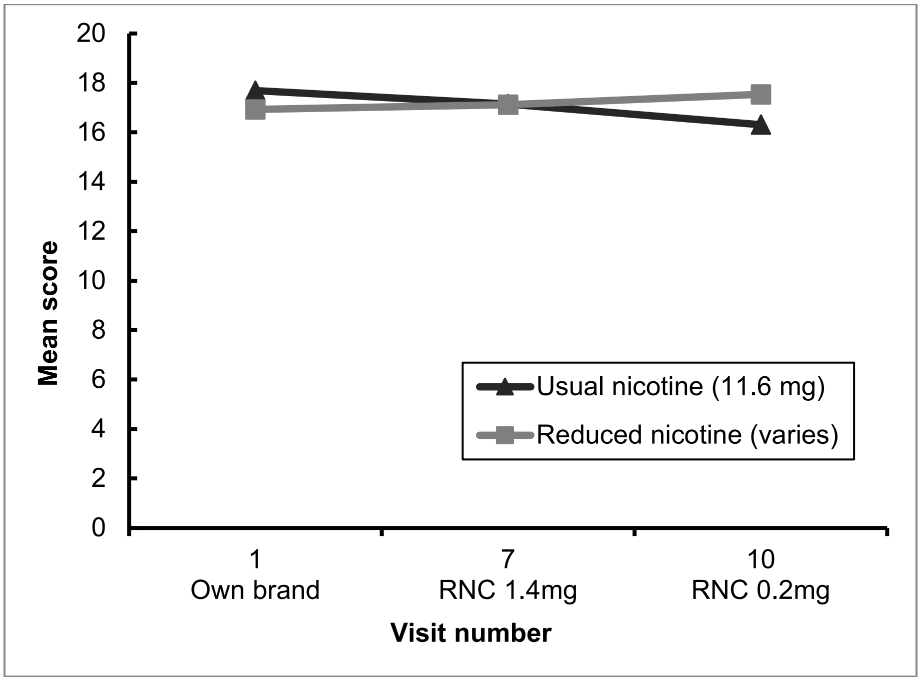

CCQ Total (Clinical COPD Questionnaire – Range: 0-6)

Figure S35. Randomized Phase Completers (UNC, n=74; RNC, n=69)

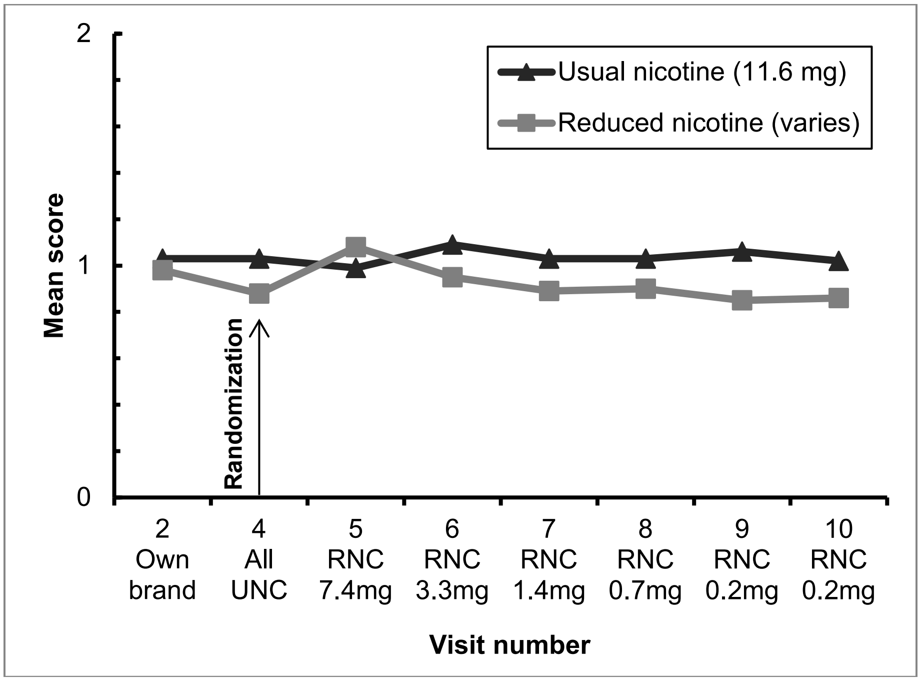

Figure S36. Randomized Phase Compliers (UNC, n=62; RNC, n=41)

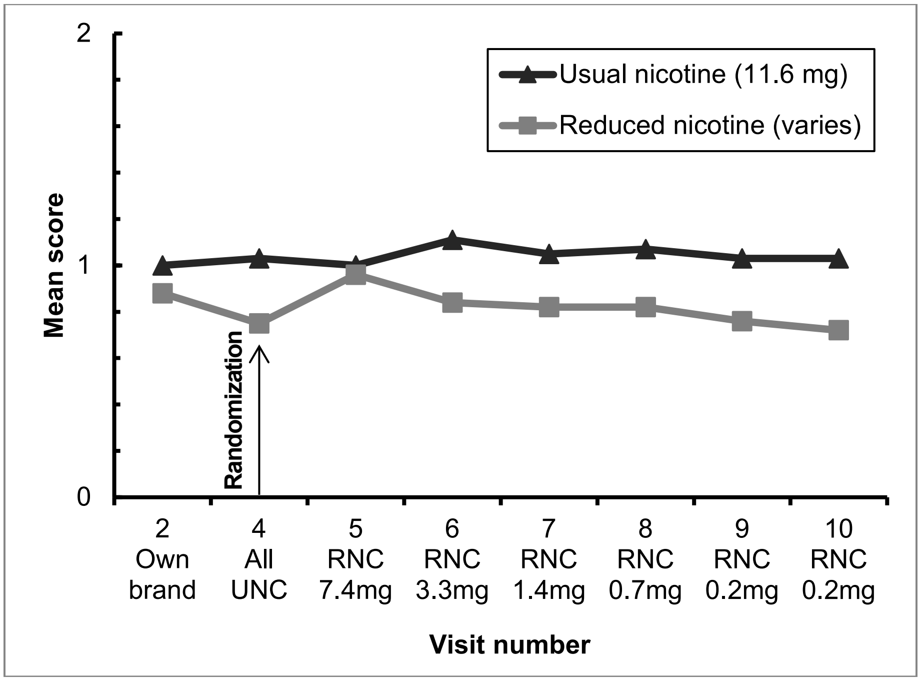

Systolic Blood Pressure (mmHg)

Figure S37. Randomized Phase Completers (UNC, n=74; RNC, n=69)

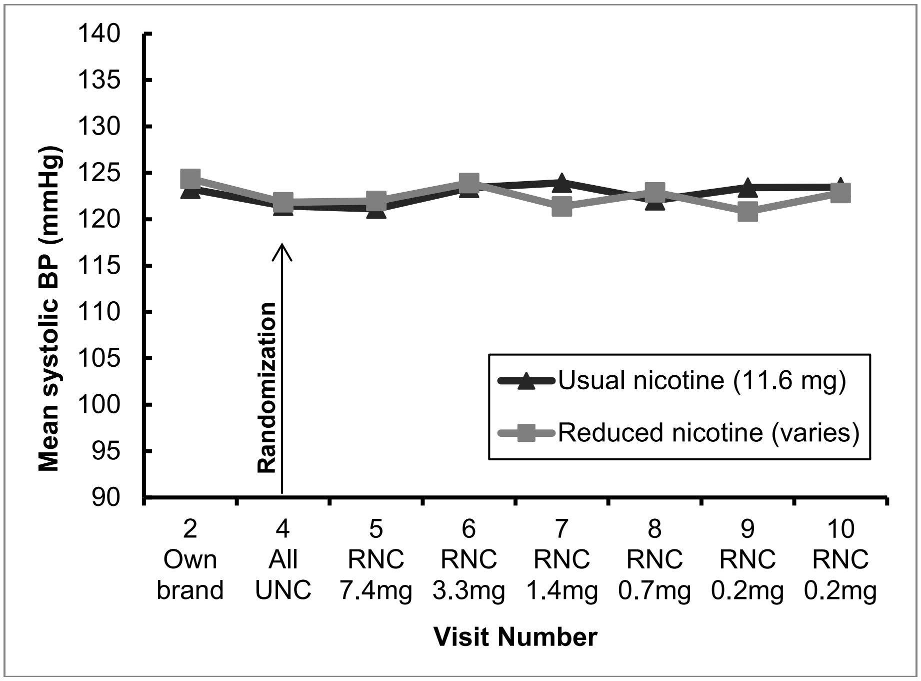

Figure S38. Randomized Phase Compliers (UNC, n=62; RNC, n=41)

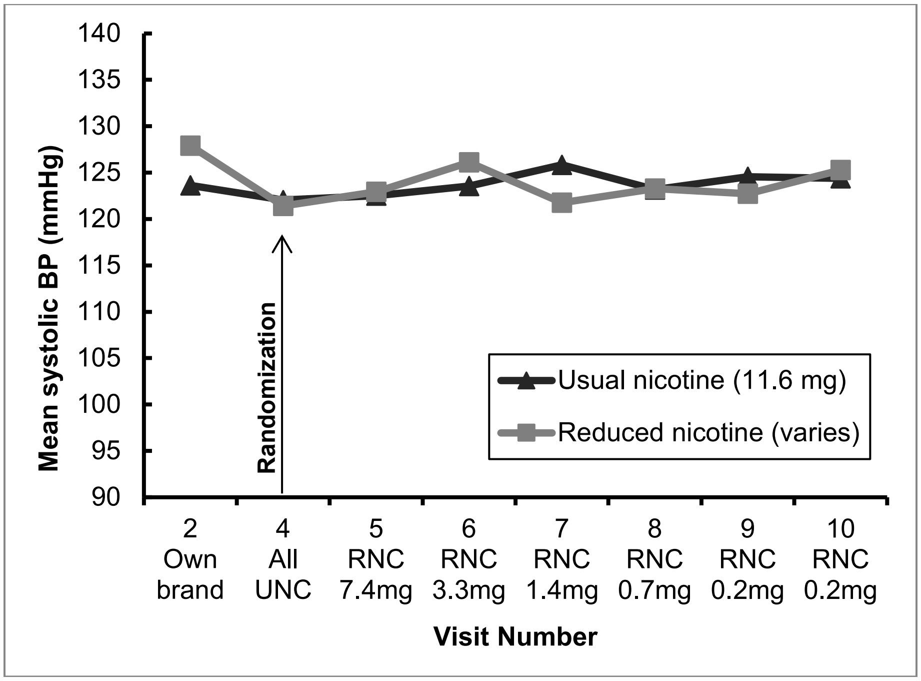

Diastolic Blood Pressure (mmHg)

Figure S39. Randomized Phase Completers (UNC, n=74; RNC, n=69)

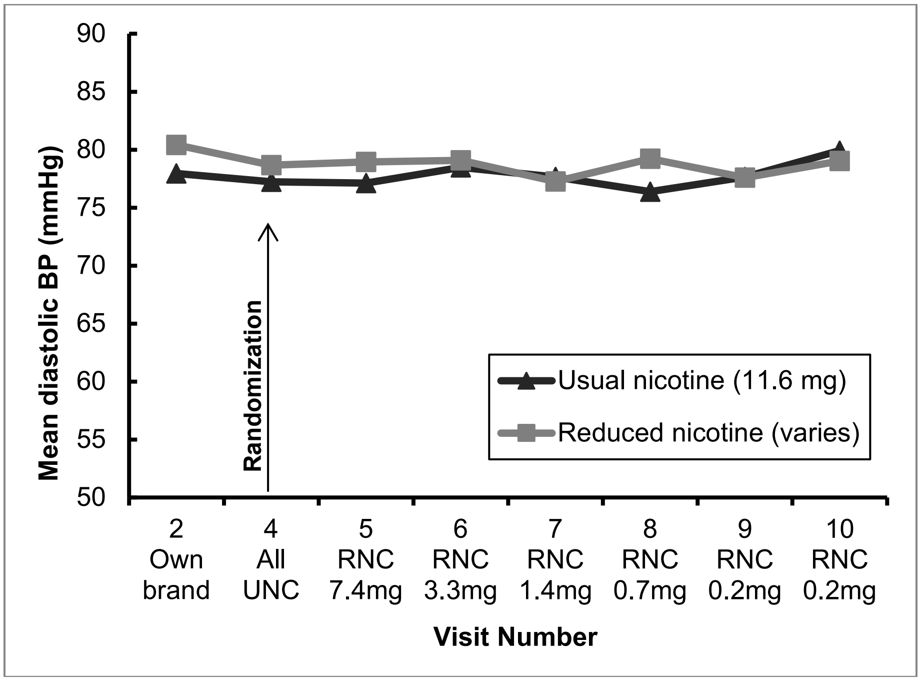

Figure S40. Randomized Phase Compliers (UNC, n=62; RNC, n=41)

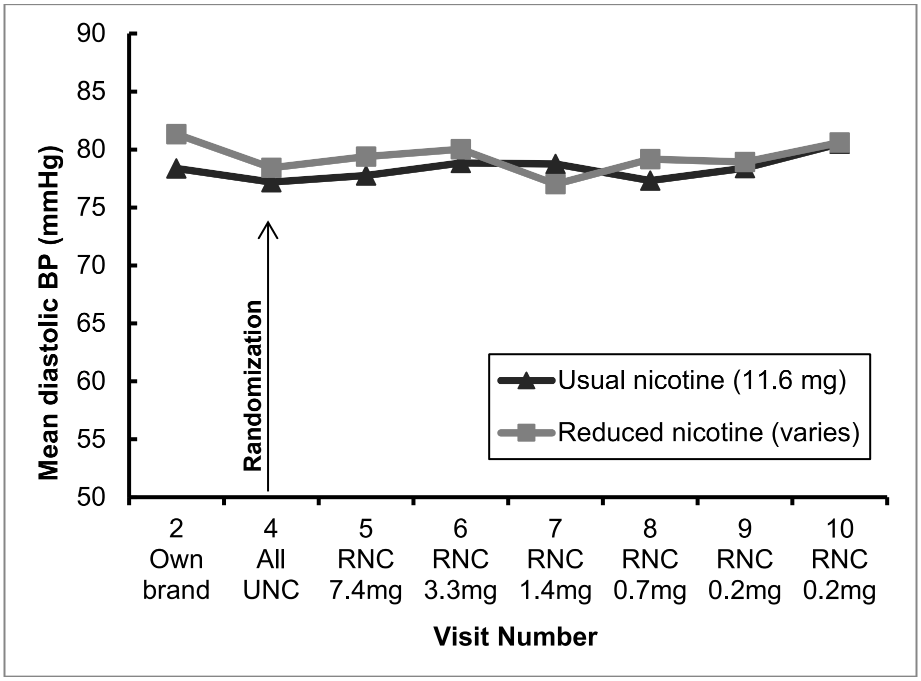

Heart Rate (beats per minute)

Figure S41. Randomized Phase Completers (UNC, n=74; RNC, n=69)

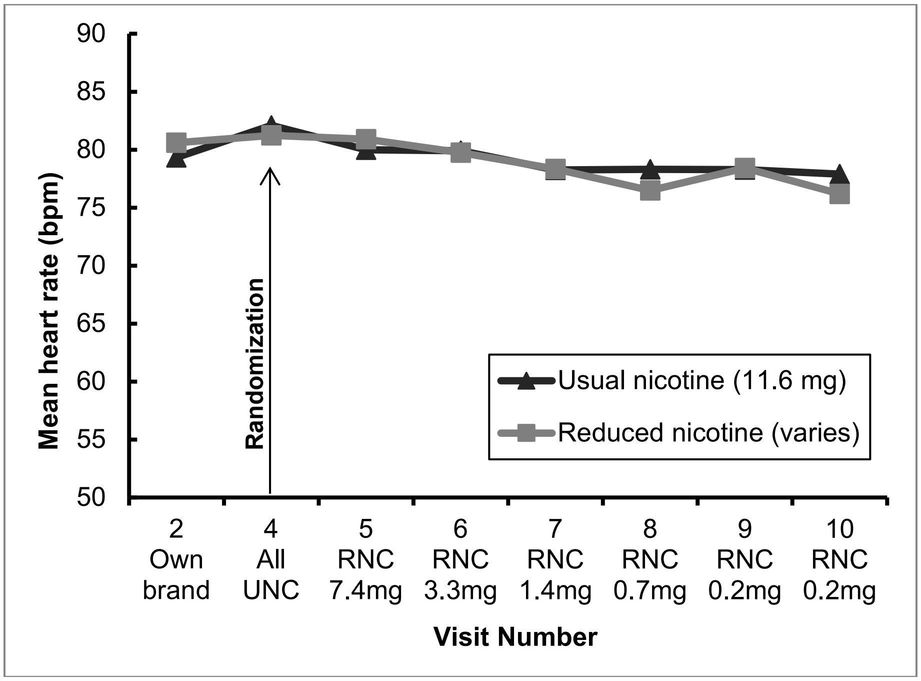

Figure S42. Randomized Phase Compliers (UNC, n=62; RNC, n=41)

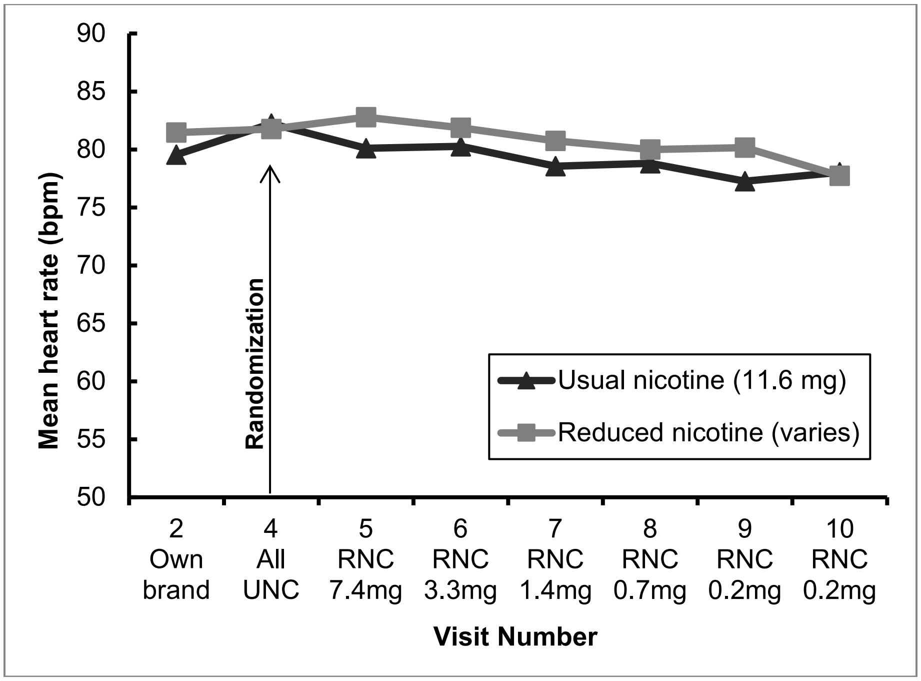

Body Weight (pounds)

Figure S43. Randomized Trial Phase Completers (UNC, n=74; RNC, n=69)

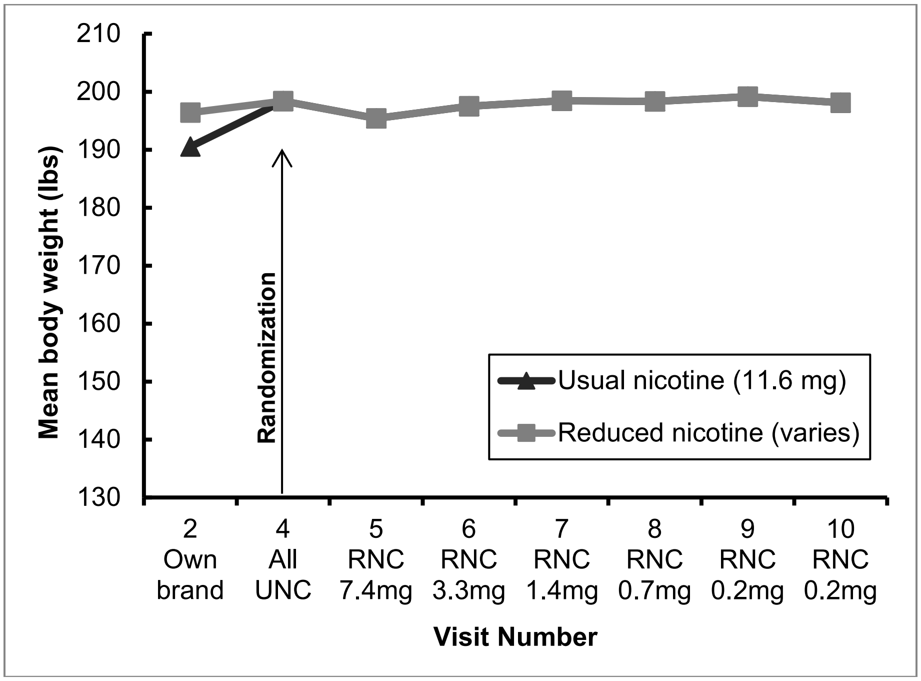

Figure S44. Randomized Phase Compliers (UNC, n=62; RNC, n=41)

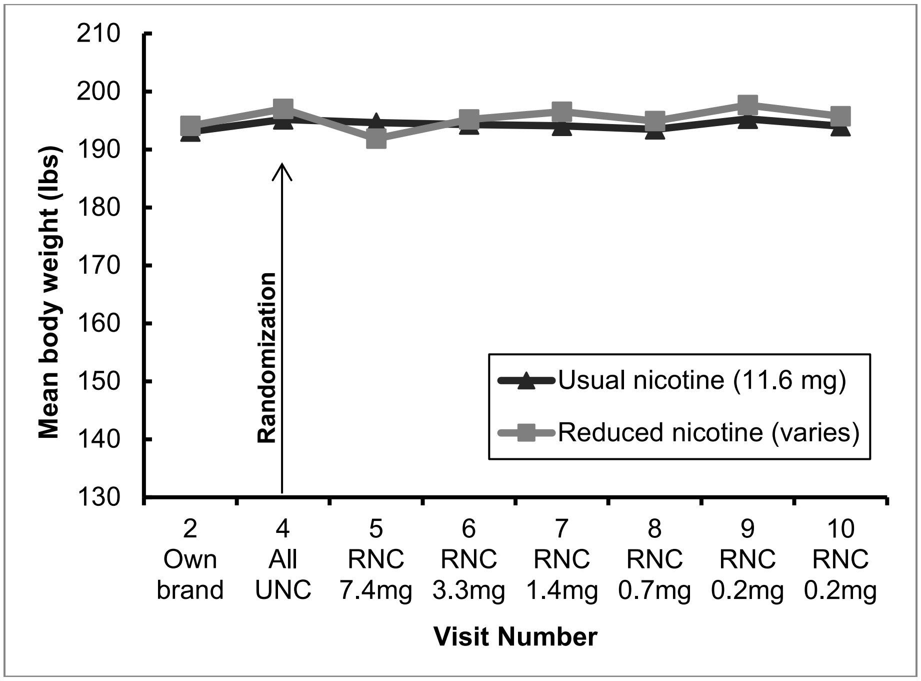

FEV1 (Forced Expiratory Volume (liters) in 1 second)

Figure S45. Randomized Phase Completers (UNC, n=74; RNC, n=69)

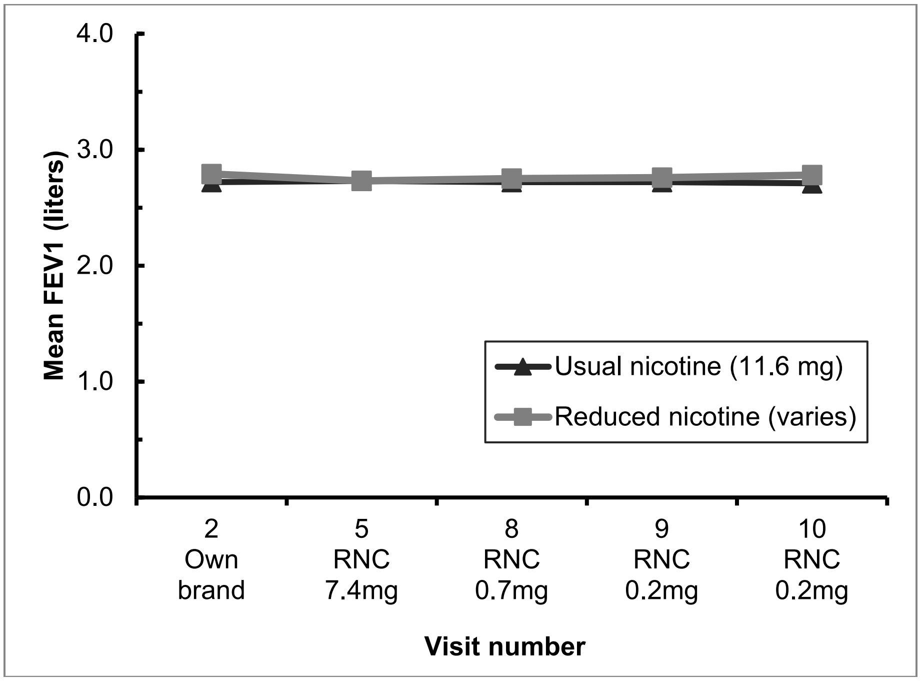

Figure S46. Randomized Phase Compliers (UNC, n=62; RNC, n=41)

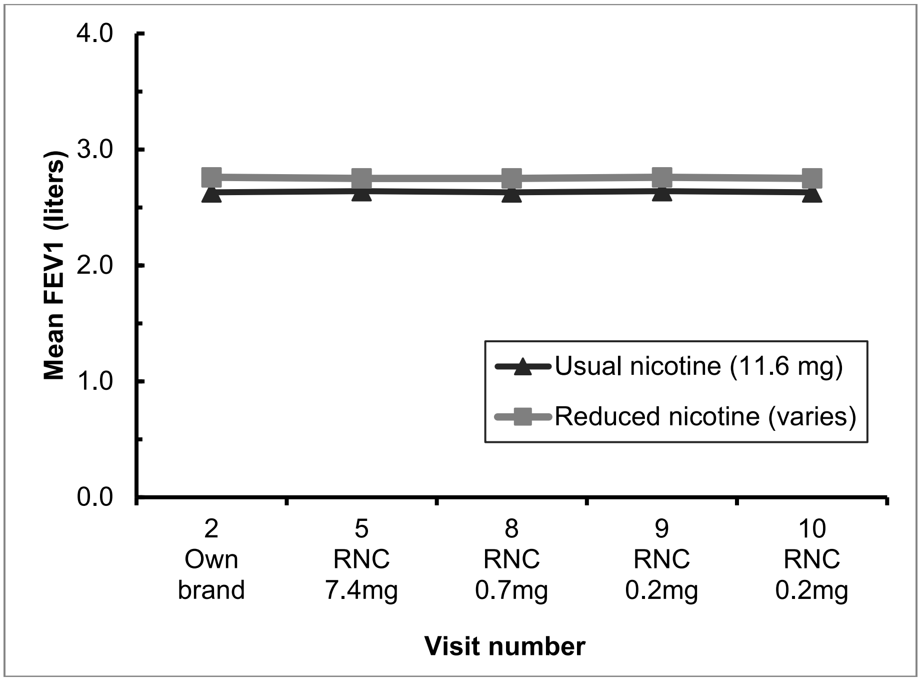

Blood Glutathione GSSP:GSH Ratio (Ratio of Glutathione to Oxidized Glutathione)
Biomarker was analyzed in a randomly selected subgroup of 50 participants (evenly stratified by study group at the PSU site) who provided both urine and blood samples at V4, V6, and V10.

Figure S47. Randomized Phase Completers (UNC, n=25; RNC, n=25)

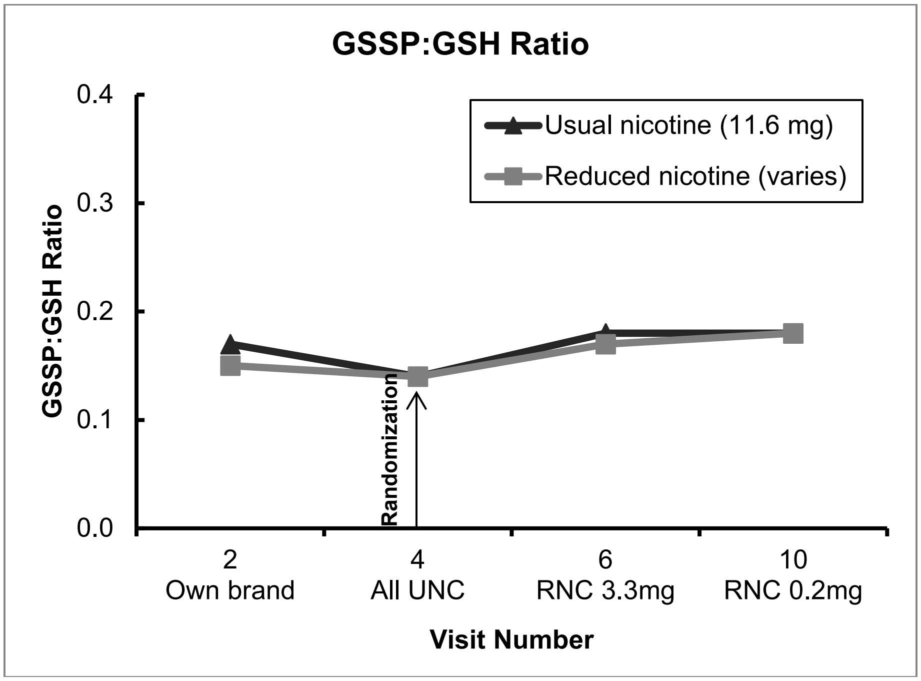

Figure S48. Randomized Phase Compliers (UNC, n=20; RNC, n=18)

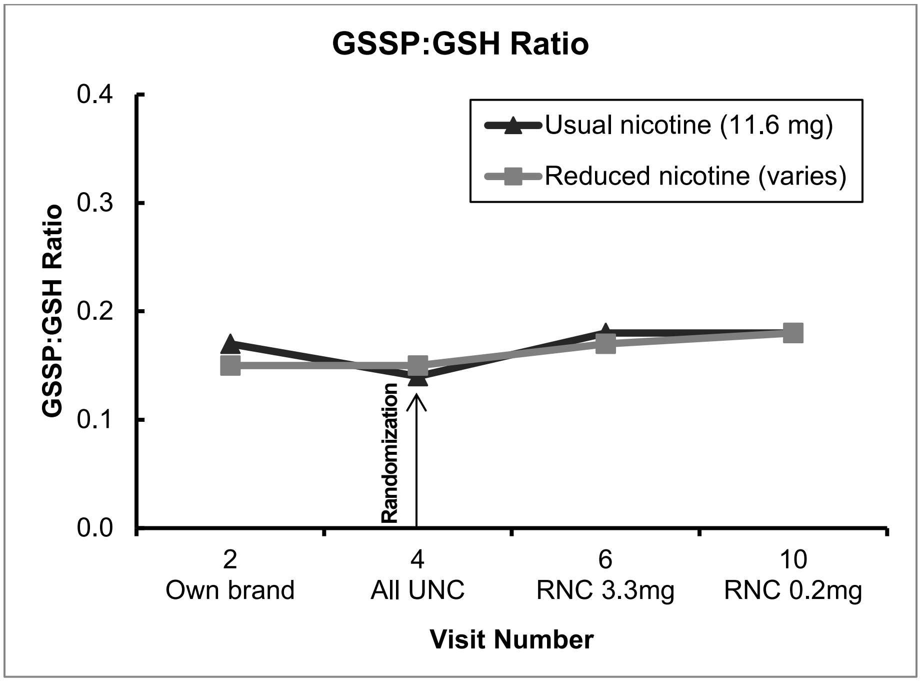

Urinary 8-Isoprostane, corrected for urinary creatinine
Biomarker was analyzed in a randomly selected subgroup of 50 participants (evenly stratified by study group at the PSU site) who provided both urine and blood samples at V4, V6, and V10.

Figure S49. Randomized Phase Completers (UNC, n=25; RNC, n=25)

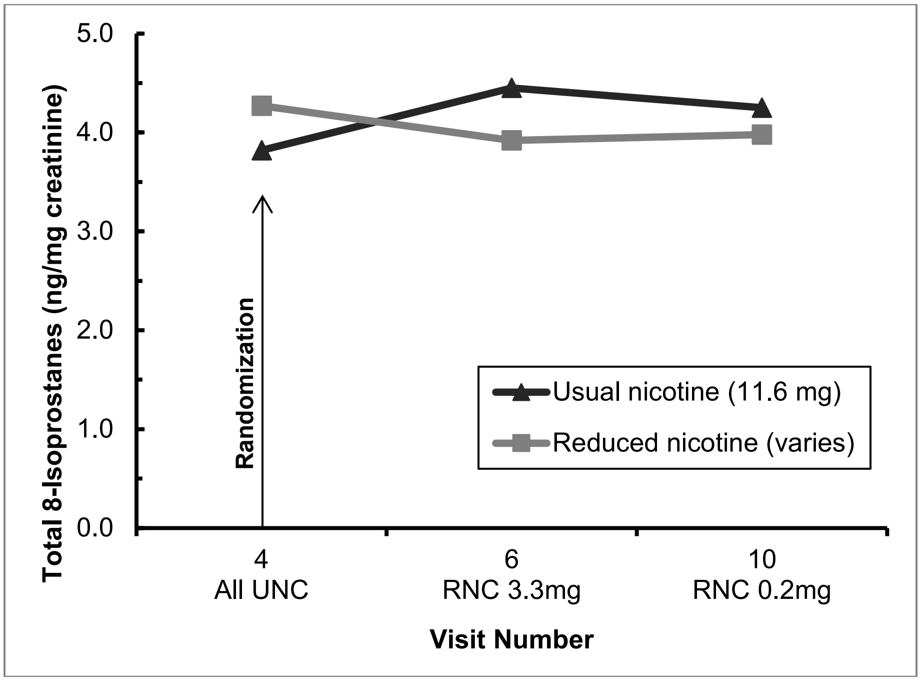

Figure S50. Randomized Phase Compliers (UNC, n=20; RNC, n=18)

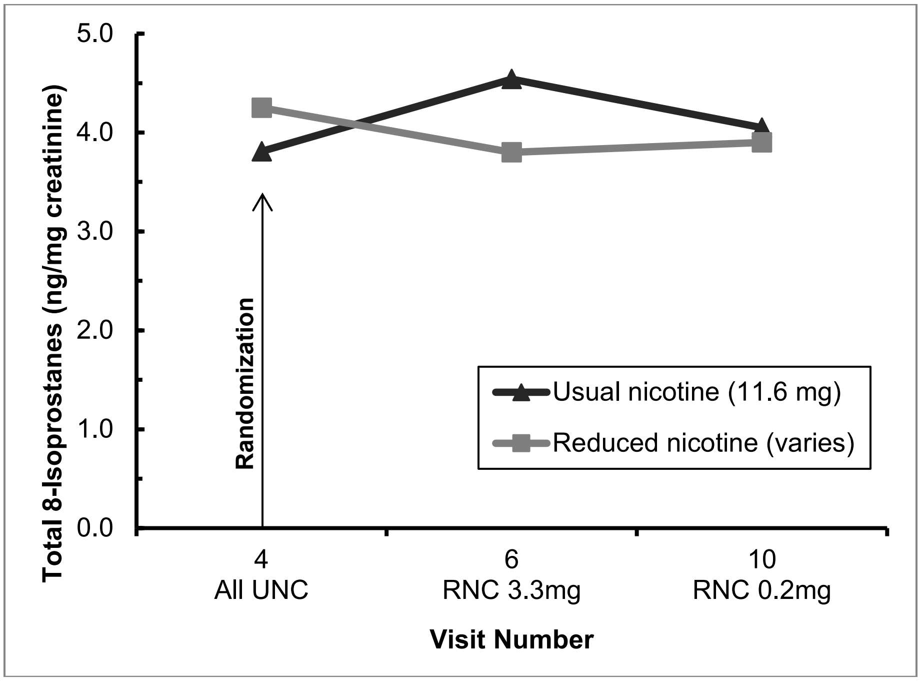

Total Urinary NNAL: 4-(methylnitrosamino)-1-(3-pyridyl)-1-butanol, corrected for urinary creatinine. Biomarker was analyzed in a randomly selected subgroup of 52 participants (evenly stratified by site and by study group) who provided urine samples at the randomization visit (V4) and the end of randomized phase visit (V10).

Figure S51. Randomized Phase Completers (UNC, n=26; RNC, n=26)

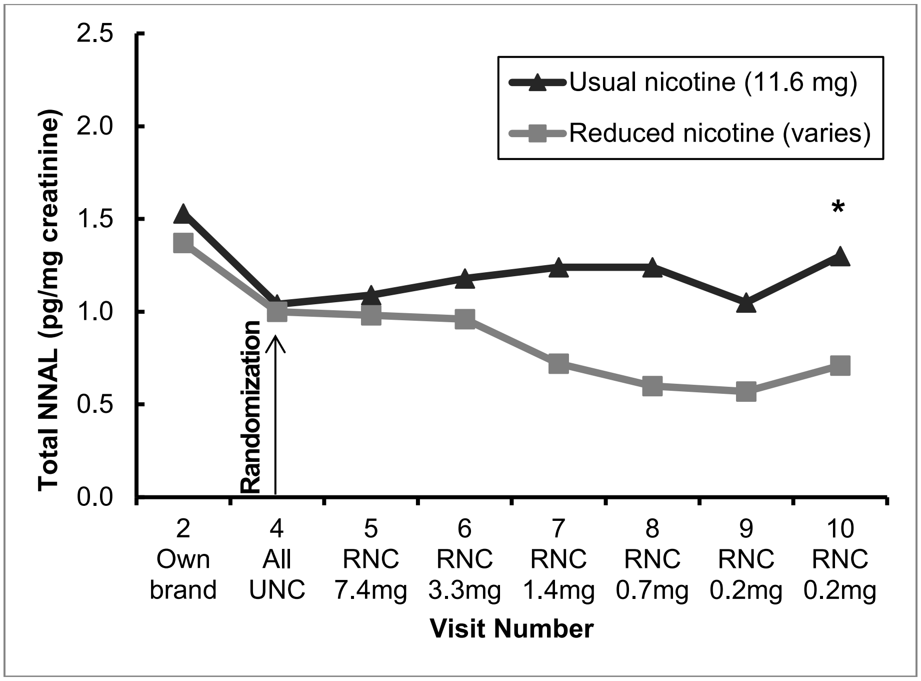

Figure S52. Randomized Phase Compliers (UNC, n=24; RNC, n=13)

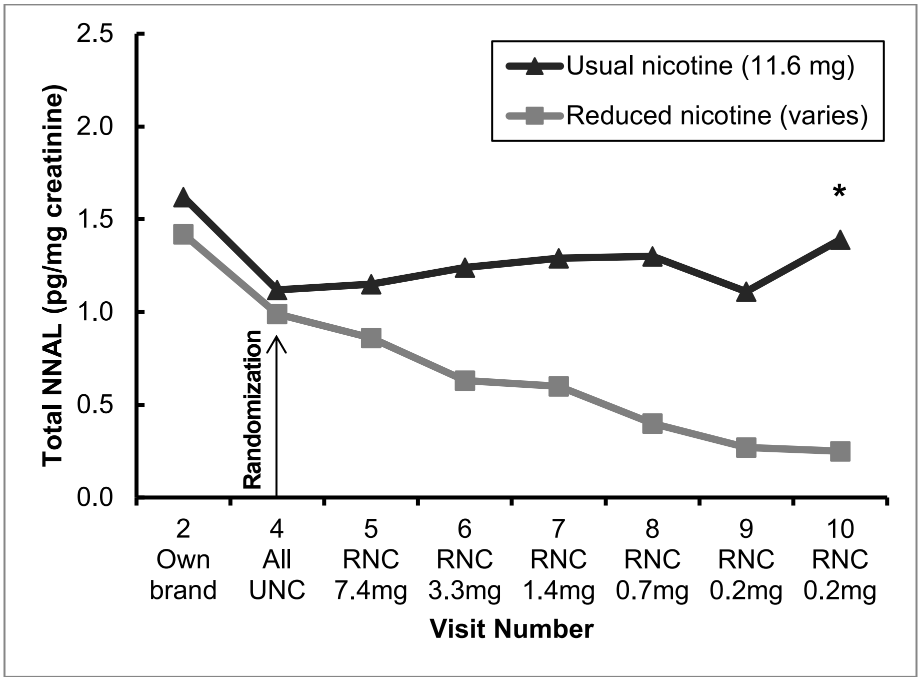

Total Urinary 1-Hydroxypyrene, corrected for urinary creatinine. Biomarker was analyzed in a randomly selected subgroup of 52 participants (evenly stratified by site and by study group) who provided urine samples at the randomization visit (V4) and the end of randomized phase visit (V10). Figure S53. Randomized Phase Completers (UNC, n=26; RNC, n=26)

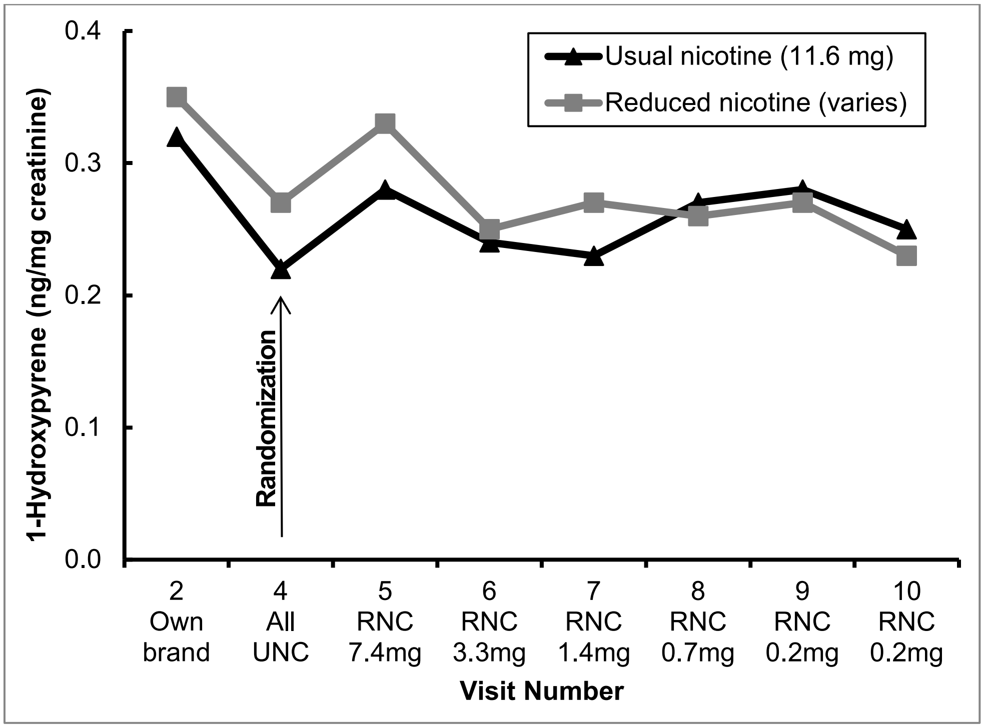

Figure S54. Randomized Phase Compliers (UNC, n=24; RNC, n=13)

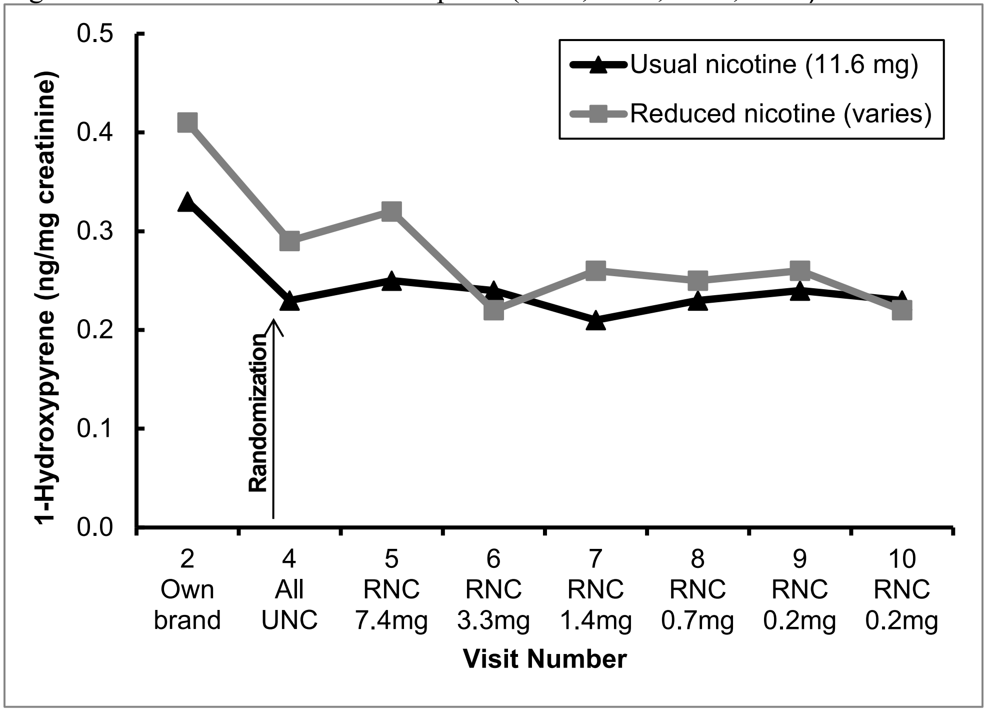

Table S1. Study Measures Time and Events Schedule

| | Baseline | | | | Randomization | | | | | | Treatment Choice | |
|---|---|---|---|---|---|---|---|---|---|---|---|---|
| | Baseline I | | Baseline II | | Step 1 | Step 2 | Step 3 | Step 4 | Step 5 | | | |
| **Study Week Number** | **0** | **1** | **2** | **3** | **6** | **9** | **12** | **15** | **18** | **21** | **25** | **33** |
| **Study Day** | 0 | 7 | 14 | 21 | 42 | 63 | 84 | 105 | 126 | 147 | 175 | 231 |
| **Study Visit Number** | 1 | 2 | 3 | 4 | 5 | 6 | 7 | 8 | 9 | 10 | 11 | 12 |
| **Measures/Questionnaires** | | | | | | | | | | | | |
| Tobacco and marijuana use and daily cigarette log | | X | X | X | X | X | X | X | X | X | X | X |
| Concomitant medications | | X | X | X | X | X | X | X | X | X | X | X |
| Adverse events(5) | | X | X | X | X | X | X | X | X | X | X | X |
| Demographics | X | | | | | | | | | | | |
| Tobacco use history, cigarette details | X | | | | | | X | | | X | | |
| NIDA drug use (past 3 mo) (6) | X | | | | | | X | | | X | | |
| Nicotine dependence ( FTND, PSCDI) (7, 8) | X | X | X | X | X | X | X | X | X | X | X | X |
| Minnesota Nicotine Withdrawal Scale (9) | X | X | X | X | X | X | X | X | X | X | X | X |
| Questionnaire on Smoking urges (10, 11) | X | X | | X | X | X | X | X | X | X | X | X |
| Anxiety symptoms questionnaire (ASQ) (12, 13) | X | | | | | | X | | | X | X | X |
| CES-D (14) | X | | | | | | X | | | X | X | X |
| Kessler K6 scale (15, 16) | X | X | | X | X | X | X | X | X | X | X | X |
| OASIS (anxiety) (17) | X | X | | X | X | X | X | X | X | X | X | X |
| QIDS (depression) (18) | X | X | | X | X | X | X | X | X | X | X | X |
| Perceived Stress Scale (19) | X | X | | X | X | X | X | X | X | X | X | X |
| Clinical COPD questionnaire (20) | | X | | X | X | X | X | X | X | X | X | X |
| NRT disbursement (if chosen by participant) | | | | | | | | | | X | X | |
| **Biomeasures/Biomarkers** | | | | | | | | | | | | |
| Weight (21) | X | X | X | X | X | X | X | X | X | X | X | X |
| Height (21) | X | | | | | | | | | | | |

| | | | | | | | | | | | | |
|---|---|---|---|---|---|---|---|---|---|---|---|---|
| Exhaled CO (22) | X | X | X | X | X | X | X | X | X | X | X | X |
| Blood pressure/Heart Rate (23) | X | X | X | X | X | X | X | X | X | X | X | X |
| Pulmonary function (spirometry) (24) | X | X | | | X | | | X | X | X | X | X |
| Urinary 8-Isoprostane[#] | | | | X | | X | | | | X | | |
| Total Urinary NNAL, 1-HOP[†] | | X | X | X | X | X | X | X | X | X | | |
| Pregnancy test | X | | | X | | X | | X | | X | X | X |
| Plasma Cotinine | | X | X | X | X | X | X | X | X | X | | |
| Blood Glutathione (GSSP:GSH ratio)[#] | | X | | X | | X | | | | X | | |
| Participant Payment | $40 | $80 | $40 | $80 | $80 | $80 | $80 | $80 | $80 | $80 | $80 | $80+ compliance |

[†]Biomarker was analyzed in a randomly selected subgroup of 52 participants (evenly stratified by site and by study group) who provided urine samples at the randomization visit (V4) and the end of randomized trial phase visit (V10). #Biomarker was analyzed in a randomly selected subgroup of 50 participants (evenly stratified by study group at the PSU site) who provided both urine and blood samples at V4, V6, and V10.

NIDA=National Institute on Drug Abuse, HONC = Hooked on Nicotine Checklist, FTND = Fagerström Test for Nicotine Dependence, PSCDI = Penn State Cigarette Dependence Index, CES-D = Center for Epidemiologic Studies Depression Scale, OASIS = Overall Anxiety Severity and Impairment Scale, QIDS = Quick Inventory of Depressive Symptomatology, COPD = chronic obstructive pulmonary disease, NRT = nicotine replacement therapy, NNAL = (4-(methylnitrosamino)-1-(3-pyridyl)-1-butanol), 1-HOP = 1-hydroxypyrene

Table S2. Nicotine Content Dosing Schedule

| Phase | Baseline I | Baseline II | Randomized Double-Blind Nicotine Reduction Phase | | | | | Treatment Choice Phase |
|---|---|---|---|---|---|---|---|---|
| **Week(s)** | **1** | **2** | **3** | **3** | **3** | **3** | **6** | **12** |
| **Cigarette type** | Own Brand | Usual Nicotine Research Cigarettes | Reduced Nicotine Step 1 | Reduced Nicotine Step 2 | Reduced Nicotine Step 3 | Reduced Nicotine Step 4 | Reduced Nicotine Step 5 | Variable |
| **Regular SPECTRUM Code** | | 600 | 500 | 400 | 300 | 200 | 102 | |
| **Menthol SPECTRUM Code** | | 601 | 501 | 401 | 301 | 201 | 103 | |
| | | *Approximate nicotine content in mgs per cigarette (mg/gram)** | | | | | | |
| **RNC** | Around 13 mg[#] (19) | 11.6 (16.5) | 7.4 (10.6) | 3.3 (4.7) | 1.4 (1.9) | 0.7 (0.9) | 0.2 (0.3) | Variable |
| **UNC** | Around 13 mg[#] (19) | 11.6 (16.5) | 11.6 (16.5) | 11.6 (16.5) | 11.6 (16.5) | 11.6 (16.5) | 11.6 (16.5) | Variable |

*These are averages of menthol/non-menthol cigarettes at each level based on estimated 0.7g tobacco content per cigarette and nicotine concentrations based on Richter et al 2016 (25).
[#]Estimated mean nicotine content and concentration based on Connolly et al (2007) (26).

A recent pharmacokinetic study (27) of the regular (non-mentholated) types of SPECTRUM research cigarettes (Codes 102, 400 and 600) reported that the boost in blood nicotine concentration from smoking a single cigarette in the lab was dose-dependent, with a boost of 0.3, 3.9 and 17.3 ng/ml for low (102s), medium (400s), and high (600s) nicotine content SPECTRUM cigarettes. The high dose SPECTRUM had a similar nicotine boost to the "preferred brand" cigarettes (19 ng/ml).

Table S3: Demographic and Smoking Characteristics for Randomized Phase Completers

| | **RNC (n = 69)** | **UNC (n = 74)** |
|---|---|---|
| Study Enrollment (V1) | | |
| Female | 71.0 (49) | 62.2 (46) |
| Race | | |
| African American | 15.9 (11) | 13.5 (10) |
| White | 71.0 (49) | 81.1 (60) |
| Other | 13.0 (9) | 5.4 (4) |
| Age (in years) | 43.4 (11, 23-65) | 45.3 (12.6, 21-65) |
| Bachelor's degree or higher | 14.5 (10) | 24.3 (18) |
| Currently employed full-time | 40.6 (28) | 40.5 (30) |
| Number of Prior Lifetime Quit Attempts; Median (Range) | 2 (0-12) | 3 (0-100) |
| No Prior Lifetime Quit Attempts | 20.3 (14) | 25.7 (19) |
| Menthol Flavor Preference | 39.1 (27) | 37.8 (28) |
| Number of years as daily smoker | 25.3 (11.3, 2-49) | 28.1 (12.9, 4-53) |
| CES-D score | 17.1 (8.5, 4-43) | 18.1 (7.1, 6-36) |
| Lifetime suicidality | 34.8 (24) | 27.0 (20) |
| Number of MINI Mood/Anxiety Disorder Diagnoses[C] | | |
| One current diagnosis | 31.9 (22) | 35.1 (26) |
| Two or more current diagnoses | 24.6 (17) | 31.1 (23) |
| Past diagnosis/-es only | 43.5 (30) | 33.8 (25) |
| Current Mood Disorder[C], % | 18.8 (13) | 21.6 (16) |
| Past Mood Disorder[C] | 69.6 (48) | 63.5 (47) |
| CurrentAnxiety Disorder[C], % | 47.8 (33) | 62.2 (46) |
| Past Anxiety Disorder[C] | 44.9 (31) | 50.0 (37) |
| Reported Use of at least one medication for psychiatric reasons | 56.5 (39) | 56.8 (42) |
| Baseline Phase I (V2) | | |
| Cigarettes per day[D] | 17.7 (10.5, 5-60) | 19.6 (10.2, 1.3-58) |
| Exhaled carbon monoxide (in ppm)[B] | 28.8 (18.1, 4-100) | 28.8 (16.6, 6-85) |
| Moderate or High Environmental Tobacco Smoke Exposure Score | 69.6 (48) | 71.6 (53) |
| Fagerström Test for Nicotine Dependence score | 5.8 (2.4, 0-10) | 5.9 (2.3, 1-10) |
| Kessler K6 score | 5.4 (5.1, 0-22) | 6.2 (4.7, 0-18) |
| Penn State Cigarette Dependence Index score | 12.6 (3.5, 5-20) | 13.4 (3.4, 6-20) |

Continuous measures reported as Mean (SD, Range) – Unless otherwise specified
Categorical measures reported as Column % (Frequency)
[B]Reflects the average of two CO measurements if >1 CO was collected at the visit for safety monitoring
[C]Current MINI Mood Diagnoses considered: Major Depressive Disorder (Past 2 Weeks), Minor Depressive Disorder (Past 2 Weeks), Dysthymia (Past 2 Years), Premenstrual Dysphoric Disorder (Past Year), Mixed Anxiety Depressive Disorder (Past Month); Current MINI Anxiety Disorders considered: Panic Disorder (Past Month),

Agoraphobia (Past Month), Social Phobia (Past Month), Specific Phobia (Past Month), Obsessive Compulsive Disorder, Post-Traumatic Stress Disorder (Past Month), Generalized Anxiety Disorder (Past 6 Months); Past or Lifetime MINI Mood Diagnoses considered: Major Depressive Disorder, Minor Depressive Disorder, Dysthymia; Past or Lifetime MINI Anxiety Diagnoses considered: Panic Disorder, Agoraphobia, Post-Traumatic Stress Disorder
[D]Daily Cigarette Consumption was measured as the total (non-study + study) number of cigarettes reportedly smoked each day via daily log. The mean number of cigarettes smoked was calculated as the 6-day average immediately preceding each study visit.

**Adverse Events During Randomized Phase**

This section summarizes the adverse events (AE) that occurred during the randomized phase of the trial (start date of the reported AE occurred after V4 and prior to V10) by study group for randomized participants only (RNC n=94, UNC n=94). The proportion of participants who have experienced at least one AE whose onset (start date) occurred during the randomized phase of the trial is 75.5% (n=71) for the RNC and 77.7% (n=73) for the UNC. There were a total of 327 reported AEs (175 for the RNC and 152 for the UNC) with an onset during the randomized phase of the trial for randomized participants. Further details of the reported AEs are in the tables below.

Table S4. All Adverse Events by Expectedness and Relatedness to the Study Cigarette

| | **RNC** | **UNC** | **Overall** |
|---|---|---|---|
| Expectedness | | | |
| Expected | 56 | 36 | 92 |
| Unexpected | 119 | 116 | 235 |
| Relatedness | | | |
| Unrelated | 63 | 47 | 110 |
| Unlikely (Remote) | 61 | 68 | 129 |
| Possible | 46 | 32 | 78 |
| Probable | 5 | 5 | 10 |

Table S5. All Adverse Events by Grade

| **Grade** | **RNC** | **UNC** | **Overall** |
|---|---|---|---|
| Mild | 106 | 109 | 215 |
| Moderate | 63 | 30 | 93 |
| Severe | 5 | 10 | 15 |
| Life-threatening | 1 | 3 | 4 |

Table S6. All Adverse Events by MedDRA Category

| MedDRA Category | RNG | UNG | Overall |
|---|---|---|---|
| Blood and lymphatic system disorders | 0 | 1 | 1 |
| Cardiac disorders | 1 | 3 | 4 |
| Ear and labyrinth disorders | 1 | 0 | 1 |
| Endocrine disorders | 2 | 0 | 2 |
| Eye disorders | 0 | 2 | 2 |
| Gastrointestinal disorders | 6 | 9 | 15 |
| General disorders and administration site conditions | 8 | 10 | 18 |
| Immune system disorders | 3 | 4 | 7 |
| Infections and infestations | 35 | 24 | 59 |
| Injury, poisoning and procedural complications | 8 | 6 | 14 |
| Investigations | 0 | 1 | 1 |
| Musculoskeletal and connective tissue disorders | 11 | 14 | 25 |
| Nervous system disorders | 13 | 5 | 18 |
| Pregnancy, puerperium and perinatal conditions | 1 | 1 | 2 |
| Psychiatric disorders | 55 | 35 | 90 |
| Reproductive system and breast disorders | 1 | 0 | 1 |
| Respiratory, thoracic and mediastinal disorders | 23 | 28 | 51 |
| Skin and subcutaneous tissue disorders | 0 | 1 | 1 |
| Social circumstances | 1 | 0 | 1 |
| Surgical and medical procedures | 4 | 4 | 8 |
| Vascular disorders | 2 | 4 | 6 |

There were a total of 13 serious adverse events (SAEs) among randomized participants during the randomized phase of the trial (RNC n=4, UNC n=9). Further details of the reported SAEs are in the tables below.

Table S7. Serious Adverse Events by Expectedness and Relatedness to the Study Cigarette

| | **RNC** | **UNC** | **Overall** |
|---|---|---|---|
| Expectedness | | | |
| Expected | 0 | 1 | 1 |
| Unexpected | 4 | 8 | 12 |
| Relatedness | | | |
| Unrelated | 2 | 4 | 6 |
| Unlikely (Remote) | 1 | 4 | 5 |
| Possible | 1 | 1 | 2 |

Table S8. Serious Adverse Events by Grade

| **Grade** | **RNC** | **UNC** | **Overall** |
|---|---|---|---|
| Moderate | 1 | 0 | 1 |
| Severe | 2 | 6 | 8 |
| Life-threatening | 1 | 3 | 4 |

Table S9. Serious Adverse Events by MedDRA Category

| **MedDRA Category** | **RNC** | **UNC** | **Overall** |
|---|---|---|---|
| Cardiac disorders | 1 | 2 | 3 |
| Infections and infestations | 2 | 1 | 3 |
| Injury, poisoning and procedural complications | 0 | 1 | 1 |
| Musculoskeletal and connective tissue disorders | 0 | 1 | 1 |
| Psychiatric disorders | 1 | 2 | 3 |
| Respiratory, thoracic and mediastinal disorders | 0 | 2 | 2 |

Psychiatric (MedDRA Category) Adverse Events

A total of 91 Psychiatric AEs were reported during the randomized phase of the trial for randomized participants (RNC n=55, UNC n=36).

Table S10. Psychiatric Adverse Events by Expectedness and Relatedness to the Study Cigarette

| | **RNC** | **UNC** | **Overall** |
|---|---|---|---|
| Expectedness | | | |
| Expected | 40 | 25 | 65 |
| Unexpected | 15 | 11 | 26 |
| Relatedness | | | |
| Unrelated | 2 | 1 | 3 |
| Unlikely (Remote) | 20 | 11 | 31 |
| Possible | 29 | 20 | 49 |
| Probable | 4 | 4 | 8 |
